# Polygenic and developmental profiles of autism differ by age at diagnosis

**DOI:** 10.1101/2024.07.31.24311279

**Authors:** Xinhe Zhang, Jakob Grove, Yuanjun Gu, Cornelia K. Buus, Lea K. Nielsen, Sharon A.S. Neufeld, Mahmoud Koko, Daniel S Malawsky, Emma M Wade, Ellen Verhoef, Anna Gui, Laura Hegemann, APEX consortium, iPSYCH Autism Consortium, PGC-PTSD Consortium, Daniel H. Geschwind, Naomi R. Wray, Alexandra Havdahl, Angelica Ronald, Beate St Pourcain, Elise B. Robinson, Thomas Bourgeron, Simon Baron-Cohen, Anders D. Børglum, Hilary C. Martin, Varun Warrier

## Abstract

Although autism has been historically conceptualised as a condition that emerges in early childhood, many autistic people are diagnosed later in life. It is unknown whether earlier and later diagnosed autism have different developmental trajectories and genetic profiles. Using longitudinal data from four independent birth cohorts, we demonstrate that two different socioemotional and behavioural trajectories are associated with age at diagnosis. In independent cohorts of autistic individuals, common genetic variants account for approximately 11% of the variance in age at autism diagnosis, comparable to the contribution of individual sociodemographic and clinical factors, which typically explain less than 15% of this variance. We further demonstrate that the polygenic architecture of autism can be decomposed into two modestly genetically correlated (r_g_ = 0.38, SE = 0.07) autism polygenic factors. One of these factors is associated with earlier autism diagnosis, and lower social and communication abilities in early childhood but is only modestly genetically correlated with ADHD and mental health conditions. Conversely, the second factor is associated with later autism diagnosis, increased socioemotional and behavioural difficulties in adolescence, and has moderate to high positive genetic correlations with Attention-Deficit/Hyperactivity Disorder and mental health conditions. These findings indicate that earlier and later diagnosed autism have different developmental trajectories and genetic profiles. Our findings have important implications for how we conceptualise autism and provide one model to explain some of the diversity within autism.

## Main

Ever since its earliest descriptions in the 1940s^1,2^, autism has been thought of as a condition that emerges in early childhood. However, a greater proportion of autistic individuals are now receiving an autism diagnosis from mid-childhood onwards than in early childhood^3–5^. One factor that may explain these findings is a shift in the conceptualisation of the condition over time, including the recognition that the behavioural signs of autism may not clearly manifest in the first three years of life^6–9^. Supporting this, some studies have demonstrated that a subset of children who do not initially meet the criteria for an autism diagnosis later receive a diagnosis.^7,10–14^ Later autism diagnosis is associated with elevated co-occurring mental health conditions^15,16^, highlighting the need to understand why some autistic people are diagnosed only later in life.

Several social, demographic, and clinical factors have been linked to age at autism diagnosis^17^. However, past studies show that individual clinical and sociodemographic factors explain only a modest proportion (typically less than 15%) of the variance in age at autism diagnosis (**Extended Data Figure 1, Supplementary Table 1**). This suggests additional factors contribute to age at autism diagnosis. One of these additional factors could be genetic differences among autistic individuals. Despite the relatively high heritability of autism^18^, the role of genetics in age at autism diagnosis has not been studied to date.

Two theoretical models can explain how genetics impacts age at autism diagnosis. In the first model, autism has a single polygenic aetiology, with the same set of genetic variants underlying autism, regardless of age at diagnosis (**Unitary Model**, **Extended Data Figure 2**). Under this model, later diagnosed autistic individuals may have subtle clinical features that are harder to recognise early in life, suggesting that they do not cross a diagnostic threshold earlier in life. This may be due to a lower genetic predisposition to autism. As they get older, additional environmental factors may alter their clinical features, eventually bringing individuals above the clinical threshold to receive an autism diagnosis later in life.

An alternative model is that earlier and later diagnosed autism have different underlying developmental trajectories and polygenic aetiologies (**Developmental Model**, **Extended Data Figure 2**). This aligns with existing evidence that the genetic influences on traits related to autism vary across development^19–21^. This model does not preclude a role for environmental factors influencing when someone receives an autism diagnosis, but implies that different sets of genetic variants are associated with earlier and later diagnosed autism.

Here we examined the evidence for these two models through four linked aims (**Extended Data Figure 3**). First, we investigated whether the trajectories of socioemotional and behavioural development are associated with age at autism diagnosis in birth cohorts. Although variable developmental trajectories have been observed among autistic individuals and their younger siblings^22^, it is unclear whether these differences in trajectories are associated with age at diagnosis. Second, we estimated the proportion of variance in age at autism diagnosis explained by common single nucleotide polymorphisms (“SNP-based heritability”). We then tested whether this is attenuated by a range of clinical and demographic factors, as predicted by the Unitary Model. Third, we investigated whether different polygenic factors are associated with earlier and later autism diagnosis, as predicted by the Developmental Model. Fourth, we estimated the genetic correlation between the autism polygenic factors related to age at diagnosis and other mental health and developmental phenotypes.

We provide a summary of the study and address potential questions regarding the implications of the findings in the **Supplementary Summary and FAQs**.

### Behavioural trajectories and diagnosis age

In Aim 1, we investigated whether autistic individuals have varying trajectories of socioemotional and behavioural trajectories and whether these are associated with age at autism diagnosis in three birth cohorts (N = 89 to 188 autistic individuals with recorded age at diagnosis between 5 to 17 years). These are the Millennium Cohort Study (MCS, participants born in 2000), and Longitudinal Study of Australian Children: Kindergarten cohort (LSAC-K, 1999) and Birth cohort (LSAC-B, 2003) (**Supplementary Table 2, Extended Data Figure 4, Supplementary Note 1**). All three cohorts collected longitudinal information on socioemotional and behavioural development using the caregiver-reported Strengths and Difficulties Questionnaire (SDQ)^23^. The SDQ has five subscales (emotional, conduct, hyperactivity/inattention, peer problems, and prosocial behaviours), and the total score of difficulties (hereafter “total difficulties”) is the summed score of the first four subscales. The SDQ is widely used, has excellent psychometric properties^24–26^, and is largely invariant across age, sex, and different populations^27–29^, suggesting that it is measuring the same latent trait across these demographic variables. In addition, the SDQ is moderately correlated with autism-specific measures^30–33^, although it does not capture all of the core diagnostic features of autism. Because not all cohorts recorded the exact age when children received their autism diagnosis, we used the child’s age during the study data collection when caregivers first reported the diagnosis as an approximation of age at autism diagnosis.

To identify latent trajectories, we used Growth Mixture Models of the SDQ total difficulties and subscale scores among autistic individuals in all three cohorts. Growth Mixture Models do not require grouping based on a priori hypothesis, but can identify latent subgroups based on longitudinal differences in SDQ scores.

Across all three birth cohorts, Growth Mixture Modeling identified a two-trajectory model as being optimal for SDQ total difficulties and the majority of subscale scores (**Supplementary Table 3, Supplementary Figures 1 -6**). The first latent trajectory was characterised by difficulties in early childhood that remained stable or modestly attenuated in adolescence (termed ‘early childhood emergent latent trajectory’). The second latent trajectory was characterised by fewer difficulties in early childhood that increased in late childhood and adolescence (termed ‘late childhood emergent latent trajectory’) (**Figure 1 A - C**).

**Figure 1:**
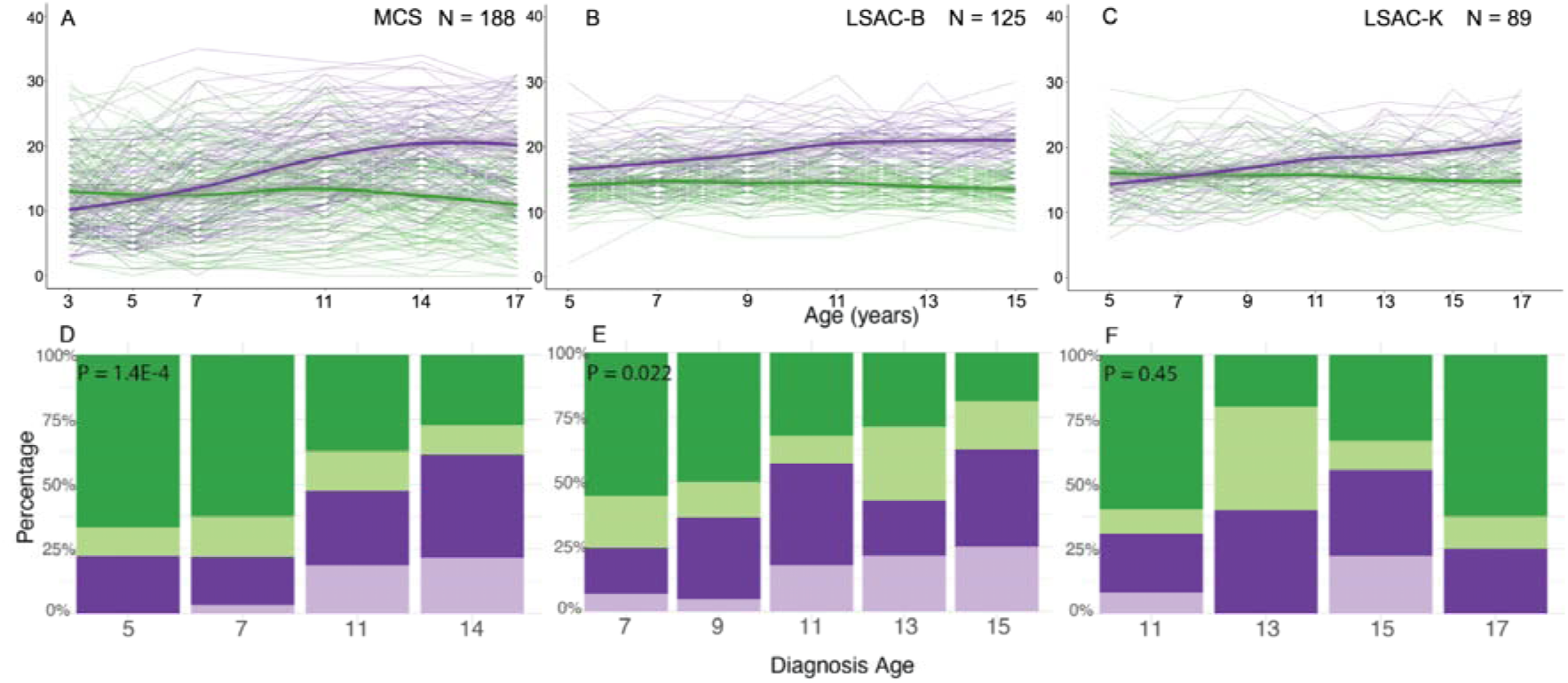
Trajectory analyses in three of the four birth cohorts. A - C: Longitudinal growth mixture models of SDQ total scores among autistic individuals, demonstrating the presence of two groups in the MCS (A), LSAC-B (B) and LSAC-K (C) cohorts. Shaded areas indicate 95% confidence intervals of the line of best fit in D - F: Stacked bar charts providing the proportion of individuals who had been diagnosed as autistic at specific ages, categorised by membership in the latent trajectories identified from the growth mixture models in MCS (D), LSAC-B (E), and LSAC-K (F) cohorts. Darker colours indicate males and lighter colours indicate females. P-values (*P*, inset in panels D-F) are from L^2^ tests (two-sided) comparing the distribution of age at autism diagnosis between the two latent trajectories (pooling the two sexes).

Autistic individuals in the early childhood emergent latent trajectory were more likely to be diagnosed as autistic in childhood compared to autistic individuals in the late childhood emergent latent trajectory in MCS (P = 1.42×10^-4^, L^2^ test) and LSAC-B (P = 2.24×10^-2^, L^2^ test) (**Supplementary Table 4, Figure 1 D - F**). This difference was not significant in LSAC-K, possibly because age 11 was the earliest when an autism diagnosis was recorded.

Sensitivity analyses in MCS confirmed the robustness of the two latent trajectories and their association with age at diagnosis among autistic children. We identified consistent results after expanding the sample to include individuals with co-occurring Attention-Deficit/Hyperactivity Disorder (ADHD) (N = 238, **Supplementary Tables 3 - 4**), and after imputing missing data to increase statistical power and reduce bias (N = 623) (**Supplementary Table 5, Supplementary Notes 2 and 3**). We also obtained consistent results when restricting the analyses to only males (N = 136, **Supplementary Table 3 - 4**), suggesting that these results were not driven by sex differences in age of diagnosis. We were unable to run equivalent female-only analyses due to the low sample sizes.

To assess the specificity of this result to autism, we tested whether similar latent trajectories were also observed in children with ADHD but not autism (N = 89; imputed N = 325) in MCS using Growth Mixture Models. Two latent SDQ trajectory classes emerged, but these were not significantly linked to age at ADHD diagnosis, except for the SDQ hyperactivity/inattention and conduct problems subscales in the imputed sample (**Supplementary Table 6, Supplementary Figures 7 and 8**), suggesting that the findings are relatively specific to autism rather than neurodevelopmental conditions broadly.

Although females receive an autism diagnosis later than males on average^34^, in all three cohorts, the sex ratio was similar between the two latent trajectories (**Supplementary Table 4**), possibly due to their relatively small sample sizes. Individuals in the late childhood emergent latent trajectories were more likely to report mental health conditions (**Supplementary Table 7**), consistent with previous epidemiological observations among later diagnosed autistic individuals^15,16^.

Using multiple regression models, we then examined the extent to which these two latent trajectories contributed to differences in age at autism diagnosis over and above sociodemographic and cognitive characteristics (**Supplementary Table 8**). In these models, SDQ latent trajectories explained 11.7% (LSAC-B) to 30.3% (MCS) of the variance in age of autism diagnosis. In contrast, sociodemographic variables explained 4.8% (LSAC-B) to 5.5% (MCS) of the total variance across cohorts, consistent with previous reports (**Extended Data Figure 1**). In the imputed MCS sample (N = 623, **Supplementary Table 5, Supplementary Note 2**), the SDQ latent trajectories and sociodemographic variables explained 56.6% and 3.2% of the variance, respectively. The effects of the sociodemographic variables were not mediated by the SDQ latent trajectories (**Supplementary Table 8, Supplementary Note 4**).

The associations between different SDQ trajectories and age at autism diagnosis were also supported by latent growth curve models fitted on earlier- and later-diagnosed autistic individuals (**Supplementary Note 5, Supplementary Table 9, Supplementary Figures 1 - 6, 9, and 10**). These results confirm that the association between SDQ trajectories and age at autism diagnosis is robust to methodological choices

### Age at autism diagnosis is heritable

The above analyses demonstrate that variation in socioemotional and behavioural trajectories, measured using the SDQ, is associated with age at autism diagnosis. Previous research has demonstrated that developmental variation traits related to autism is partly explained by genetic factors^19,20,35–38^. A corollary of this is that genetic factors may also be associated with age at autism diagnosis.

Subsequently, in Aim 2, we tested whether age at autism diagnosis is heritable in two large cohorts of autistic individuals using genetic data and information on age at autism diagnosis. This includes (1) the Danish-based iPSYCH cohort (N_total_ = 18,965), a population-based sample derived from the Danish national registries that includes autistic individuals, and (2) the US-based cohort of autistic individuals (SPARK^39^: N_total_ = 28,165; **Extended Data Figures 5 - 6**), which recruits families with at least one autistic individual through online platforms and medical centers across the United States. In SPARK, we conducted initial analyses in a discovery subset of 18,809 autistic individuals (SPARK Discovery), and replicated key findings in an additional sample of 9,356 autistic individuals that only became available after initial analyses were completed (SPARK Replication). SPARK and iPSYCH differed in the diagnostic classification system (iPSYCH: ICD and SPARK: DSM), and median age at diagnosis (iPSYCH: median = 10, median absolute deviation = 4; SPARK: median = 4, median absolute deviation = 2.7).

We conducted Genome-wide Association Study (GWAS) in iPSYCH and across both the Discovery and Replication samples of SPARK, using age at autism diagnosis as a quantitative trait. In all three samples, we identified significant and consistent SNP-based heritability of approximately 11% for age at autism diagnosis (**Figure 2A, Supplementary Table 10**). This is larger than, or similar to, the variance explained by several other clinical and sociodemographic factors tested in SPARK (**Extended Data Figure 1, Supplementary Table 1**).

**Figure 2:**
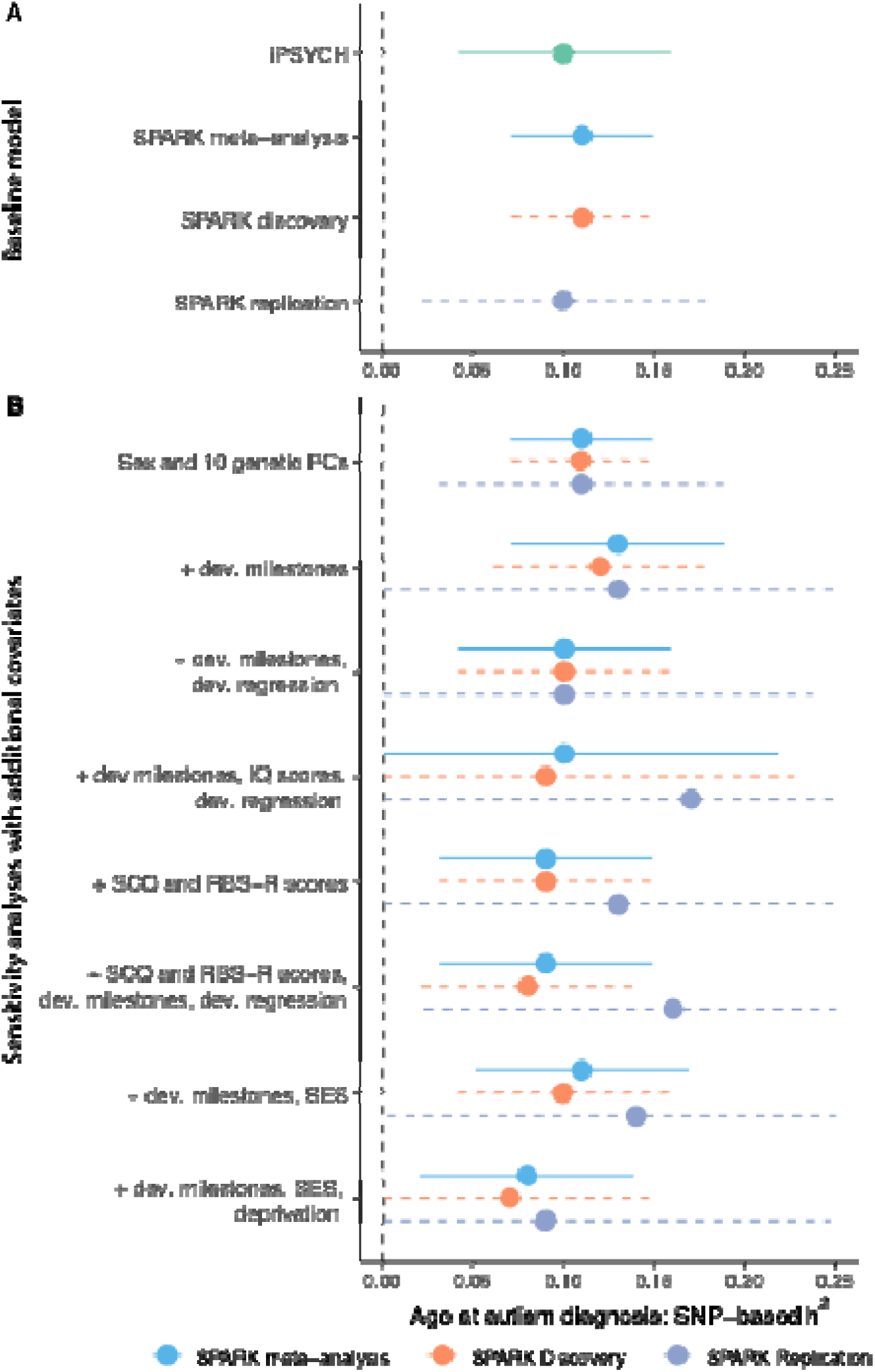
Heritability of age at autism diagnosis. A. SNP-based heritability for age at autism diagnosis in the SPARK (calculated using single-component genome-wide complex trait analysis with genomic-relatedness-based restricted maximum likelihood approach [GCTA-GREML] for the SPARK Discovery cohort (orange dashed line, N = 16,786), SPARK Replication cohort (lilac dashed line, N = 8,558) and a meta-analysis of the two (light blue solid line, N = 25,344)) and iPSYCH (calculated using LDSC, green, N = 18,965). B. SNP-based heritability (GCTA-GREML) in the SPARK cohorts after accounting for various clinical and sociodemographic factors. ‘+’ indicates the baseline model in addition to the specified covariates. The x-axis has been truncated at 0 and 0.25. For A and B, central points represent SNP-based heritability estimates and error bars indicate 95% confidence intervals. Sample sizes for 3B are provided in Supplementary Table 10.

In contrast to the effect of common genetic variants, in a subsample of SPARK with available data for both parents and their autistic child (N = 6,206 trios), we observed no association between age at autism diagnosis and rare *de novo* variants or inherited protein truncating or missense variants in highly constrained genes (**Supplementary Table 11**). This may possibly be due to low statistical power, or reflect later autism diagnosis in some carriers of *de novo* mutations due to diagnostic overshadowing by co-occurring intellectual disability (ID) or global developmental delay^40,41^.

We next tested if this SNP-based heritability of age at autism diagnosis is consistent with either of the two theoretical models outlined earlier. The Unitary Model assumes that later diagnosis reflects subtle or less severe clinical features, and so the SNP-based heritability of age at autism diagnosis may simply reflect the severity of autism features. Alternatively, the SNP-based heritability may reflect additional genetic influences associated with co-occurring developmental delays, developmental regression, or ID, which may lead to an earlier diagnosis. It may also reflect the heritable component of parental socioeconomic status and neighbourhood deprivation - proxies for parental awareness and healthcare access that affect diagnostic timing. Controlling for any of these measures should attenuate the SNP-based heritability.

In contrast, the Developmental Model assumes that SNP-based heritability of age at autism diagnosis reflects a mixture of different polygenic factors that are correlated with age at diagnosis but independent of these covariates. Under this model, a significant SNP-based heritability should persist after controlling for clinical, developmental and sociodemographic measures.

In line with the Developmental Model, we found that the SNP-based heritability did not significantly attenuate after controlling for parental sociodemographic measures, clinical features, or co-occurring developmental delays and conditions (**Figure 2B, Supplementary Table 10**). This is inconsistent with the Unitary Model, although imperfect and incomplete measurement of clinical and developmental phenotypes may limit this conclusion.

A second prediction of the Unitary Model is that earlier diagnosis is associated with greater polygenic propensity for autism compared to later diagnosis (**Extended Data Figure 2**). Under this model, all autistic individuals would have higher polygenic propensity for autism compared to non-autistic controls. This would result in negative genetic correlations between GWAS of age at autism diagnosis and GWAS of autism, with the magnitude of this negative correlation decreasing as the median age at diagnosis in the autism GWAS samples increases.

We tested this prediction using 13 different but partly overlapping autism GWASs including six GWASs stratified by age at diagnosis and two GWASs stratified by sex (**Box 1**, **Figure 3**). The genetic correlation between age at autism diagnosis and different autism GWASs varied systematically, becoming increasingly positive as the median age at diagnosis increased (**Figure 3, Supplementary Table 12**). However, contrary to the expectation under the Unitary Model, we observed positive genetic correlations between age at autism diagnosis and autism GWAS comprising later-diagnosed autistic individuals. These findings support the existence of different genetic architectures across diagnostic age groups, aligning with the Developmental Model.

**Figure 3:**
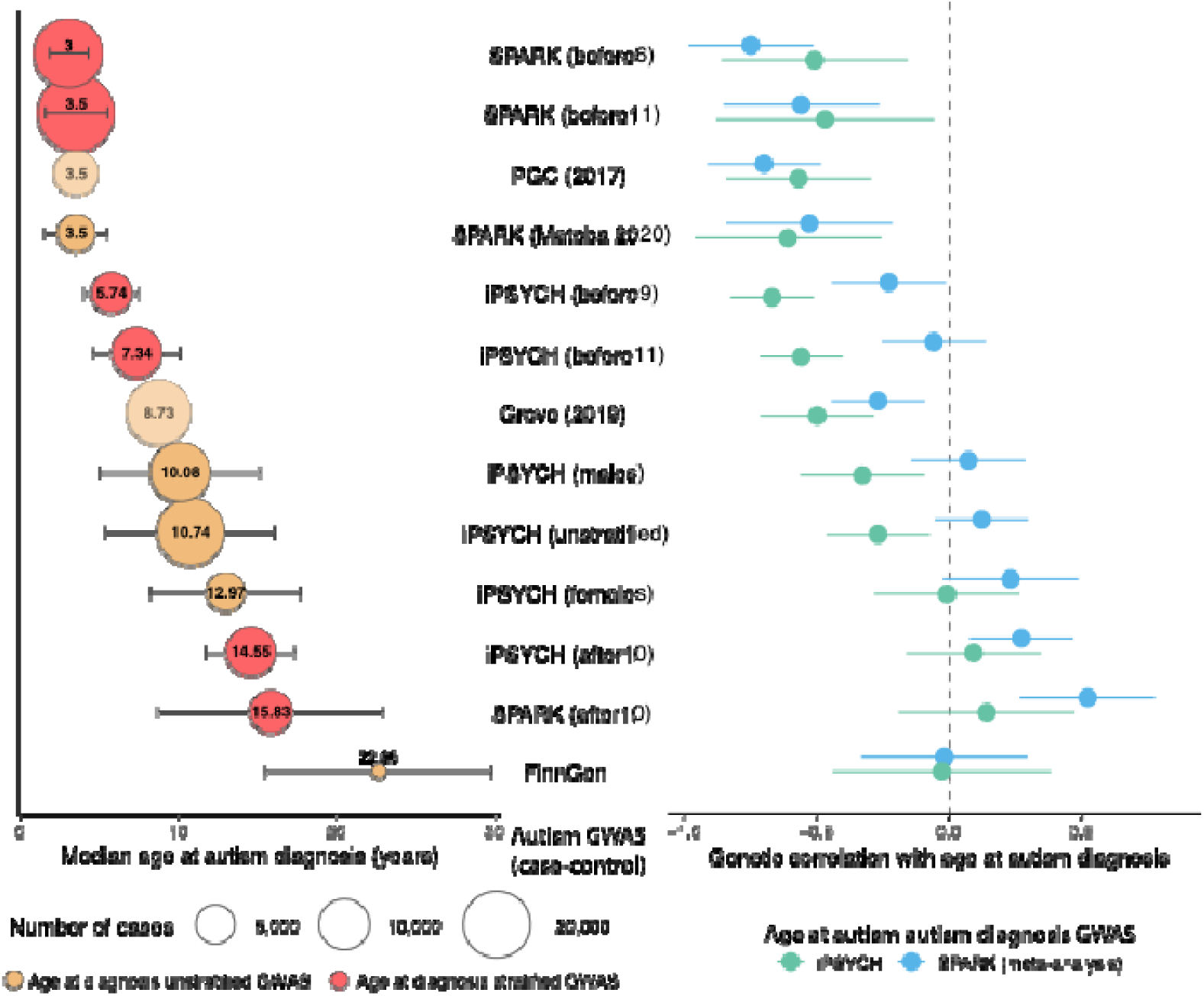
Median age at autism diagnosis and genetic correlations with age at autism diagnosis across different GWAS cohorts. Left panel: Median age at diagnosis (years) with error bars representing median absolute deviation. Circle size indicates the number of autistic individuals (cases) in the GWAS, and exact sample sizes are provided in Box1. Beige circles represent GWAS unstratified by age at diagnosis. Red circles represent GWAS stratified by age at diagnosis. Lighter, more transparent circles indicate studies with no information about age at autism diagnosis, and the median ages have been inferred based on other available information (i.e., PGC and Grove). Right panel: Genetic correlations with age at autism diagnosis for both SPARK (blue, meta-analysis, N = 28,165) and iPSYCH (green, N = 18,965) datasets, with error bars representing 95% confidence intervals.

Furthermore, the male- and female-stratified autism GWAS from iPSYCH had similar genetic correlation with age at autism diagnosis. This suggests that the pattern of genetic correlation between age at diagnosis and the autism GWASs does not reflect differences in the sex ratio of participants across the autism GWASs.

The genetic correlation with some of the autism GWASs differs significantly between the age at diagnosis GWASs in iPSYCH versus meta-analysed SPARK (**Figure 3**). This reflects the fact that these two age at diagnosis GWASs are only moderately genetically correlated with each other (Genetic correlation (r_g_)= 0.51, standard error (SE) = 0.19, P = 7.56×10^-3^), which is possibly due to the different recruitment strategies and resulting differences in the median age at autism diagnosis in the two cohorts (**Supplementary Note 6**).

### Box 1: Summary of the autism GWAS used in this study

Autism GWAS not stratified by age at diagnosis

- SPARK (Matoba 2020): Case-pseudocontrol design (4,535 pairs) with family-based ascertainment across the United States. Median age at diagnosis = 3.5 (1.97) years
- FinnGen (Data Release r10): Population-based sample from Finland (646 cases vs. 301,879 controls). Median age at diagnosis = 22.66 (7.16) years.
- PGC-2017: Meta-analyses of several case-control and case-pseudo control datasets (7,387 cases vs. 8,567 controls). The majority of the cases met the diagnostic criteria for autism under DSM-IV-TR/ICD-10 or earlier (i.e., onset of features before age 3) after screening using the Autism Diagnostic Observation Schedule and the Autism Diagnostic Interview-Revised), making this a clinically well characterised cohort. Although age at diagnosis is unavailable, the majority of the participants were recruited as trios through medical or research centres in the United States. Given this similarity in ascertainment to SPARK, we anticipate the age at diagnosis to be similar to that of SPARK trios (Matoba 2020). Approximate median age at diagnosis = 3.5
- iPSYCH (unstratified): Population-based sample derived from Danish national registries, including individuals born 1980-2008 (19,870 autistic vs. 39,078 non-autistic individuals). Median age at diagnosis = 10.74 (5.35)
- iPSYCH (males): Males only subset of iPSYCH (15,025 autistic vs. 19,763 controls). Median age at diagnosis = 10.08 (5.07)
- iPSYCH (females): Females only subset of iPSYCH (4,845 autistic vs. 19,315 controls). Median age at diagnosis = 12.97 (4.75)
- Grove et al., 2019: Meta-analysis of a subset of the iPSYCH and PGC samples (18,381 cases vs. 27,969 controls). As age at diagnosis for PGC is unavailable, we calculated an estimated age by weighing the median age at diagnosis of iPSYCH with the estimated median age in PGC by their respective sample size. Approximate median age at diagnosis = 8.73.

Autism GWAS stratified by age at diagnosis

#### SPARK (using unaffected family members as controls), meta-analysed from the Discovery and Replication subsets

- Diagnosed before age 11 (SPARK_before11_): 27,881 autistic individuals; selected to match iPSYCH stratification; median age at diagnosis = 3.5 (1.97) years. This cutoff period was chosen to reflect the cutoff used in the Latent Growth Curve models, and represents a time window characterised by the onset of puberty, transition from primary to secondary school, and an increase in the number of autistic girls being diagnosed.
- Diagnosed after age 10 (SPARK_after10_): 6,243 autistic individuals; median age at diagnosis = 15.83 years (7.16).
- Diagnosed before age 6 (SPARK_before6_): 21,435 autistic individuals; categorised as "early-diagnosed" based on previous research^8^; median age at diagnosis = 3 (1.23) years.

#### iPSYCH (population-based controls)

- Diagnosed before age 11 (iPSYCH_before11_): 9,500 autistic vs. 36,667 non-autistic individuals; median age at diagnosis = 7.34 (2.76) years. The cutoff was chosen to reflect the cutoff used in the Latent Growth Curve models and SPARK age at diagnosis stratified GWAS.
- Diagnosed after age 10 (iPSYCH_after10_): 9,231 autistic vs. 36,667 non-autistic individuals; median age at diagnosis = 14.55 (2.84) years.
- Diagnosed before age 9 (iPSYCH_before9_): 5,451 autistic vs. 36,667 non-autistic individuals; created to provide additional age resolution; median age at diagnosis = 5.74 (1.69) years

### Two autism polygenic factors

These findings suggest that the age at autism diagnosis reflects a mixture of different age-dependent polygenic traits (Developmental Model) rather than a single polygenic trait (Unitary Model). Under the Developmental Model, one would expect that the genetic correlations between different autism GWAS will differ according to the difference in the median age of diagnosis between the GWAS.

In Aim 3, we tested this by estimating genetic correlations among the 13 autism GWASs (**Box 1**). We observed genetic correlations ranging from 0.02 (SE = 0.13) to 1 (SE = 0.01) (**Figure 4A, Supplementary Table 13**). We observed a gradient in the genetic correlations related to the similarity in median age at diagnosis between cohorts. Cohorts with the most similar median ages at diagnosis (differing by ≤2 years) showed the highest genetic correlations (r_g_ = 0.88-1), while correlations progressively decreased as age differences increased. The lowest genetic correlation of (r_g_ = 0.02, SE = 0.13) was observed between SPARK_before6_ (median age ∼ 3) and SPARK_after10_ (median age ∼ 16).

**Figure 4:**
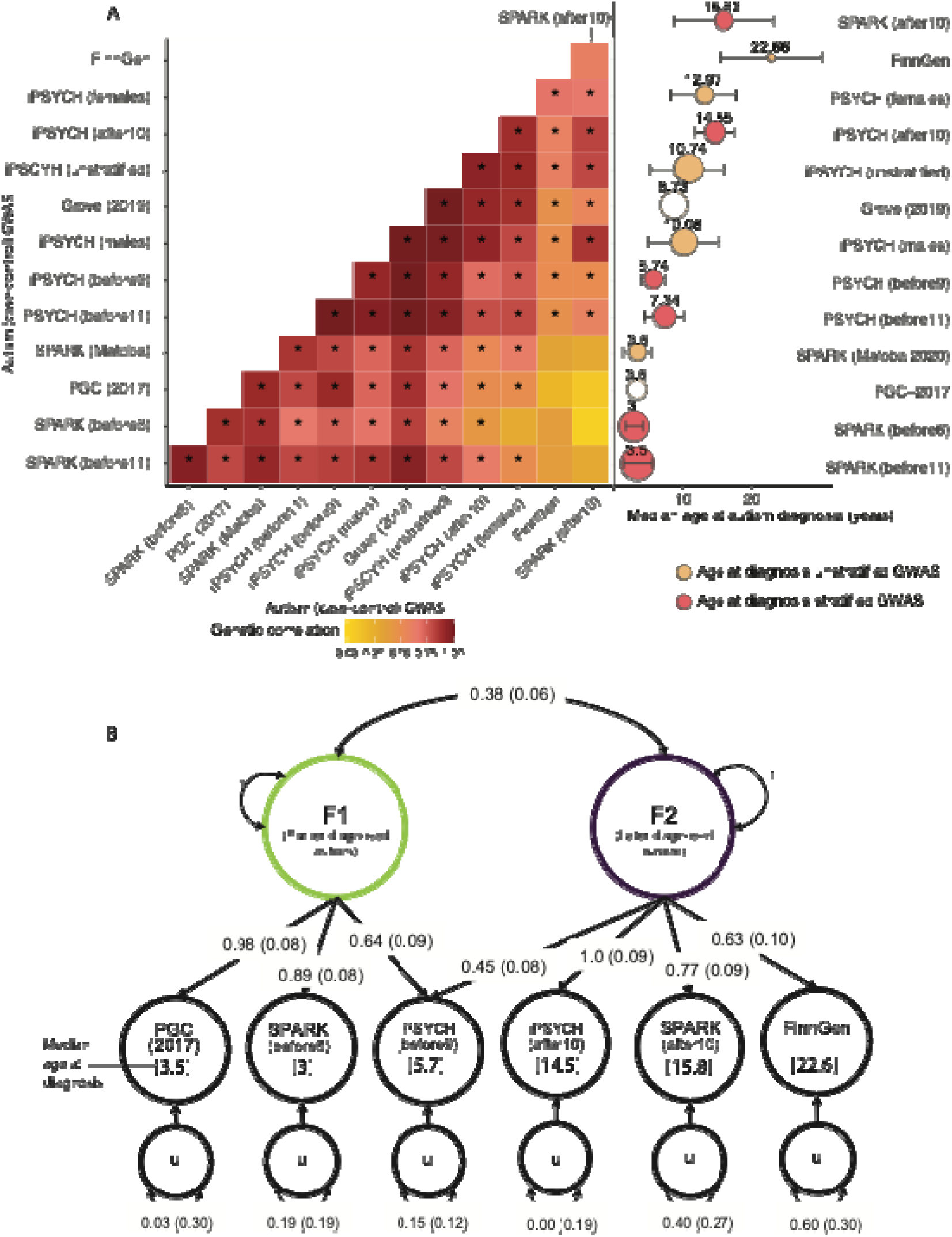
Two genetic latent factors in autism. A. Genetic correlation heatmaps of all GWAS of autism (left). Asterisk indicates significant genetic correlations after Benjamini-Yekutieli adjustment. The right panel provides median age at autism diagnosis for the same GWAS (indicated by the number on top of the circle). Error bars indicate median absolute deviation. Size of the circles indicate sample size of cases. For both panels, GWAS have been ordered based on hierarchical clustering of the genetic correlations. B. Structural Equation Model illustrating the two-correlated-genetic-factor models for autism, using six minimally overlapping autism GWAS datasets. F1 = Factor 1, F2 = Factor 2. One-headed arrows depict the regression relationship pointing from the independent variables to the dependent variables; the numbers on the arrows represent the regression coefficients of the factor loadings, with the standard errors provided in parentheses. Covariance between variables are represented as two-headed arrows linking the variables. The numbers on the two-headed arrows can be interpreted as genetic correlation estimates with the standard errors provided in parentheses. Residual variances for each GWAS dataset are represented using a two-headed arrow connecting the residual variable (u) to itself. Standard errors are provided in parentheses.

Hierarchical clustering of the genetic correlations identified two broad, overlapping clusters that differed by age at autism diagnosis. One cluster comprised GWAS of autism in cohorts with predominantly childhood diagnosed individuals, and the other comprised GWAS of autism in cohorts with a large fraction of individuals diagnosed in adolescence or later, consistent with predictions from the Developmental Model.

We formally tested this by modelling the genetic covariance using structural equation models in GenomicSEM^42^, testing six theoretical models (**Supplementary Table 14**). Using six minimally overlapping GWAS for autism with wide variation in age at autism diagnosis among those listed in Box 1. We found that a correlated two-factor model was the most parsimonious and fit the data best (Akaike information criterion: 38.64, confirmatory fit index : 0.99, standardised root mean residual: 0.08, **Figure 4B**). Factor 1 (Earlier diagnosed autism factor) was defined by the GWAS with predominantly early childhood diagnosed individuals (PGC-2017, SPARK_before6_, with a median age at diagnosis of 3). Factor 2 (Later diagnosed autism factor) was defined primarily by GWAS with adolescent or adult diagnosed individuals (iPSYCH_after10_, FinnGen, and SPARK_after10_). The cross loading of iPSYCH_before9_ (median age at diagnosis ∼5.7) suggests that Factor 2 may impact behaviours in mid/late childhood as well. The two factors had a modest genetic correlation (r_g_ = 0.38, SE = 0.06). Sensitivity analyses confirmed the robustness of the above results using partly different GWAS, where we identified a two-correlated-factor model as the best fitting model, with similar moderate genetic correlations between the two factors (r_g_ = 0.37, SE = 0.06 - 0.52, SE = 0.10, **Supplementary Table 14**).

The earlier diagnosed autism factor was negatively genetically correlated with age at autism diagnosis (**Figure 5**). The later diagnosed autism factor was positively genetically correlated only with age at autism diagnosis from SPARK. Genetic correlation with the autism GWAS stratified by sex showed that both autism factors had stronger genetic correlations with autism in males than females, with a larger difference for the earlier diagnosed autism factor (**Figure 5**), consistent with established sex differences in age at autism diagnosis.

**Figure 5:**
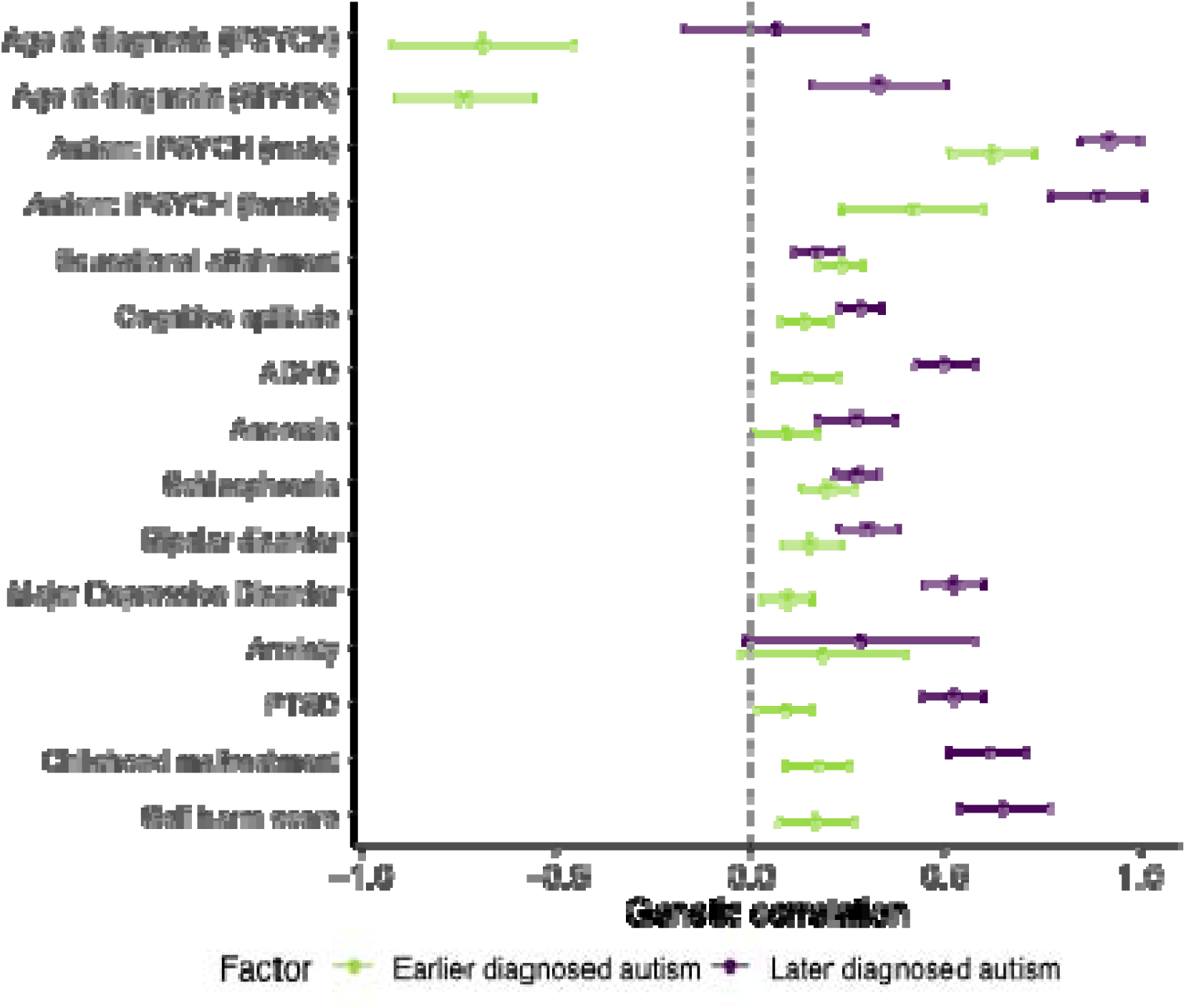
Genetic correlation between the two autism polygenic factors and a range of mental health, neurodevelopmental, and cognitive traits. Central points indicate the estimate (genetic correlation), error bars indicate 95% confidence intervals, and points with an asterisk (*) indicate significant p-values (two-sided) with Benjamini-Yekutieli adjustment. Sample sizes are provided in Supplementary Table 18.

Additional analyses using polygenic scores confirmed that the association between the two polygenic autism factors and age at autism diagnosis is not due to several confounding factors using within-family approaches, nor due to differences in clinical and demographic factors, and co-occurring developmental conditions (**Supplementary Note 7, Supplementary Table 15 - 17**). Taken together, the above findings demonstrate that earlier and later diagnosed autism have different polygenic aetiologies, supporting the Developmental Model.

### Genetic correlations with autism factors

The above analyses indicate that there are at least two polygenic factors associated with age at autism diagnosis. Since later-diagnosed autistic individuals have higher rates of mental health conditions^15,16^, we hypothesised this might partly be because earlier and later diagnosed autism factors have differing genetic correlations with mental health and cognitive phenotypes. In Aim 4, we investigated this hypothesis using genetic correlation analyses.

The earlier diagnosed autism factor (Factor 1) had modest (r_g_ ∼ 0.1-0.2) but significant genetic correlation with educational attainment, cognitive aptitude, ADHD, various mental health and related conditions (**Supplementary Table 18, Figure 5**). The later diagnosed autism factor (Factor 2) showed a statistically similar genetic correlation with educational attainment but significantly higher genetic correlations (r_g_ ∼ 0.5-0.7) with ADHD and a range of other mental health and related conditions, including Depression, PTSD, childhood maltreatment, and self-harm. After accounting for genetic effects on ADHD, we saw an attenuated but significant moderate genetic correlation between later diagnosed autism (Factor 2) and mental health conditions, suggesting shared genetics with ADHD do not fully explain the elevated correlation between later diagnosed autism and mental health phenotypes (**Supplementary Table 18, Supplementary Figure 11**). Sensitivity analyses using age at diagnosis stratified GWAS from iPSYCH and SPARK yielded largely consistent genetic correlation results, suggesting that these results are not due to cohort differences (**Supplementary Table 19, Extended Data Figure 7**).

The higher genetic correlation between later diagnosed autism and other mental health conditions may indicate diagnostic misclassification (where individuals with other conditions incorrectly receive an autism diagnosis) or diagnostic overshadowing (where the presence of co-occurring mental health conditions can delay an autism diagnosis). Under the Unitary Model, the genetics of later diagnosed autism would reflect the additive genetic effects of earlier diagnosed autism and of other mental health conditions. However, decomposition of the autism genetic signal using genomicSEM indicated that later diagnosed autism cannot be entirely attributed to the polygenic effects of earlier diagnosed autism and six other mental health conditions tested: schizophrenia, ADHD, anorexia nervosa, depression, bipolar disorder, and PTSD (**Supplementary Note 8**). Thus, the later diagnosed autism genetic factor does not represent the additive genetic effects of earlier diagnosed autism and mental health conditions.

Finally, we investigated whether the two autism polygenic factors related to age at diagnosis differ in their associations with developmental traits. Cross-sectionally, these genetic factors showed few significant differences in genetic correlation or polygenic score association with developmental phenotypes measured at age three or earlier. The exceptions were age at onset of walking^43^ and expressive vocabulary at 2 - 3 years^44^, with which the earlier diagnosed autism factor was positively genetically correlated, but the later diagnosed was not (**Supplementary Note 9, Supplementary Table 20 - 22**). However, longitudinal polygenic score analyses across two birth cohorts revealed differential genetic effects of earlier versus later diagnosed autism factors on SDQ total difficulties scores over time (**Supplementary Note 9, Supplementary Table 23**).

## Discussion

These results indicate that earlier and later diagnosed autism are associated with different developmental trajectories, and are only modestly genetically correlated with each other. This possible framework provides one axis of heterogeneity to describe the widely acknowledged clinical and genetic heterogeneity within autism, that thus far has been challenging to identify. This finding of two or more developmentally variable polygenic latent traits for autism is robust to various observed clinical and demographic factors (**Supplementary Note 10**), including sex and ID.

Consistent with the larger literature on developmental variation in autism^22^, our analyses of socioemotional and behavioural trajectories across multiple birth cohorts converge with the genetic findings - polygenic scores for earlier and later diagnosed autism have different associations with developmental changes in SDQ total difficulties scores (**Supplementary Note 9**). These results suggest that the timing of autism diagnosis may partly reflect etiologically different developmental pathways, rather than purely environmental or diagnostic factors. Our findings are consistent with the wider literature that demonstrates that genetic influences on traits related to autism vary across development in the general population^19,20^.

This two-polygenic trait genetic model provides one framework to understand genetic heterogeneity in autism, and the varying patterns of genetic correlations between different GWAS of autism and other phenotypes. For example, previous GWAS of autism (including PGC-2017) found limited genetic correlation with ADHD, contrary to findings from more recent autism GWAS (e.g., Grove et al., 2019). We show that this is explained by the different average age at diagnosis across these GWAS (**Box 1**, **Figure 4**), as the genetic correlation between autism and ADHD increases with later age at autism diagnosis (**Figure 6**). These findings were confirmed using within-family analyses that demonstrated over-transmission of ADHD polygenic scores primarily to individuals with a later autism diagnosis (**Supplementary Table 24**).

Both the later diagnosed genetic autism factor (**Figure 5**) and the late childhood emergent latent trajectory of SDQ total difficulties scores (**Figure 2**) are associated with greater mental health problems (**Figure 6 and Supplementary Table 7**). This suggests that epidemiological findings of greater mental health difficulties among later diagnosed autistic individuals^15,16^ may be partly explained by the Developmental Model of autism. Given that autistic females are, on average, diagnosed later in life, research that investigates sex and gender differences in both autism and co-occurring conditions^16,46^ needs to account for genetic confounding associated with age at autism diagnosis. Findings that may appear to reflect sex differences may additionally reflect differences associated with age at diagnosis. For example, the higher prevalence of mental health problems in autistic females^16,46^ compared to males attenuates when restricting to autistic individuals diagnosed before age five^16^.

These findings must be interpreted considering several limitations. First, the SNP-based heritability for age at autism diagnosis is only about 11% (**Figure 3**), and other observed developmental and demographic factors typically explain less than 15% of the variance (**Extended Data Figure 2**). This suggests that there are several other factors that contribute to age at autism diagnosis. We find that the genetic effects on age at autism diagnosis are not mediated by several of these measured developmental and demographic factors, but we acknowledge that there may be several unmeasured factors that may mediate the genetic effects. Furthermore, the substantial variation across the datasets explored highlights that age at autism diagnosis is immensely complex, and varies across geography and time. Local cultural factors, access to healthcare, gender bias, stigma, ethnicity, and camouflaging likely have an impact on who receives a diagnosis and when. Second, our trajectory models (**Figure 2**) were built using only the SDQ, which measures a wide range of parent-reported neurodevelopmental and mental health traits. Although the SDQ is correlated with an autism diagnosis, it does not fully capture core autistic traits, and other measures of autistic traits were not available in the birth cohorts. Third, it is likely that other dimensions contribute to genetic heterogeneity in autism. For example, a significant proportion of the variation in the FinnGen autism GWAS was not explained by either of the two factors (**Figure 5**). Fourth, we use ’earlier’ and ’later’ diagnosed autism as relative terms, reflecting that developmental and polygenic differences represent a gradient (**Figure 5, Supplementary Figure 10**) rather than discrete categories, and because there is no consensus on age thresholds for early versus late diagnosis^47^. Fifth, autism diagnoses in the birth cohorts used in the current study rely on community-based caregiver or self-reporting rather than standardised clinical assessments. As such, there may be varying delays between the emergence of autistic features and a formal autism diagnosis. However, our findings can guide future research using longitudinal cohorts, and particularly sibling studies, that systematically track the emergence of autism features over time. Finally, our genetic analyses focused on common genetic variants in genetically inferred European ancestries due to limited GWAS data from other populations, highlighting the need for future research to examine the transferability of these findings across diverse genetic ancestries.

In conclusion, we find that the developmental trajectories and polygenic architecture of autism varies with age at diagnosis. These findings partly explain the varying genetic correlations among the different GWAS of autism and between autism and various mental health conditions. These findings provide further support for the hypothesis that the umbrella term “autism” describes multiple phenomena with differing aetiologies, developmental trajectories, and correlations with mental health conditions. These findings have implications for how we conceptualise neurodevelopment more broadly, and for understanding diagnosis, sex and gender differences, and co-occurring health profiles in autism.

## Supporting information

Supplementary Notes, Figures, and FAQs

## Methods

### A note on terminology

We use the term autistic and non-autistic to refer to people with and without an autism diagnosis^48^. Sex, males, and females refer to sex assigned at birth.

### Variance explained in age at autism diagnosis: literature review and analyses of the SPARK cohort

To contextualise the SNP-based heritability and the variance explained by SDQ total difficulties and subscale scores, we conducted a review of the variance in the age at autism diagnosis explained by various sociodemographic and clinical factors, including sex and autism severity. Using Google Scholar and PubMed, we searched for studies published between 1998 and December 10, 2024 using combinations of the following terms in the title or abstract: "age at diagnosis" AND “autism”, "autism” AND “age," and "diagnosis age” AND "autism”. We also used these search terms with alternative terminology for autism, including "autism spectrum condition," "autism spectrum disorder," and "ASD". This search resulted in over 1,700 studies. Manual review identified 184 studies that investigated factors associated with age at autism diagnosis. Of these 13 quantified the variance explained using measures such as R^2^ or η^2^, and were included in our final analyses (**Supplementary Table 1**).

In addition, we also calculated the variance explained in the US-based cohort of autistic individuals and their families, SPARK^39^, where we used the v9 release of the phenotypic data. We focussed on the variance explained by sociodemographic factors (sex, reported race, household income, mother’s education, and father’s education), cognitive and developmental factors (reported IQ score, reported intellectual disability, age at walking independently, age at first words, language regression, other regression), and autism severity (scores on the Social Communication Questionnaire and Repetitive Behaviour Scale-Revised). After excluding individuals with missing data, we quantified the variance explained by these factors using relative importance analysis (Lindemann, Merenda and Gold - 1980 method) in 5,773 autistic individuals diagnosed before age 22. Thereafter, analyses were conducted using the *relaimpo* (v2.2-7) package in R, which allowed us to examine all variables’ contributions simultaneously ^49^.

### Trajectory analyses of birth cohorts

#### Cohorts

We used four population-based birth cohorts that vary both in the ages at which data was collected from participants and the calendar years of data collection (**Extended Data Figure 3**). Briefly, the four cohorts included are the UK-based Millennium Cohort Study (MCS)^50^, the Australia-based Longitudinal Study of Australian Children - Birth (LSAC-B) and Kindergarten (LSAC-K) cohorts^51,52^, and the Ireland-based Growing Up in Ireland (GUI) Child cohort (aka Cohort 98’)^52,51^. All children included in the cohorts were born in the 21st century. Further details about the cohorts are provided in the **Supplementary Note 1.** We used data from MCS, LSAC-B, and LSAC-K for the Growth Mixture Models. We did not use data from GUI for Growth Mixture Models as there were only three time points, which is not sufficiently powered to identify two or more trajectories. All four cohorts were used for Latent Growth Curve Models.

As indicated in **Supplementary Table 2**, these cohorts were selected because they were (1) longitudinal in nature, (2) nationally representative, and (3) included key data on behavioural profiles and neurodevelopmental diagnosis. These overlapping features across datasets allowed for cross-country comparisons and generalisation^53^.

### Measures

#### Autism and ADHD diagnosis and age at diagnosis

In all cohorts, across multiple sweeps, the main caregiver was asked if the participant had a diagnosis of autism (**Extended Data Figure 3**). For age at diagnosis, we used the age at the sweep when caregivers first reported their child’s autism diagnosis in every cohort, to maximise sample sizes and ensure consistency across cohorts for effective comparisons. For instance, if an autism diagnosis was reported for the participant for the first time at the age 11 sweep, we considered age at diagnosis to be 11 years. Although the specific age at diagnosis was provided for LSAC-B and LSAC-K, we opted not to use this, as we identified errors in some reports where months and years of diagnosis were swapped or not reported.

In MCS, we used reports of both autism and ADHD diagnoses to conduct additional sensitivity analyses. For our primary analyses, we included a narrowly defined sample of children with consistently reported autism diagnoses by primary and proxy caregivers (when both were available) and no other reported neurodevelopmental diagnosis (particularly ADHD). To assess the generalisability of our results and increase the sample size, we then expanded the sample to include all children with any reported diagnosis of autism (results reported in **Supplementary Note 3**). This expanded sample included cases regardless of whether the diagnoses were consistent across sweeps or caregivers, and included those with co-occurring ADHD. Additionally, we imputed the independent variables and covariates for autistic individuals with missing information, as detailed below (more details in **Supplementary Note 2** and **Supplementary Note 3**). Finally, to assess the specificity of the trajectories for autism, we conducted analyses among children who had a consistent ADHD diagnosis but no diagnosis of autism. Imputation was also performed within this ADHD-only sample, to increase the sample size.

#### Strengths and Difficulties Questionnaire (SDQ)

We used the SDQ to capture social, emotional, and behavioural profiles of participants, with repeated measures from 3 to 18 years across cohorts (**Extended Data Figure 3**). SDQ comprises 25 statements that caregivers are asked to rate on a 3-point Likert scale (“not true”, “somewhat true”, and “certainly true”) based on the child’s symptoms or behaviours over the past six months. There are five subscales, each containing five items, which assess emotional symptoms, conduct problems, hyperactivity-inattention, peer relationship problems, and prosocial behaviours respectively^23^. The first four subscales assess difficulties, and their combined total score ranges from 0 to 40, with higher scores indicating greater difficulties. The fifth subscale (prosocial behaviours) represents strengths and ranges from 0 to 10, with higher scores indicating more prosocial behaviours. We analysed the total score and each subscale separately. The SDQ demonstrates good test-retest reliability and criterion validity across countries^24–26^. Its five-factor structure (each subscale as a factor) has shown consistency and invariance across age, sex, and ethnic background^24,28^. The SDQ captures several core features of mental health and neurodevelopmental conditions, including autism and ADHD^54^. Only children with complete data of SDQ across all sweeps were included in the analyses, except for imputation analyses.

#### Sociodemographic measures

Sociodemographic measures were included as covariates to account for their impact on age at diagnosis in each cohort (**Supplementary Table 25**). Specific measures and available information vary across cohorts, but we generally included sex, ethnic background, maternal age at delivery, child’s cognitive aptitude, household socio-economic status (SES), and deprivation level of the living area, to account for factors that may impact the age when someone receives an autism diagnosis^55,56^. Only subsets of children in the complete-SDQ samples, with complete data for these sociodemographic factors, were included in the respective analyses.

In MCS, although various census classifications for ethnic groups were available, we opted to use a binary indicator to identify non-white ethnic minorities. Ethnicity data were not collected in either LSAC cohort. Instead, visible ethnic minority status was determined primarily by parental country of birth and the language(s) spoken at home.^57^ Maternal age at delivery was collected only in MCS. In other cohorts, we used maternal age (in years) at first sweep of data collection to reflect the variation in maternal age at delivery.

In MCS, we identified multiple variables linked to cognitive ability, SES, and area deprivation. Similarly, in LSAC-B, we identified multiple measures linked to SES, although there were no measures with sufficient sample size linked to cognitive ability or area deprivation. Subsequently, we conducted principal component analysis (PCA) for cognitive abilities, SES, and area deprivation in MCS, and for SES in LSAC-B. PCA was performed using a wide range of measures collected across sweeps (**Supplementary Table 25**), with one variable excluded from any pair with a correlation coefficient greater than 0.70 to address multicollinearity. The first principal component (PC1) explained more than 40% of the variance for each corresponding factor. In contrast, subsequent components contributed substantially less, supporting the use of the respective PC1 as the summary measure for cognitive ability, SES, and area deprivation (**Supplementary Table 25**).

Intellectual disability (ID) was defined as scoring two standard deviations below the mean on the PC1 of the cognitive aptitude factor, consistent with prior studies. No autistic children in the MCS or LSAC cohorts who had measures of cognitive aptitude met the criteria for ID, likely due to participation bias. All PCA analyses were conducted in R using the *prcomp()* function ^58^.

### Statistical analyses

#### Growth Mixture Models and Latent Growth Curve Models

We used two methods to model the longitudinal trajectories of SDQ total and subscale scores. First, we conducted Growth Mixture Models to identify whether there were latent groups of autistic individuals based on their trajectories of SDQ total and subscale scores. Growth Mixture Models assume that the sample consists of multiple mixed effects models, each capturing a subgroup trajectory with shared intercept and slope^59^. We fitted models with one to four groups for each subscale and SDQ total scores in each cohort, using the *lcmm (v2.1.0)* package in R^60^. The optimal number of latent trajectories were then determined by comparing fit indices, including Bayesian Information Criterion (BIC) values, classification quality measure (entropy), and substantive interpretation. Models with lower BIC values and higher entropy were favoured^61^. Models identifying subgroups with less than 5% of the sample size were not considered due to poor statistical reliability and limited practical significance^62^. We compared the distribution of group memberships across diagnostic ages and across sexes using L^2^ tests.

Second, we used linear Latent Growth Curve Models to identify the latent trajectories of SDQ total and subscale scores in the three groups (childhood diagnosed, adolescent diagnosed, and the general population) for all cohorts. Each linear model included a latent intercept to represent the initial level of the outcome variable, and a linear latent slope to represent the mean rate of change over time. Childhood diagnosed (diagnosed before ages 9 - 11, depending on the cohort), and adolescent diagnosed (diagnosed after the ages of 9 - 11, depending on the cohort) were *a priori* defined (see **Extended Data Figure 3**). We chose this 9 - 11 age window as our cutoff since it aligns with the onset of puberty, the transition from primary to secondary school, and aligns with epidemiological evidence showing increased autism incidence among females during this window^34^. An earlier cutoff was not feasible as only MCS (ages 5, 7) and GUI (age 7) recorded autism diagnoses before this window. A later cutoff was not possible due to no autism diagnoses in MCS after age 14. To further examine the relationship between age at diagnosis and socioemotional and behavioural outcomes, we conducted additional Latent Growth Curve Models for autistic children using stepwise groupings by age at diagnosis in MCS and LSAC-B (see **Supplementary Note 5** and **Supplementary Figure 10**). All individuals who lacked an autism diagnosis (and additionally, an ADHD diagnosis in MCS) were included in the general population group.

Given the well-known sex differences in age at autism diagnosis^34^, we also applied the same models stratified by sex, i.e., estimating latent intercept and slope for each sex, within the autistic samples. All Latent Growth Curve Models were fitted under the structural equation modelling framework using the *lavaan* (v 0.6-19) package in R^63^.

#### Association with sex and mental health phenotypes

To examine the association between GMM-derived SDQ latent trajectories and mental health phenotypes in MCS, LSAC-B, and LSAC-K, adjusting for sex), we applied multiple regression in autistic individuals.

#### Variance explained in age at autism diagnosis and mediation analysis in birth cohorts

Multiple regression analyses were conducted in MCS, LSAC-B and LSAC-K to investigate the association between an age at autism diagnosis (the outcome variable), SDQ total difficulties and subscale latent trajectories memberships identified in optimal Growth Mixture Models, while also accounting for other sociodemographic covariates. We did not detect any multicollinearity among the variables using variance inflation factors.

The relative importance of each predictor was assessed using dominance analysis^64^. We employed the *misty*^65^ (v 0.6.8) package in R for this analysis, using a correlation matrix extracted from the fitted model via the *lavInspect* function from the *lavaan* (v 0.6-19) package^63^. This approach leverages the correlation matrix to consider not only individual predictors but also the correlations among them, providing a more comprehensive assessment of their relative importance^66^.

To examine potential causal pathways, mediation analyses were conducted, allowing sociodemographic factors to indirectly influence the age at diagnosis through their effects on SDQ latent trajectory memberships identified in the optimal Growth Mixture Model. Using structural equation modelling in the *lavaan* (v 0.6-19) package ^63^, both direct and indirect effects were assessed, with their significance calculated using bootstrapping analysis. Further details are provided in **Supplementary Note 4**.

To investigate the specificity of our findings to autism, we conducted Growth Mixture Models, Latent Growth Curve Models, regression, and mediation analyses in individuals with ADHD but without a co-occurring autism diagnosis in the MCS cohort (N = 89, **Supplementary Table 6,** results presented in **Supplementary Note 5**). ADHD diagnoses were available in the same sweeps as autism diagnoses, reported at ages 5, 7, 11, and 14. Carers were asked the following question: ‘Has a doctor or other health professional ever told you that <child’s name> had Attention Deficit Hyperactivity Disorder (ADHD)?’.

#### Imputation

To assess the impact of missingness, we applied *softImpute* (v.1.4-1), to impute missing data for all children with an autism or ADHD diagnosis reported by any carer in any sweep in the MCS cohort (Autism: N = 623, **Supplementary Table 5**; ADHD: N = 325, **Supplementary Table 6**). *SoftImpute* was chosen for its computational efficiency in handling large-scale matrices through low-rank approximation, effectively preserving underlying structure of input data. Further information is provided in **Supplementary Note 2**.

### SPARK cohort: Genotyping, quality control and imputation

We used data from the SPARK cohort^39^ iWES2 v1 dataset (released in Feb 2022) which included data from 70,487 autistic individuals and their families as the SPARK Discovery cohort. Data from SPARK iWES v3 (released August 2024) which included an additional 71,267 autistic individuals and their families was included in the SPARK Replication cohort. All participants in the Discovery cohort were genotyped using the Illumina Global Screening Array (GSA_24v2-0_A2) and in the Replication cohort using the Twist Bioscience genome-wide SNP capture panel for genotyping-by-sequencing. To avoid false positives due to fine-scale population stratification, we restricted the analyses to individuals of genetically-inferred European ancestries (Discovery: N = 51,869 and Replication: N= 50,211, autistic and non-autistic participants), which was provided by the SPARK consortium. From this, we excluded individuals with genotyping rate < 98%, individuals with sex mismatches, and those with excess heterozygosity (3 standard deviations from the mean heterozygosity). Where trio data was available, trios with greater than 5% Mendelian errors were excluded, resulting in 47,170 (Discovery) and 48,750 (Replication) autistic and non-autistic individuals. We included genetic variants with minor allele frequency [MAF] > 1%, genotyping rate > 95%, and that were in Hardy Weinberg Equilibrium (HWE P > 1×10^-6^), resulting in 518,189 (Discovery) and 1,225,308 (Replication) SNPs.

We used this quality controlled genotype data for imputation, calculating genetic principal components, and inferring relatedness among individuals. We inferred genetic relatedness using KING^67^ (v.2.3.2). For genetic principal component analysis, we pruned SNPs for linkage disequilibrium (LD) (maximum r^2^ = 0.1) and removed the human leukocyte antigen (HLA) region. Using PC-AiR^68^ in GENESIS (v2.22.2), we first calculated principal components (PCs) in genetically unrelated individuals and then projected the PCs onto related individuals. We imputed genotypes using the TOPMED imputation panel^69^ on the Michigan imputation server (v1.7.3)^70^ using Minimac4^70^ and after phasing using Eagle v2.5^71^. Post imputation, variants were converted from GRCh38/hg38 to GRCh37/hg19 using liftOver. We restricted downstream analyses only to variants with MAF > 0.1% and with an imputation R^2^ > 0.6.

### SPARK cohort: Association analyses

#### PGS association analyses

Polygenic scores (PGS) were calculated using PRScs^72^ (v. 1.1.0) which uses a Bayesian shrinkage prior. PGS were calculated for autism diagnosed before age 11 (iPSYCH_before11_), and autism diagnosed after age 10 (iPSYCH_after10_), generated using the iPSYCH2015^73^ cohort, details of which are provided below. For simplicity we refer to this cohort as iPSYCH throughout. PGS were calculated and analysed separately for the Discovery and Replication cohorts and meta-analysed using inverse-variance weighted meta-analysis^74^.

We ran separate linear regression analyses between each of the two PGS and age at autism diagnosis (converted to years in all analyses) in the quality controlled dataset. We excluded individuals older than 22 to focus on those who had an autism diagnosis using either the DSM-IV^75^ or DSM-5, retaining a maximum of 18,809 (Discovery cohort) and 9,383 (Replication cohort) autistic individuals for PGS analyses. This criteria also allowed us to focus on individuals who received their diagnosis in childhood or adolescence, as older adults may have missed an earlier diagnosis of autism due to secular changes in societal attitudes towards autism. The baseline model included ID (16.34% had caregiver reported ID), sex, and the first 10 genetic principal components as covariates. We ran eight different sensitivity analyses by including various covariates in addition to the covariates included in the baseline model. First we ran three models to account for developmental and clinical covariates: age at walking and age at first words (Model 2), age at walking and first words, autism severity (SCQ and RBS-R total scores), caregiver reported IQ scores, and language or other regression (Model 3), stratified analyses restricted to individuals without ID who can speak in longer sentences (Model 4). Next we ran two models accounting for sociodemographic factors: parental SES (Model 5) and, additionally, area deprivation (Model 6). We controlled for any attentional and behavioural diagnosis (Model 7), for DSM edition (DSM 4 vs DSM 5, Model 8), and trio status (Model 9). Finally, we also ran sensitivity analyses after stratifying by sex.

In the SPARK cohort, we obtained data for age at achieving nine developmental milestones (in months) for autistic individuals. For all milestones, we excluded individuals who were greater than five median absolute deviations from the median. We ran multiple linear regression with PGS for iPSYCH_before11_ and iPSYCH_after10_ GWASs with sex and the first 10 genetic principal components as covariates. Yet again, we ran the analyses for both the Discovery and Replication cohorts and meta-analysed it using inverse-variance weighted meta-analysis.

#### Rare high-impact de novo variants and inherited variants

We identified rare (MAF < 0.1%) *de novo* and inherited variants in complete trios from SPARK as previously described^40^. We identified high impact protein truncating variants by restricting to variants in loss-of-function observed/expected upper bound fraction (LOEUF)^76^ highly constrained decile (LOEUF < 0.37) that were annotated as either frameshift, stop gained, or start lost; and had a loss-of-function transcript effect estimator (LOFTEE) “high confidence annotation”. To identify high-impact *de novo* missense variants, we restricted to variants in LOEUF highly constrained genes (LOEUF < 0.37), and had an MPC (missense badness, PolyPhen-2, and constraint) score^77^ > 2.

We ran regression analyses for (1) high-impact *de novo* and inherited protein truncating variants, (2) missense variants, and (3) by combining both protein truncating and missense variants. We included sex and the first 10 genetic principal components as covariates.

### Genome-wide association studies

#### GWAS of age at autism diagnosis

We generated a GWAS of age at autism diagnosis (in years) in the quality controlled dataset from SPARK, restricting it to autistic individuals who were under 22 years of age (Discovery: N = 18,809 and Replication: N = 9,356), and SNPs with a MAF > 1%. GWAS was generated using FastGWA^78^ with sex, ID, and the first 10 genetic principal components as covariates. We meta-analysed the GWAS from the SPARK Discovery and Replication cohorts using inverse-variance weighted meta-analysis in Plink^79,80^ (2.0). In iPSYCH, we generated an additional GWAS of age at autism diagnosis (in years) in a quality controlled dataset of unrelated individuals with sex and ID included as covariates using FastGWA^78^, restricting to SNPs with an MAF > 1%. To keep it consistent with SPARK, we excluded individuals who were diagnosed after age 22, leaving a total sample of 18,965 individuals. Briefly, pre-imputation quality control of the iPSYCH data was performed using the Ricopili pipeline^81^, prephased using Eagle v.2.3.5, and imputed using Minimac3^82^, using the downloadable version of the Haplotype Reference Consortium (HRC)^83^ (accession no. EGAD00001002729). Further details of quality control and imputation are provided in Als et al., 2023^84^.

#### GWAS of autism stratified by age at diagnosis

We generated three age at autism diagnosis stratified GWAS in SPARK using (unscreened) non-autistic parents and siblings as controls (Discovery: N_control_ = 24,965 and Replication: N_control_ = 33,302). The three GWAS were: (1) SPARK, diagnosed before age 6 (SPARK_before6_; Discovery: N_autistic_ = 14,578 and Replication: N_autistic_ = 6,857); (2) SPARK, diagnosed before age 11 (SPARK_before11_, N_autistic_ = 18,719 and Replication: N_autistic_ = 9,162); and (3) SPARK, diagnosed after age 10 (SPARK_after10_, N_autistic_ = 3,358 and Replication: N_autistic_ = 2,885). For these analyses, we did not restrict it to individuals under 22 to increase sample size. Of note, SPARK_before11_ overlaps with the SPARK_before6_ cohort. GWAS were generated using quality controlled SNPs with a MAF > 1% using FastGWA-GLMM^85^. We included age at recruitment into the study (to account for the use of parents as controls, who potentially lack an autism diagnosis due to secular changes in attitudes and diagnosis), sex and the first 10 genetic principal components as covariates. Fast-GWA GLMM can account for relatedness and fine-scale population stratification even in family-based samples like SPARK. Given the relatively low sample size of the Replication cohort we meta-analysis the Replication and Discovery cohort GWAS using inverse-variance weighted meta-analyses in Plink^79,80^ (v2.0).

Although inclusion of unscreened related individuals as controls can decrease heritability and statistical power to identify loci^86^, we used the GWAS to primarily conduct genetic correlation and related analyses. To ensure the robustness of these models we: (1) confirmed that the attenuation ratio for all GWAS was not significantly greater than 1; (2) generated an additional GWAS of SPARK without stratifying by age at autism diagnosis using the same methods and confirmed a high genetic correlation (r_g_ = 0.92, SE = 0.17) with a previous SPARK GWAS^87^ which used a case-pseudocontrol approach; and (3) in the genomicSEM analyses, ran sensitivity analyses using a trio-based SPARK GWAS^87^ in lieu of the age at diagnosis stratified GWAS from SPARK and confirmed our findings.

We generated three GWAS of autism stratified by age at autism diagnosis in the iPSYCH cohort^73^. The primary GWAS used in the analyses were GWAS of autism diagnosed before age 11 (iPSYCH_before11_: 9,500 autistic and 36,667 non-autistic individuals) and autism diagnosed after age 10 (iPSYCH_after10_: 9,231 autistic and 36,667 non-autistic individuals). We chose to subdivide the iPSYCH cohort at age 10 because we observed an increase in SDQ scores in birth cohorts at this age, and because age 10 is associated with an increase in diagnosis of females in epidemiological samples^34^. Additionally, we conducted a GWAS of autism diagnosed before age nine (iPSYCH_before9_: 5,451 autistic and 36,667 non-autistic individuals).

All individuals included in these GWAS from iPSYCH were born between May 1980 and December 2008 to mothers who were living in Denmark. GWAS was conducted on unrelated individuals of European ancestry, with the first 10 genetic principal components included as covariates using logistic regression as provided in PLINK.

### Heritability, genetic correlation, and genomicSEM

Heritability analyses for age at autism diagnosis were conducted using a single-component genome-wide complex trait analysis with genomic-relatedness-based restricted maximum likelihood approach (GCTA-GREML v1.94.1)^88,89^ in unrelated autistic individuals using the quality controlled genetic data in SPARK. We estimated SNP-based heritability first after including sex and the first ten genetic principal components as covariates, and then additionally with ID as a covariate (baseline model). We ran several sensitivity analyses after including additional covariates: (1) developmental milestones (age at first words and age at walking); (2) developmental milestones and developmental regression (language regression and other regression); (3) developmental milestones, developmental regression, and IQ scores; (4) SCQ and RBS-R scores; (5) SCQ and RBS-R scores, developmental milestones, and developmental regression; (6) developmental milestones and SES; and (7) developmental milestones, SES, and deprivation.

We conducted genetic correlation analyses using LDSC, using linkage disequilibrium scores from the north-west European populations.

We conducted genetic correlation analyses among different autism GWAS using LDSC (v.1.0.1). This included a European-only case-pseudocontrol GWAS in SPARK^87^ (4,535 case-pseudocontrol pairs); GWAS from FinnGen (Data Release - r10)^90^ (646 cases and 301,879 controls), the PGC-2017 autism GWAS^91^ (7,387 cases and 8,567 controls), GWAS from iPSYCH, and age at diagnosis stratified GWAS from SPARK. The iPSYCH GWAS included an unstratified (19,870 autistic individuals [15,025 males and 4,845 females] and 39,078 controls) and sex-stratified GWAS^92^, and three age at diagnosis stratified GWAS as mentioned earlier.

For genomicSEM^42^ (v.0.0.5) analyses, we restricted to six GWAS with minimal sample overlap, without high genetic correlation (r_g_ > 0.95), and with wide variation in age at diagnosis to conduct genomicSEM analyses using autosomes. Using the patterns of genetic correlations observed we tested an age at diagnosis related correlated two-factor model. We additionally tested: (1) a single-factor model; (2) a correlated two-factor “geography” model where three US-based autism GWAS loaded onto one factor, and three Europe-based autism GWAS loaded onto a second factor; (3) a bifactor model based on age at diagnosis; (4) a bifactor model based on the geography of the cohorts; and (5) a hierarchical factor model based on age at diagnosis. The two-factor model was chosen as it had lower RMSEA and higher CFI and was more parsimonious than the bifactor model. We ran sensitivity analyses using different GWAS of autism as input and confirmed that the two-correlated-factor model was the best fitting model of the models tested.

### Analyses in ALSPAC and MCS

#### Genetic quality control for ALSPAC

We obtained quality controlled and imputed genotype data from ALSPAC^93–95^. Further details about the cohort are provided in the **Supplementary Note 1.** Briefly, ALSPAC children were genotyped using the Illumina HumanHap550 quad chip genotyping platforms by 23andme. Individuals were excluded due to sex mismatches, excess heterozygosity, missingness > 3%, and insufficient sample replication (Identical-By-Descent [IBD] < 0.8). After multidimensional scaling, and comparison with Hapmap II (release 22), only individuals of genetically inferred European ancestries were retained. SNPs with low frequency (MAF < 1%), poor genotyping (call rate < 95%) and deviations from Hardy-Weinberg equilibrium (P < 5×10^-7^) were removed. 9,115 subjects and 500,527 SNPs passed quality control. Genotypes were phased using ShapeIT, and imputation was done using the Haplotype Reference Consortium panel using the Michigan imputation server. After imputation, we further removed low frequency SNPs (MAF < 1%). Further details of the quality control and imputation of ALSPAC are provided here: https://proposals.epi.bristol.ac.uk/alspac_omics_data_catalogue.html#org89bb79b. Genome-wide genotype data was generated by Sample Logistics and Genotyping Facilities at Wellcome Sanger Institute and LabCorp (Laboratory Corporation of America) using support from 23andMe.

#### Genetic quality control for MCS

We also obtained quality controlled and imputed data from MCS. Briefly, MCS samples were genotyped using the Illumina Global Screening Array^96^. Individuals were excluded due to sex mismatches, excess heterozygosity, and missingness > 2%. We identified European samples using the GenoPred pipeline^97^ (https://github.com/opain/GenoPred). SNPs with low frequency (MAF < 1%), poor genotyping (call rate < 97%) and deviations from Hardy-Weinberg equilibrium (P < 1×10^-6^) were removed. Imputation was conducted using Minimac4^70^ using the TOPMED reference panel^69^ in the Michigan imputation server^70^. Post imputation, SNPs with an imputation R^2^ INFO score < 0.8, with > 3% missing, and with a MAF < 1% were excluded. Further details are available here: https://cls-genetics.github.io/docs/MCS.html

#### PGS association with SDQ

PGS for both ALSPAC and MCS were calculated in individuals of genetically inferred European ancestries. Genetic principal components were calculated for both cohorts using PC-AiR as described earlier. We calculated PGS for iPSYCH_before11_ and iPSYCH_after10_ and used these in all analyses in the MCS to keep it consistent with analyses in SPARK.

We obtained scores on the SDQ total and subscales for six ages in the MCS and five ages in ALSPAC. We ran cross-sectional analysis at each age using multiple linear regression with PGS for iPSYCH_before11_ and iPSYCH_after10,_ with sex, age, and the first 10 genetic principal components as covariates. Additionally, we ran multiple linear mixed effects regression using *lme4* (v.1.1.27.1) package in R^98^, fitting a PGS by age interaction term to investigate if the effects of PGS on SDQ change over time.

To investigate if the differences in association between MCS and ALSPAC were due to differences in ascertainment between the two cohorts, we matched ALSPAC to MCS using entropy balancing^99^ and re-ran the PGS association analyses. Entropy balancing is a reweighting technique that ensures the covariate distributions are identical between groups. This method uses optimisation algorithms to assign weights to individuals such that the weighted average of the covariates in ALSPAC (the larger genotyped cohort) matches that of MCS (the smaller genotyped cohort), minimising confounding biases and increasing comparability. We used the child’s biological sex, maternal age at delivery, and maternal highest educational qualification at first data collection in each cohort as matching factors. Entropy balancing was conducted using the *ebal* (v 0.1-8) package in R^100^.

#### PGS association with social communication skills and autism diagnosis

In ALSPAC, we obtained understanding of simple phrases (e.g., “do you want that”, or “come here”) and gesture scores from the Macarthur-Bates Communicative Development Inventories^101^ at 15 months of age (**Supplementary Note 9**). We conducted multiple linear regression using PGS for iPSYCH_before11_ and iPSYCH_after10_, with sex, age, and the first 10 genetic principal components as covariates.

Autism diagnosis in the MCS was obtained using parent/caregiver reports of autism/asperger syndrome diagnosis by a doctor at ages 5, 7, 11, and 14. We identified individuals with an autism diagnosis at age 7 or earlier, age 11 or earlier, or between ages 11 and 14. We conducted Firth’s bias-reduced multiple logistic regression (*logistf* (v 1.26.0) package in R) using PGS for iPSYCH_before11_ and iPSYCH_after10_, with sex, age and the first 10 genetic principal components covariates.

## Ethics

Ethical approval for individual cohorts were obtained independent of the current study. Ethical approval for ALSPAC was obtained from the ALSPAC Ethics and Law Committee and the Local Research Ethics Committees. Ethical approval for each sweep of MCS was obtained from NHS Research Ethics Committees (MREC). Ethical approval for LSAC was obtained from the Australian Institute of Family Studies Human Research Ethics Committee. Ethical approval for GUI was obtained from a dedicated Research Ethics Committee set up by the Department of Children, Equality, Disability, Integration and Youth. Ethical approval for SPARK was obtained from the Western Institutional Review Board-Copernicus Group IRB Protocol #20151664). The Danish Scientific Ethics Committee, the Danish Health Data Authority, the Danish data protection agency and the Danish Neonatal Screening Biobank Steering Committee approved the iPSYCH study. Ethical approval for the analyses of de-identified data used in this study was obtained from the Cambridge Human Biology Research Ethics Committee (HBREC.2020.07).

## Code availability

- Lavaan (LGCM) (v.0.6-19) : https://lavaan.ugent.be/tutorial/growth.html
- lcmm(GMM), v 2.2.1: https://github.com/CecileProust-Lima/lcmm
- Softimpute (v.1.4-1): https://cran.r-project.org/web/packages/softImpute/softImpute.pdf
- Misty (v 0.6.8) : https://cran.r-project.org/web/packages/misty/index.html
- PRScs (v.1.1.0): https://github.com/getian107/PRScs
- fastGWA and GCTA (v.1.94.1): https://yanglab.westlake.edu.cn/software/gcta/#Overview
- GenomicSEM (v.0.0.5): https://github.com/GenomicSEM/GenomicSEM
- LDSC (v1.0.1): https://github.com/bulik/ldsc
- KING (v.2.3.2): https://www.kingrelatedness.com/manual.shtml
- Plink 2.0: https://www.cog-genomics.org/plink/2.0/
- GENESIS (v2.22.2): https://github.com/UW-GAC/GENESIS
- Lme4 (v.1.1.27.1): https://github.com/lme4/lme4/
- Logistf (v.1.24): https://cran.r-project.org/web/packages/logistf/index.html
- Ebal (v 0.1-8): https://cran.r-project.org/web/packages/ebal/ebal.pdf
- Relaimpo (v2.2-7): https://cran.r-project.org/web/packages/relaimpo/relaimpo.pdf
- SPARK quality control, imputation and GWAS: https://github.com/vwarrier/SPARK_iWES2_imputation/
- Bespoke genetic analyses code: https://github.com/vwarrier/autism_agediagnosis/

## Data availability

- SPARK autism GWAS: https://bitbucket.org/steinlabunc/spark_asd_sumstats/src
- Finngen autism GWAS: https://www.finngen.fi/en/access_results
- iPSYCH autism GWAS (unstratified, sex-stratified and age at diagnosis stratified, age at diagnosis) can be obtained from Anders Borglum and Jakob Grove.
- Psychiatric GWAS summary stats: https://pgc.unc.edu/for-researchers/download-results/
- GWAS educational attainment: https://thessgac.com/papers/
- GWAS cognitive aptitude: https://cncr.nl/research/summary_statistics/
- For ALSPAC, the study website contains details of all the data that is available through a fully searchable data dictionary and variable search tool": http://www.bristol.ac.uk/alspac/researchers/our-data/
- For MCS, data can be obtain after application through the UK Data Service: https://beta.ukdataservice.ac.uk/datacatalogue/series/series?id=2000031
- Summary statistics for the SPARK-based age at diagnosis GWAS, and the age at diagnosis stratified GWAS generated from the genomicSEM models will be made available upon publication.

## Acknowledgements

This research was supported by funding from the Simons Foundation for Autism Research Initiative, the Wellcome Trust (214322\Z\18\Z and 226392/Z/22/Z), Horizon-Europe R2D2-MH (grant agreement number 101057385), and UKRI (10063472). For the purpose of open access, we have applied a CC BY public copyright licence to any author-accepted manuscript version arising from this submission. S.B.-C. also received funding from the Autism Centre of Excellence at Cambridge, the Templeton World Charitable Fund, the MRC and the National Institute for Health Research Cambridge Biomedical Research Centre. The research was supported by the National Institute for Health Research Applied Research Collaboration East of England. Any views expressed are those of the author(s) and not necessarily those of the funder. Some of the results leading to this publication have received funding from the Innovative Medicines Initiative 2 Joint Undertaking under grant agreement no. 777394 for the project AIMS-2-TRIALS. This joint undertaking receives support from the European Union’s Horizon 2020 research and innovation program and the EFPIA and Autism Speaks, Autistica and the SFARI. The iPSYCH team was supported by grants from the Lundbeck Foundation (R102-A9118, R155-2014-1724 and R248-2017-2003), the NIMH (1R01MH124851-01 to A.D.B.), and EU’s Horizon Europe program (R2D2-MH; grant agreement no. 101057385 to A.D.B.). The Danish National Biobank resource was supported by the Novo Nordisk Foundation. High-performance computer capacity for handling and statistical analysis of iPSYCH data on the GenomeDK HPC facility was provided by the Center for Genomics and Personalized Medicine and the Centre for Integrative Sequencing, iSEQ, Aarhus University, Denmark (grant to A.D.B.). The UK Medical Research Council and Wellcome (Grant ref: 217065/Z/19/Z) and the University of Bristol provide core support for ALSPAC. This publication is the work of the authors and the authors will serve as guarantors for the contents of this paper. A comprehensive list of grants funding is available on the ALSPAC website (http://www.bristol.ac.uk/alspac/external/documents/grant-acknowledgements.pdf). R2D2-MH has been funded by Horizon Europe [grant agreement no. 101057385], by UK Research and Innovation (UKRI) under the UK government’s Horizon Europe funding guarantee [grant no.10039383] and by the Swiss State Secretariat for Education, Research and Innovation (SERI) under contract number 22.00277. A.H. and L.E.H. were supported by the Norwegian Research Council (#336085) and the Norwegian Health Authority (#2020022; #2022029; #2022083). EV and BSTP are funded by the Max Planck Society.

We are grateful to the Centre for Longitudinal Studies (CLS), UCL Social Research Institute, for the use of these data and to the UK Data Service for making them available. However, neither CLS nor the UK Data Service bear any responsibility for the analysis or interpretation of these data. This paper uses unit record data from Growing Up in Australia, the Longitudinal Study of Australian Children. The study is conducted in partnership between the Department of Social Services (DSS), the Australian Institute of Family Studies (AIFS) and the Australian Bureau of Statistics (ABS). The findings and views reported in this paper are those of the author and should not be attributed to DSS, AIFS or the ABS. Growing Up in Ireland (GUI) has been funded by the Government of Ireland through the Department of Children, Equality, Disability, Integration and Youth (DCEDIY) and the Central Statistics Office (CSO). Results in this report are based on analysis of data from Research Microdata Files provided by the Central Statistics Office (CSO). Neither the CSO nor the DCEDIY take any responsibility for the views expressed or the outputs generated from these analyses. We are extremely grateful to all the families who took part in this study, the midwives for their help in recruiting them, and the whole ALSPAC team, which includes interviewers, computer and laboratory technicians, clerical workers, research scientists, volunteers, managers, receptionists and nurses. This study includes data from the Norwegian Mother, Father and Child Cohort Study (MoBa), conducted by the Norwegian Institute of Public Health. We are grateful to all the participating families.

We thank Alex Kwong, Tamsin Ford, Will Mandy, and Andrew Grotzinger for helpful discussions.

## Ethics declarations

ADB received speakers’ fee from Lundbeck. The authors declare no competing interests.

## Author contributions

XZ and VW conducted most of the analyses, with the remainder being conducted by JG, YG, CKB, and LKN. MK, EV, AG, LH, AH, AR, BSt.P, and ADB provided summary statistics for various analyses. SASN, DSM, and EMW carried out data preparation and quality control, with assistance from KES, VKC, PD, SL, TM, MK, SA and DB. VW supervised the analyses and directed the study with inputs from HCM and JG. VW and XZ wrote the initial draft with input from HCM. AG, SASN, AH, BSt.B, AR, DSM, EW, DHG, NRW, EBR, TB, and SBC provided intellectual input. All authors read and commented on the final manuscript.

## APEX Consortium

Deep Adhya, Carrie Allison, Bonnie Ayeung, Rosie Bamford, Simon Baron-Cohen, Richard Bethlehem, Tal Biron-Shental, Graham Burton, Wendy Cowell, Jonathan Davies, Joanna Davis, Dori Floris, Alice Franklin, Lidia Gabis, Daniel Geschwind, Ramin Ali Marandi Ghoddousi, David M. Greenberg, Yuanjun Gu, Alexandra Havdahl, Alexander Heazell, Rosemary Holt, Matthew Hurles, Yumnah Khan, Meng-Chuan Lai, Madeline Lancaster, Michael Lombardo, Hilary Martin, Jose Gonzalez Martinez, Jonathan Mill, Mahmoud Koko Musa, Kathy Niakan, Adam Pavlinek, Lucia Dutan Polit, Marcin Radecki, David Rowitch, Jenifer Sakai, Laura Sichlinger, Deepak Srivastava, Alexandros Tsompanidis, Florina Uzefovsky, Varun Warrier, Elizabeth Weir, Xinhe Zhang.

## iPSYCH Autism working group

Anders Borglum, Jonas Bybjerg-Grauholm, Jakob Grove, David M. Hougaard, Ole Mors, Preben Bo Mortensen, Merete Nordentoft and Thomas Werge.

## PGC-PTSD consortium

Caroline M. Nievergelt, Adam X. Maihofer, Elizabeth G. Atkinson, Chia-Yen Chen, Karmel W. Choi, Jonathan R. I. Coleman, Nikolaos P. Daskalakis, Laramie E. Duncan, Renato Polimanti, Cindy Aaronson, Ananda B. Amstadter, Soren B. Andersen, Ole A. Andreassen, Paul A. Arbisi, Allison E. Ashley-Koch, S. Bryn Austin, Esmina Avdibegoviç, Dragan Babić, Silviu-Alin Bacanu, Dewleen G. Baker, Anthony Batzler, Jean C. Beckham, Sintia Belangero, Corina Benjet, Carisa Bergner, Linda M. Bierer, Joanna M. Biernacka, Laura J. Bierut, Jonathan I. Bisson, Marco P. Boks, Elizabeth A. Bolger, Amber Brandolino, Gerome Breen, Rodrigo Affonseca Bressan, Richard A. Bryant, Angela C. Bustamante, Jonas Bybjerg-Grauholm, Marie Bækvad-Hansen, Anders D. Børglum, Sigrid Børte, Leah Cahn, Joseph R. Calabrese, Jose Miguel Caldas-de-Almeida, Chris Chatzinakos, Sheraz Cheema, Sean A. P. Clouston, Lucía Colodro-Conde, Brandon J. Coombes, Carlos S. Cruz-Fuentes, Anders M. Dale, Shareefa Dalvie, Lea K. Davis, Jürgen Deckert, Douglas L. Delahanty, Michelle F. Dennis, Frank Desarnaud, Christopher P. DiPietro, Seth G. Disner, Anna R. Docherty, Katharina Domschke, Grete Dyb, Alma Džubur Kulenović, Howard J. Edenberg, Alexandra Evans, Chiara Fabbri, Negar Fani, Lindsay A. Farrer, Adriana Feder, Norah C. Feeny, Janine D. Flory, David Forbes, Carol E. Franz, Sandro Galea, Melanie E. Garrett, Bizu Gelaye, Joel Gelernter, Elbert Geuze, Charles F. Gillespie, Slavina B. Goleva, Scott D. Gordon, Aferdita Goçi, Lana Ruvolo Grasser, Camila Guindalini, Magali Haas, Saskia Hagenaars, Michael A. Hauser, Andrew C. Heath, Sian M. J. Hemmings, Victor Hesselbrock, Ian B. Hickie, Kelleigh Hogan, David Michael Hougaard, Hailiang Huang, Laura M. Huckins, Kristian Hveem, Miro Jakovljević, Arash Javanbakht, Gregory D. Jenkins, Jessica Johnson, Ian Jones, Tanja Jovanovic, Karen-Inge Karstoft, Milissa L. Kaufman, James L. Kennedy, Ronald C. Kessler, Alaptagin Khan, Nathan A. Kimbrel, Anthony P. King, Nastassja Koen, Roman Kotov, Henry R. Kranzler, Kristi Krebs, William S. Kremen, Pei-Fen Kuan, Bruce R. Lawford, Lauren A. M. Lebois, Kelli Lehto, Daniel F. Levey, Catrin Lewis, Israel Liberzon, Sarah D. Linnstaedt, Mark W. Logue, Adriana Lori, Yi Lu, Benjamin J. Luft, Michelle K. Lupton, Jurjen J. Luykx, Iouri Makotkine, Jessica L. Maples-Keller, Shelby Marchese, Charles Marmar, Nicholas G. Martin, Gabriela A. Martínez-Levy, Kerrie McAloney, Alexander McFarlane, Katie A. McLaughlin, Samuel A. McLean, Sarah E. Medland, Divya Mehta, Jacquelyn Meyers, Vasiliki Michopoulos, Elizabeth A. Mikita, Lili Milani, William Milberg, Mark W. Miller, Rajendra A. Morey, Charles Phillip Morris, Ole Mors, Preben Bo Mortensen, Mary S. Mufford, Elliot C. Nelson, Merete Nordentoft, Sonya B. Norman, Nicole R. Nugent, Meaghan O’Donnell, Holly K. Orcutt, Pedro M. Pan, Matthew S. Panizzon, Gita A. Pathak, Edward S. Peters, Alan L. Peterson, Matthew Peverill, Robert H. Pietrzak, Melissa A. Polusny, Bernice Porjesz, Abigail Powers, Xue-Jun Qin, Andrew Ratanatharathorn, Victoria B. Risbrough, Andrea L. Roberts, Alex O. Rothbaum, Barbara O. Rothbaum, Peter Roy-Byrne, Kenneth J. Ruggiero, Ariane Rung, Heiko Runz, Bart P. F. Rutten, Stacey Saenz de Viteri, Giovanni Abrahão Salum, Laura Sampson, Sixto E. Sanchez, Marcos Santoro, Carina Seah, Soraya Seedat, Julia S. Seng, Andrey Shabalin, Christina M. Sheerin, Derrick Silove, Alicia K. Smith, Jordan W. Smoller, Scott R. Sponheim, Dan J. Stein, Synne Stensland, Jennifer S. Stevens, Jennifer A. Sumner, Martin H. Teicher, Wesley K. Thompson, Arun K. Tiwari, Edward Trapido, Monica Uddin, Robert J. Ursano, Unnur Valdimarsdóttir, Miranda Van Hooff, Eric Vermetten, Christiaan H. Vinkers, Joanne Voisey, Yunpeng Wang, Zhewu Wang, Monika Waszczuk, Heike Weber, Frank R. Wendt, Thomas Werge, Michelle A. Williams, Douglas E. Williamson, Bendik S. Winsvold, Sherry Winternitz, Christiane Wolf, Erika J. Wolf, Yan Xia, Ying Xiong, Rachel Yehuda, Keith A. Young, Ross McD Young, Clement C. Zai, Gwyneth C. Zai, Mark Zervas, Hongyu Zhao, Lori A. Zoellner, John-Anker Zwart, Terri deRoon-Cassini, Sanne J. H. van Rooij, Leigh L. van den Heuvel, AURORA Study, Estonian Biobank Research Team, FinnGen Investigators, HUNT All-In Psychiatry, Murray B. Stein, Kerry J. Ressler and Karestan C. Koenen

## Extended Data Figures

**Extended Data Figure 1:**
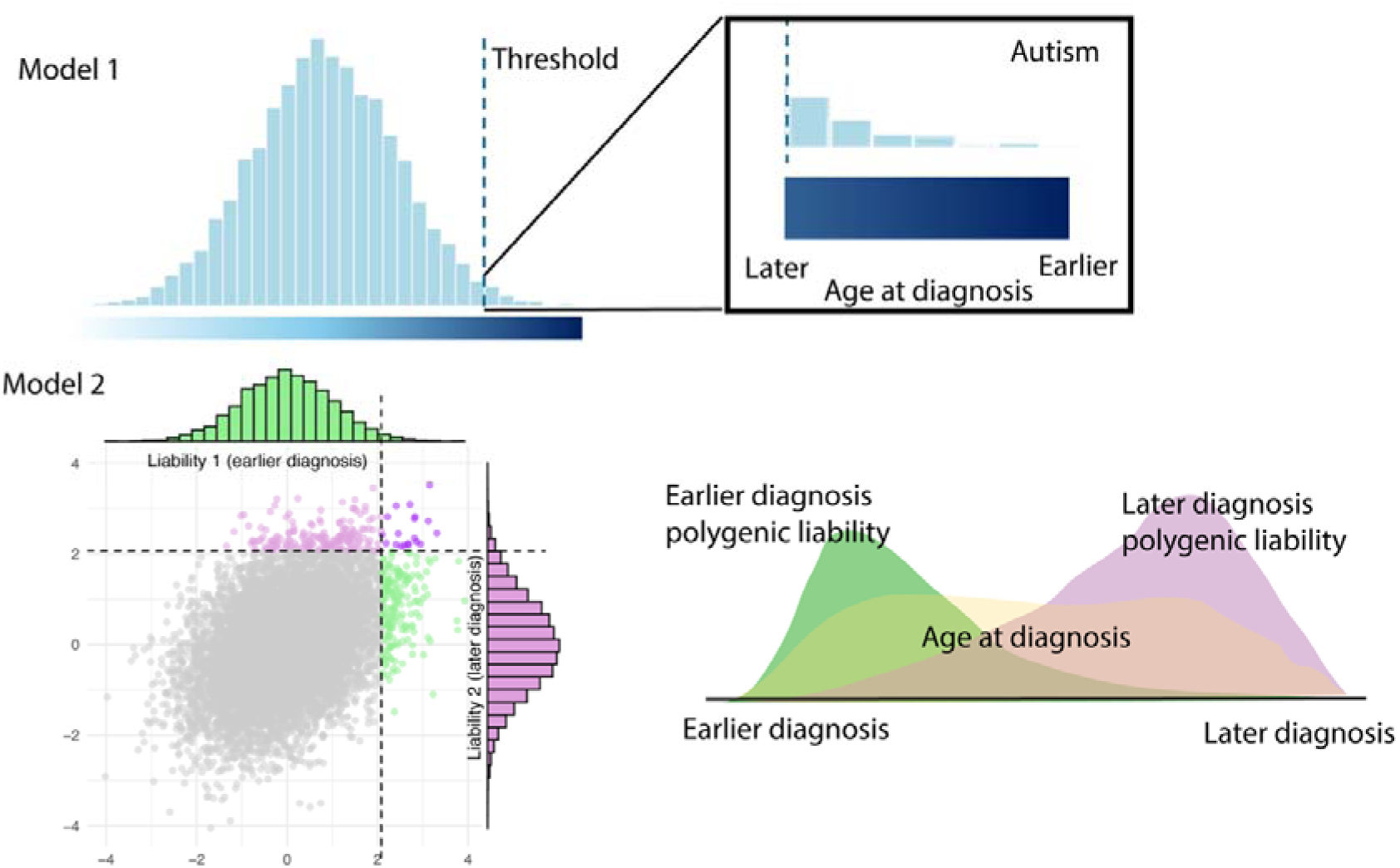
Variance in age at diagnosis explained by various clinical and sociodemographic factors. Variance explained (R² or η²) in age at autism diagnosis by clinical and sociodemographic factors, identified from the review of literature (1998-2023). Variables are grouped into sociodemographic (MAD, SES), socioemotional-behavioural (SDQ scores), sex, clinical (e.g., IQ, regression, language ability), and autism severity (e.g, SCQ, ADOS, ADI-R, RBS-R) categories. Circle size represents sample size, with larger circles indicating larger cohorts. Colored points denote variables analysed in the current study. Inset shows factors that explain greater than 10% of the variance in age at autism diagnosis. Note: None of these studies account for additional family, service access, and contextual factors known to influence diagnostic timing. Abbreviations: MAD, Maternal Age at Delivery; IQ, Intelligence Quotient; SES, Socio-economic Status; ADOS, Autism Diagnostic Observation Schedule; ADI-R, Autism Diagnostic Interview-Revised; RBS-R, Repetitive Behavior Scale-Revised; SCQ, Social Communication Questionnaire; SDQ, Strengths and Difficulties Questionnaire; SDQ Total, SDQ Total Difficulties.

**Extended Data Figure 2:**
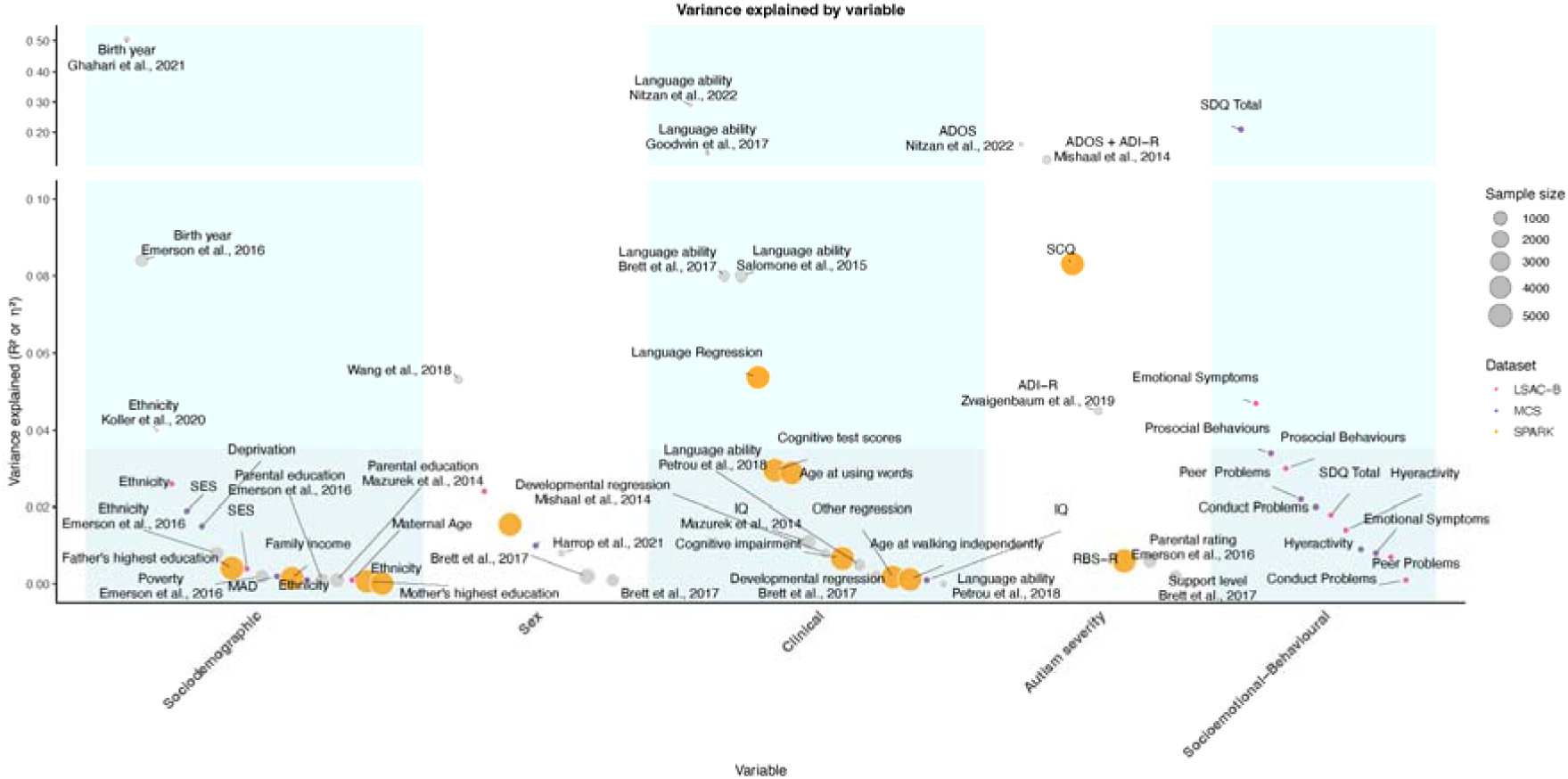
Two aetiological polygenic models of autism. In Model 1 (Unitary Model), we assume a single liability threshold polygenic model. In this model, autism emerges from a unitary polygenic aetiology. Autistic individuals diagnosed later have lower polygenic predisposition than individuals diagnosed earlier. In Model 2 (Developmental Model), we model two correlated age-dependent polygenic liabilities.

**Extended Data Figure 3:**
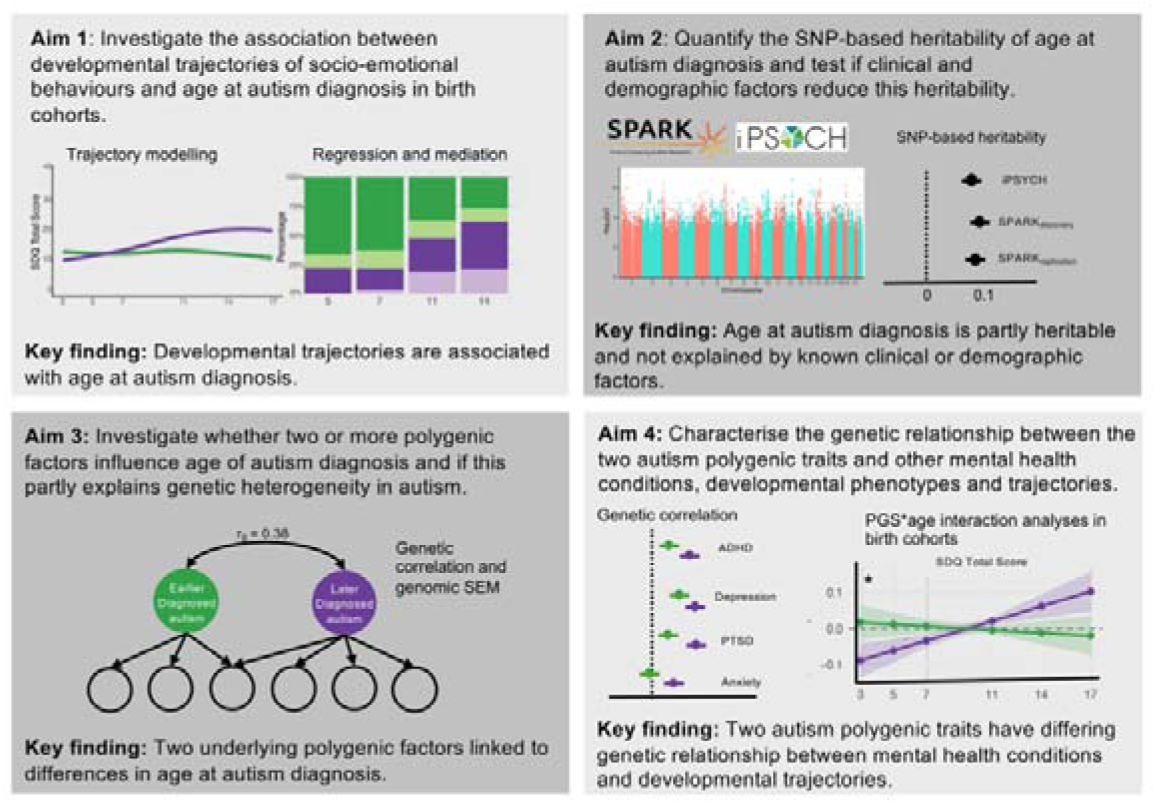
Schematic of the study aims. The study consists of four linked aims to understand whether the developmental trajectories and polygenic etiology of autism differs by age at diagnosis. In Aim 1, we modelled socioemotional and behavioural trajectories among autistic individuals in birth cohorts and investigated their association with age at autism diagnosis. In Aim 2, estimated the SNP-based heritability of age at autism diagnosis and whether it attenuates when accounting for various clinical and demographic factors. In Aim 3, we investigated whether the varying patterns of genetic correlations observed among different GWAS of autism can be explained by different polygenic factors associated with age at diagnosis. In Aim 4, we investigated the genetic relationship between the two autism polygenic factors and mental health and developmental phenotypes.

**Extended Data Figure 4:**
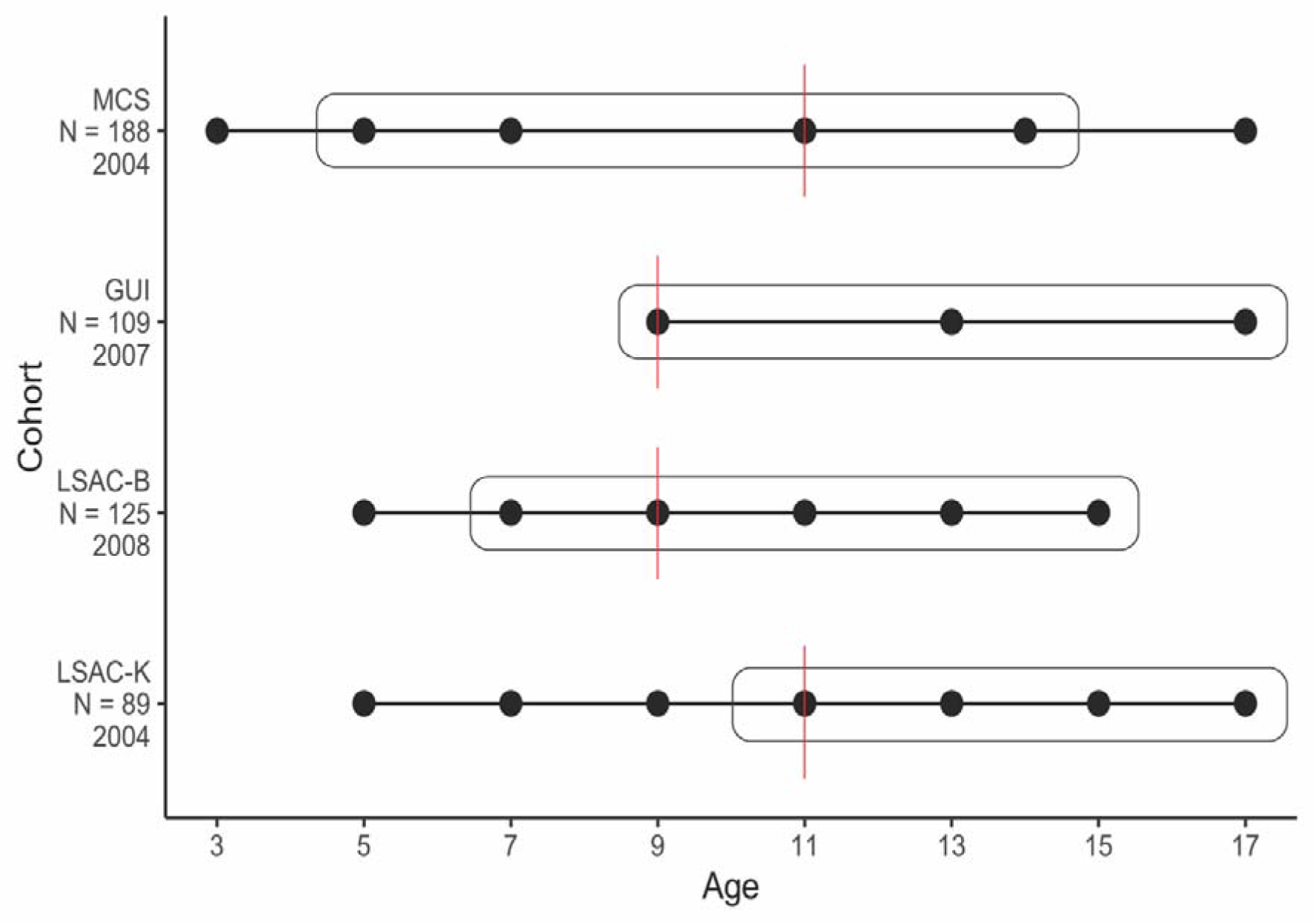
Schematic diagram of the birth cohorts included in the study. Schematic diagram of the cohorts included in the study and the ages when data was collected for SDQ scores (dots) and autism diagnosis (in boxes). Reports of autism diagnosis were available at ages: MCS - 5,7,11,14; GUI - 9,13,17; LSAC-B - 7,9,11,13,15; and LSAC-K: 11,13,15,17. MCS = Millennium Cohort Study; GUI = Growing up in Ireland (cohort ’98); LSAC-B = Longitudinal Study of Australian Children (Birth cohort); LSAC-K = Longitudinal Study of Australian Children (Kindergarten cohort). Sample sizes and the year of initial SDQ data collection for each cohort are shown on the ordinate axis. The age cutoff used in the Latent Growth Curve Models for each cohort is indicated by a red line. GUI was used only for Latent Growth Curve Models and excluded from Growth Mixture Models.

**Extended Data Figure 5:**
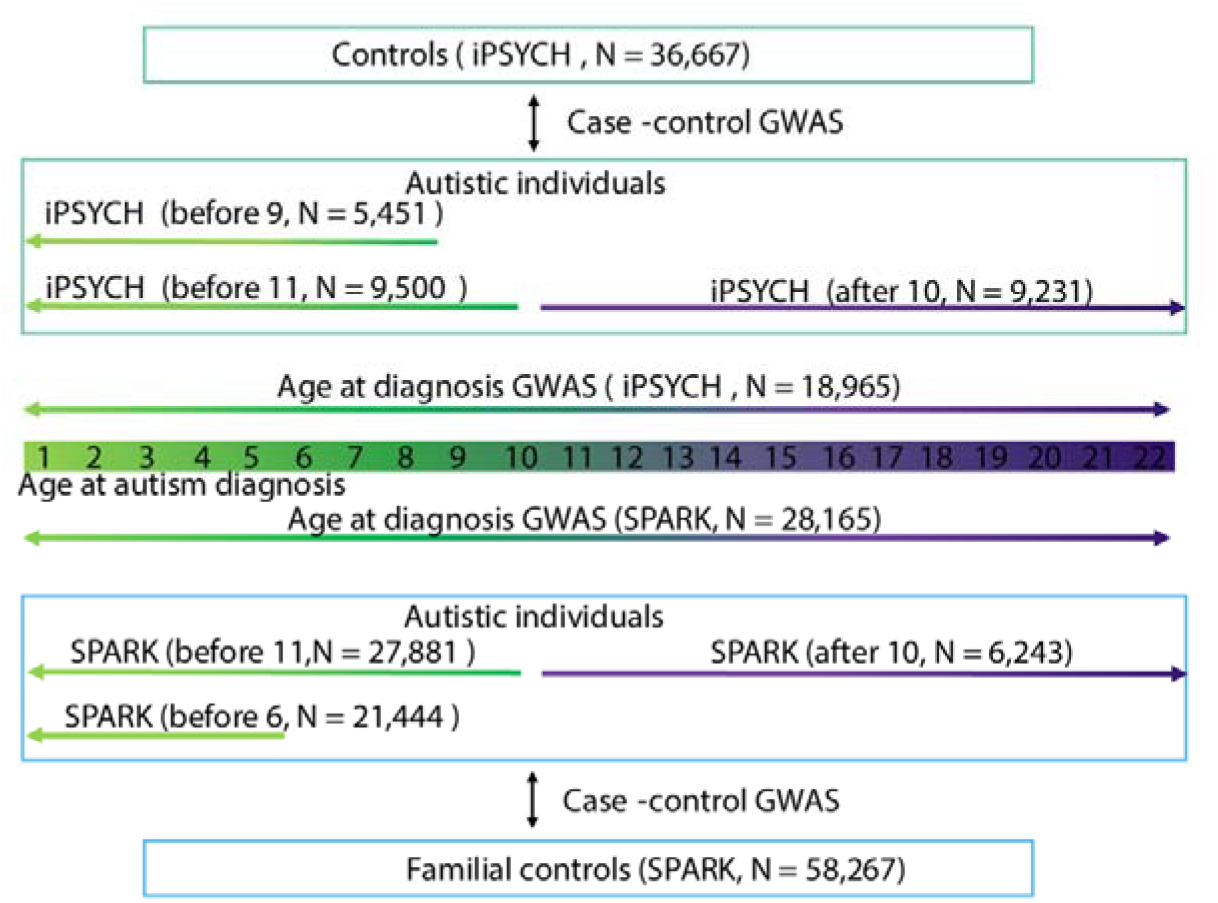
Schematic diagram of age at autism diagnosis GWAS and age stratified autism GWAS. Schematic diagram illustrating the main GWAS conducted in the study using the SPARK and iPSYCH cohorts. We conducted two age at autism diagnosis GWAS. In addition, we conducted six case-control GWAS, where autistic individuals were stratified based on their age at autism diagnosis.

**Extended Data Figure 6:**
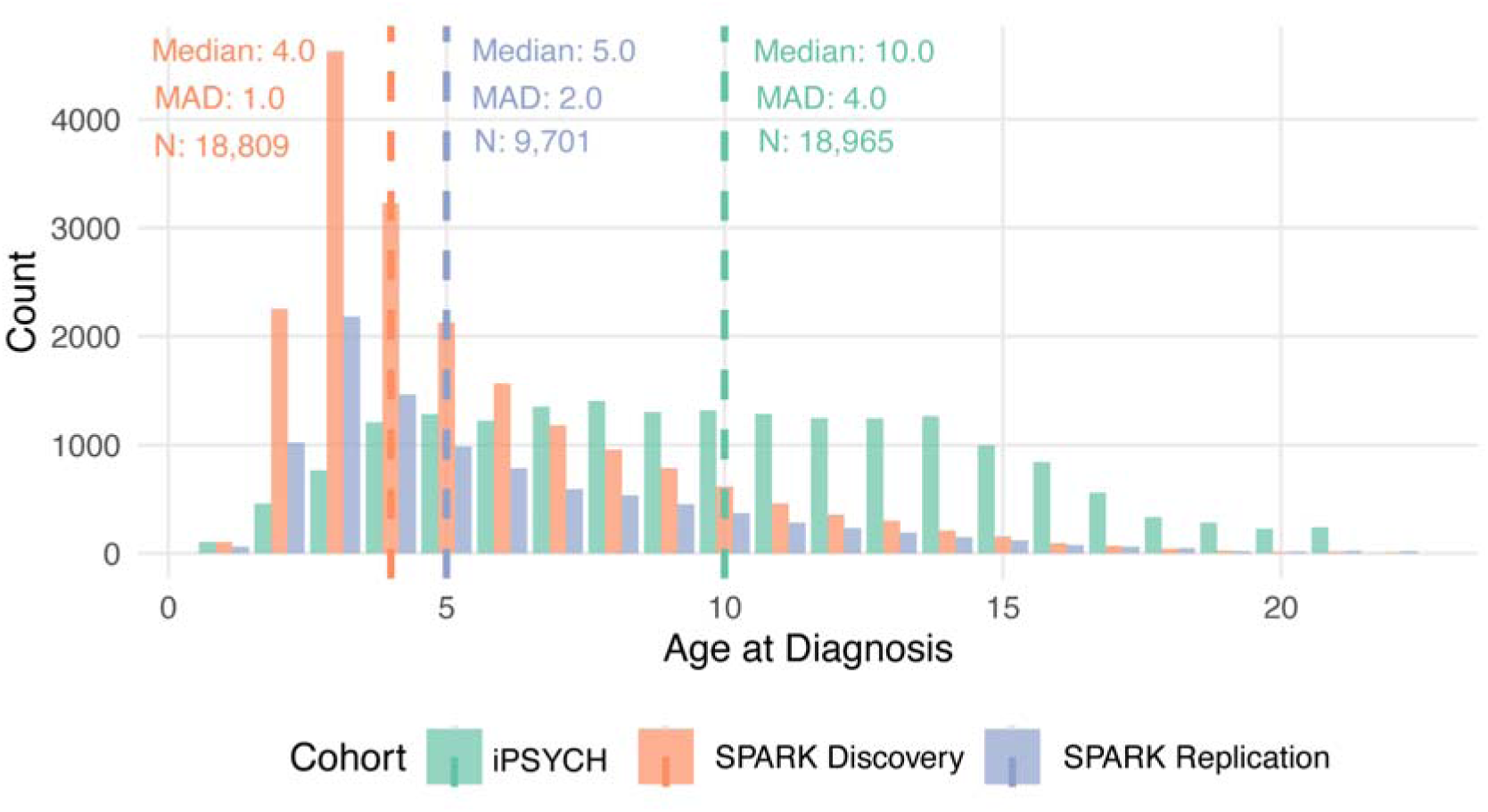
Distribution of age at autism diagnosis in iPSYCH and SPARK. Frequency histograms of age at autism diagnosis in iPSYCH and SPARK. Median and median absolute deviation (MAD) for age at diagnosis, and sample sizes (N) have been provided.

**Extended Data Figure 7:**
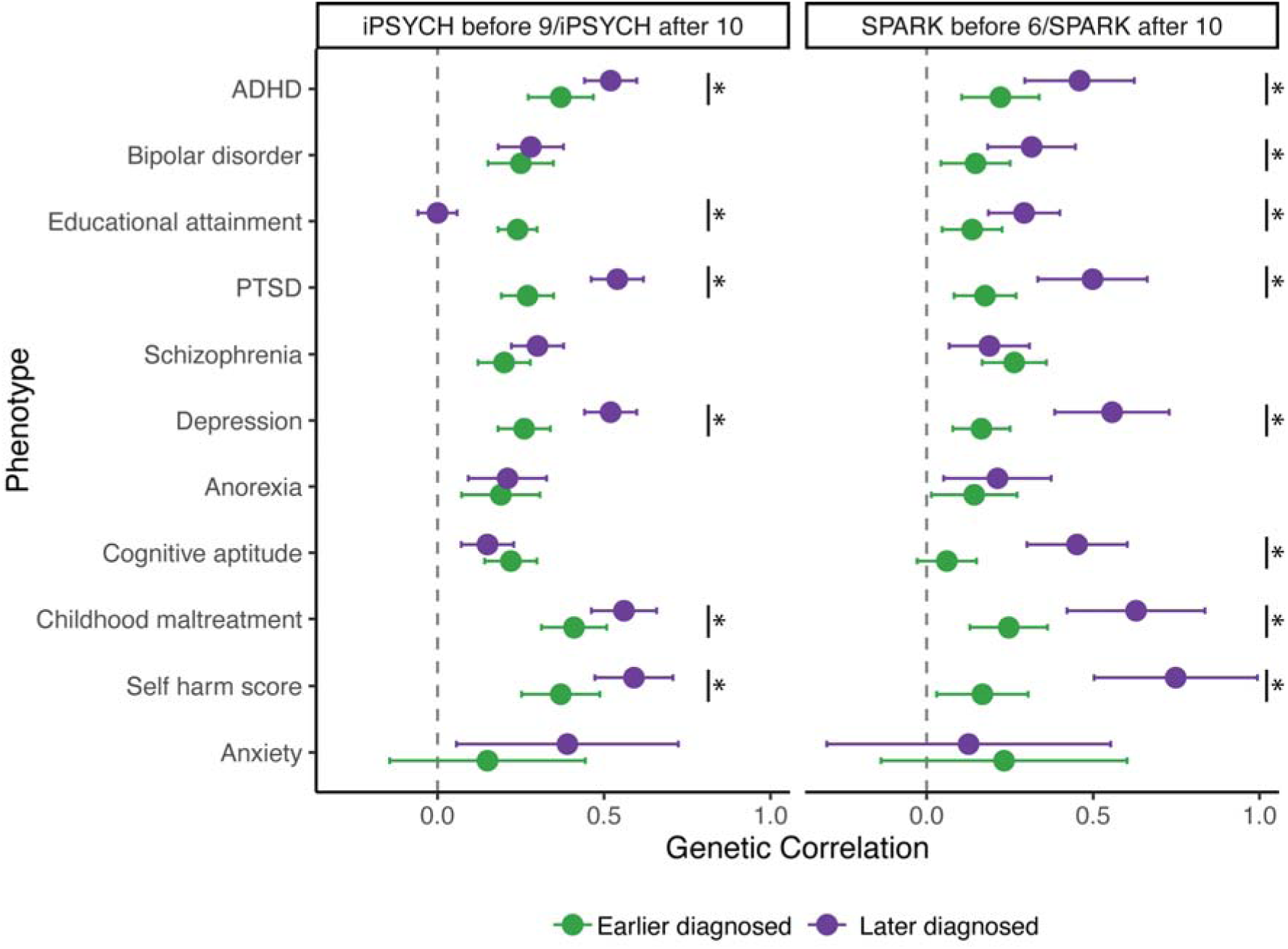
Within-cohort genetic correlation between age at diagnosis stratified autism GWAS and mental health and cognition related phenotypes. Genetic correlation between age at autism stratified GWAS in SPARK (meta-analysed from Discovery and Replication cohorts) and iPSYCH and other mental health and cognition related phenotypes. Points represent genetic correlation estimates and whiskers indicate 95% confidence intervals. Green represents the earlier diagnosed autism GWAS (iPSYCH before 9 and SPARK before 6), and purple represents later diagnosed autism GWAS (iPSYCH and SPARK after 10). Asterisk (*) indicates significantly different genetic correlation between the earlier and later diagnosed GWAS (P < 0.05, two-tailed Z test).

