## Supplementary Notes, Figures, and FAQs for "Polygenic and developmental profiles of autism differ by age at diagnosis"

**Note 1: Overview of the birth cohorts	3**

Millennium Cohort Study (MCS)	3

Longitudinal Study of Australian Children (Birth and Kindergarten, LSAC-B and LSAC-K)	3

Growing Up in Ireland	4

ALSPAC	4

**Note 2: Imputation	6**

Note 2, Figure 1	7

Note 2, Figure 2	8

**Note 3: GMM river plots and explanation	9**

Note 3, Figure 1	10

Note 3, Figure 2	11

Note 3, Figure 3	12

Note 3, Figure 4	13

Note 3, Figure 5	14

Note 3, Figure 6	15

Note 3, Figure 7	16

Note 3, Figure 8	17

**Note 4: Mediation analyses	18**

**Note 5: Latent growth curve modelling (LGCM)	19**

**Note 6: Genetic correlation among different age at autism diagnoses GWAS	21**

Note 6, Figure 1	22

**Note 7: Association between autism PGS and age at autism diagnosis	23**

Note 7, Figure 1	25

**Note 8: Genetic decomposition of the autism effects	26**

Note 8, Figure 1	26

Note 8, Figure 2	28

**Note 9: Earlier and later diagnosed autism are differently associated with developmental trajectories	29**

Note 9, Figure 1	31

Note 9, Figure 2	32

**Note 10: Impact of various demographic and clinical characteristics on age at autism diagnosis	33**

1. Clinical factors: Changes in diagnostic criteria over time	33

2. Clinical factors: Intellectual disability and co-occurring developmental delays	34

3. Clinical factors: Country/continent level differences in diagnosis	35

4. Demographic factors: Sex	35

5. Demographic factors: Parental characteristics	35

**Supplementary Figure 1	37**

**Supplementary Figure 2	38**

**Supplementary Figure 3	39**

**Supplementary Figure 4	40**

**Supplementary Figure 5	41**

**Supplementary Figure 6	42**

**Supplementary Figure 7	43**

**Supplementary Figure 8	44**

**Supplementary Figure 9	45**

**Supplementary Figure 10	48**

**Supplementary Figure 11	50**

**References	51**

**Summary	55**

**FAQs	56**

**Glossary of terms	60**

### Note 1: Overview of the birth cohorts

#### Millennium Cohort Study (MCS)

The Millennium Cohort Study is a longitudinal study that follows the lives of approximately 19,000 children born in 2000 - 2001, along with their families, across the United Kingdom. The study participants were born over a 12-month period starting in September 2000 in England and Wales, and over a 13.5-month period starting in November 2000 in Scotland and Northern Ireland.

The sample design ensured an overrepresentation of families residing in areas with high levels of child poverty and areas in England with significant ethnic minority populations. The initial data collection was conducted through a home-based survey when the children were nine months old, gathering information on various aspects, including circumstances surrounding pregnancy and birth, early life experiences, and the socio-economic backgrounds of the families. Subsequent data collection waves have occurred at ages 3, 5, 7, 11, 14, and 17, allowing researchers to track the development and life trajectories of these children as they progress through various stages of life.

Ethical approval was obtained from Multi-centre Research Ethics Committees.

This cohort was used in the Growth Mixture Model, Latent Growth Curve Model, and Polygenic Score analyses.

#### Longitudinal Study of Australian Children (Birth and Kindergarten, LSAC-B and LSAC-K)

*Growing Up in Australia*: The Longitudinal Study of Australian Children (LSAC) is a longitudinal study that follows two cohorts of about 5,000 children and their families randomly selected from across Australia. The sampling strategy ensured that the number of children selected reflects the overall child population distribution across the Australian states/territories. The B (“baby”) cohort comprises children born between March 2003 and February 2004 (aged 0-1 years in the first data collection sweep); the K (“kindergarten”) cohort comprises children born between March 1999 and February 2000 (aged 4-5 years in the first data collection sweep). Since 2004, data collections have happened biennially using multiple methods, including face-to-face interviews, computer-assisted telephone interviews, self-completed questionnaires, physical measures, and linking to administrative data (e.g., MySchool). From Sweep 3 onwards, there is data on children of the same age from both cohorts at different time points, featuring the unique LSAC “accelerated cross-sequential” design.

These cohorts were used in the Growth Mixture Model and Latent Growth Curve Model analyses.

|  | Sweep 1 (2004) | Sweep 2 (2006) | Sweep 3 (2008) | Sweep 4 (2010) | Sweep 5 (2012) | Sweep 6 (2014) | Sweep 7 (2016) | Sweep 8 (2018) | Sweep 9 (2020-21) |
| --- | --- | --- | --- | --- | --- | --- | --- | --- | --- |
| **B**  **Cohort** | 0-1  Years | 2-3  Years | 4-5  Years | 6-7  Years | 8-9  Years | 10-11  Years | 12-13  Years | 13-14  Years | 15-16  Years |
| **K**  **Cohort** | 4-5  Years | 6-7  Years | 7-8  Years | 9-10  Years | 11-12  Years | 12-13  Years | 13-14  Years | 15-16  Years | 17-18  Years |

#### Growing Up in Ireland

Growing up in Ireland (GUI) is a longitudinal study that follows children living in the Republic of Ireland. We used one of the two cohorts, Cohort 98’ (aka the Child Cohort), who were born in 1998. Since 2008, data collections have been conducted every four years, from parents, teachers, and young people themselves. We did not study the other cohort, Cohort 08’ (aka the Infant Cohort), due to the unavailability of information on autism diagnoses in the earliest few sweeps.

This cohort was used in Latent Growth Curve Model analyses. We did not include this in the Growth Mixture Model analyses, as there were only three time points (sweeps) and we were underpowered to identify two or more trajectories.

#### ALSPAC

Pregnant women resident in Avon, UK with expected dates of delivery between 1st April 1991 and 31st December 1992 were invited to take part in the ALSPAC study.^1,2^ 20,248 pregnancies have been identified as being eligible and the initial number of pregnancies enrolled was 14,541. Of the initial pregnancies, there were a total of 14,676 foetuses, resulting in 14,062 live births and 13,988 children who were alive at 1 year of age. When the oldest children were approximately 7 years of age, an attempt was made to bolster the initial sample with eligible cases who had failed to join the study originally. As a result, when considering variables collected from the age of seven onwards (and potentially abstracted from obstetric notes) there are data available for more than the 14,541 pregnancies mentioned above: The number of new pregnancies not in the initial sample (known as Phase I enrolment) that are currently represented in the released data and reflecting enrolment status at the age of 24 is 906, resulting in an additional 913 children being enrolled. The total sample size for analyses after the age of seven is therefore 15,447 pregnancies, resulting in 15,658 foetuses. Of these 14,901 children were alive at 1 year of age.

Of the original 14,541 initial pregnancies, 338 were from women who had already enrolled with a previous pregnancy, meaning 14,203 unique mothers were initially participating in the study. As a result of the additional phases of recruitment, a further 630 women who did not enrol originally have provided data since their child was 7 years of age. This provides a total of 14,833 unique women (G0 mothers) enrolled in ALSPAC as of September 2021.

Please note that the study website contains details of all the data that is available through a fully searchable data dictionary and variable search tool: <http://www.bristol.ac.uk/alspac/researchers/our-data/>

Ethical approval for the study was obtained from the ALSPAC Ethics and Law Committee and the Local Research Ethics Committees. Consent for biological samples has been collected in accordance with the Human Tissue Act (2004)^3^. Informed consent for the use of data collected via questionnaires and clinics was obtained from participants following the recommendations of the ALSPAC Ethics and Law Committee at the time.

This cohort was included in the PGS analyses. In ALSPAC, we had access to autism assessments only at one time point and were unable to include it in the Growth MIxture and Latent Growth Curve Models.

### Note 2: Imputation

To examine the impact of our sample restrictions, we conducted imputation in the full sample of autistic children identified in MCS (N = 623), compared to the smaller MCS sample with more stringent inclusion criteria used for primary analyses (N = 188). Across the SDQ variables, missingness ranged from 9.47% to 38.52%, with higher missingness observed in later sweeps. Sociodemographic variables had moderate missingness of around 12%, cognitive ability and developmental milestone variables showed variable patterns of missingness ranging from 5% to over 25% (see details in **Supplementary Table 5**).

Imputation of missing data in MCS was done using SoftImpute^4^, an imputation method based on matrix completion techniques. SoftImpute employs a soft-thresholding operation on the singular value decomposition (SVD) of the incomplete data matrix, iteratively refining the approximation of missing values. Leveraging low-rank matrix approximation, SoftImpute is particularly suitable for large-scale datasets due to its computational efficiency. Moreover, by focusing on the underlying structure of the data instead of fitting specific predictive models for each variable type, as in MICE, SoftImpute can effectively capture and preserve complex relationships and interactions over time in our data.

In this study, the regularisation parameter *λ* was set to the largest possible values, determined by the maximum singular value obtained from the Singular Value Decomposition of the input matrix. This choice encourages simpler solutions with lower-rank structures, promoting generalisation and preventing overfitting of the imputed data. Also, the maximum rank (*rank.max*), which determines the maximum number of singular values to retain during the low-rank approximation, was set to be one less than the minimum dimension of the input matrix, enabling retention of as many patterns as possible while maintaining computational efficiency. Auxiliary variables, including sociodemographic variables and developmental milestones, were included in the imputation to improve the precision of estimated values.

Visual inspection of density plots (**Note 2, Figures 1 and 2**) and *Kolmogorov-Smirnov (KS)* tests demonstrated that the distributions of imputed data restored that of the original data (see **Supplementary Table 5**). However, imputation quality was poorer for subscale scores at age 17, possibly due to high attrition.

SDQ total difficulties scores were then calculated from imputed subscale scores in the corresponding sweeps. To generate principal components of sociodemographic factors and cognition, imputed data were combined with the non-imputed raw data in MCS. Subsequently, principal component analyses were conducted separately for (i) cognitive aptitude measures; (ii) household socio-economic status; and (iii) living area deprivation following the procedures outlined in **Methods.**

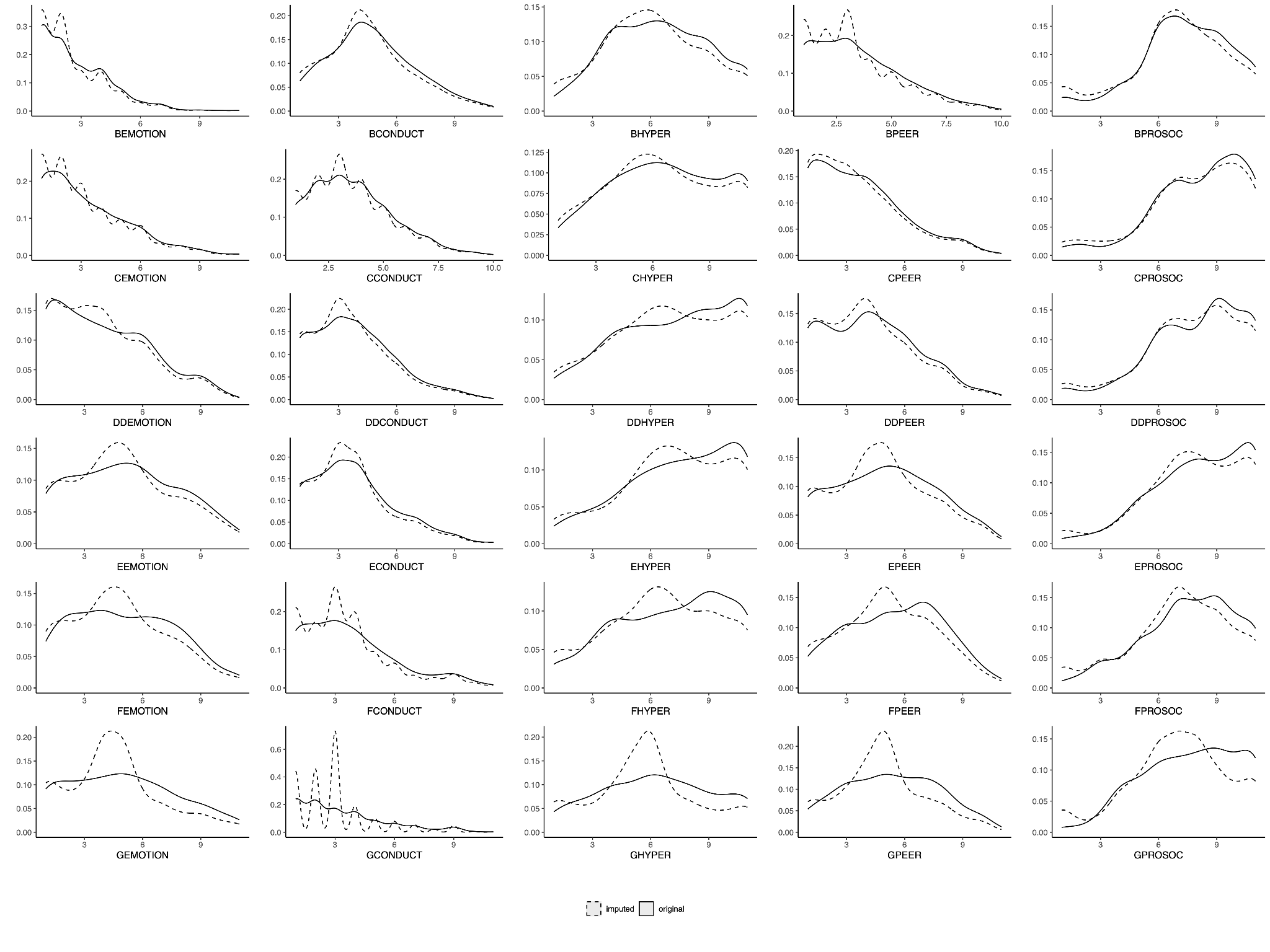

##### Note 2, Figure 1

*Density plots of raw and imputed SDQ subscale scores for imputation quality evaluation. Solid lines represent original raw data, while dashed lines represent imputed data. For each graph, the first letter represents the sweep using an ordinal coding system (e.g.,B = Sweep 2 at age 3, ..., G = Sweep 7 at age 17). EMOTION = Emotional Symptoms; CONDUCT = Conduct Problems; HYPER = Hyperactivity/Inattention; PEER = Peer Relationship Problems; PROSOC = Prosocial Behaviours.*

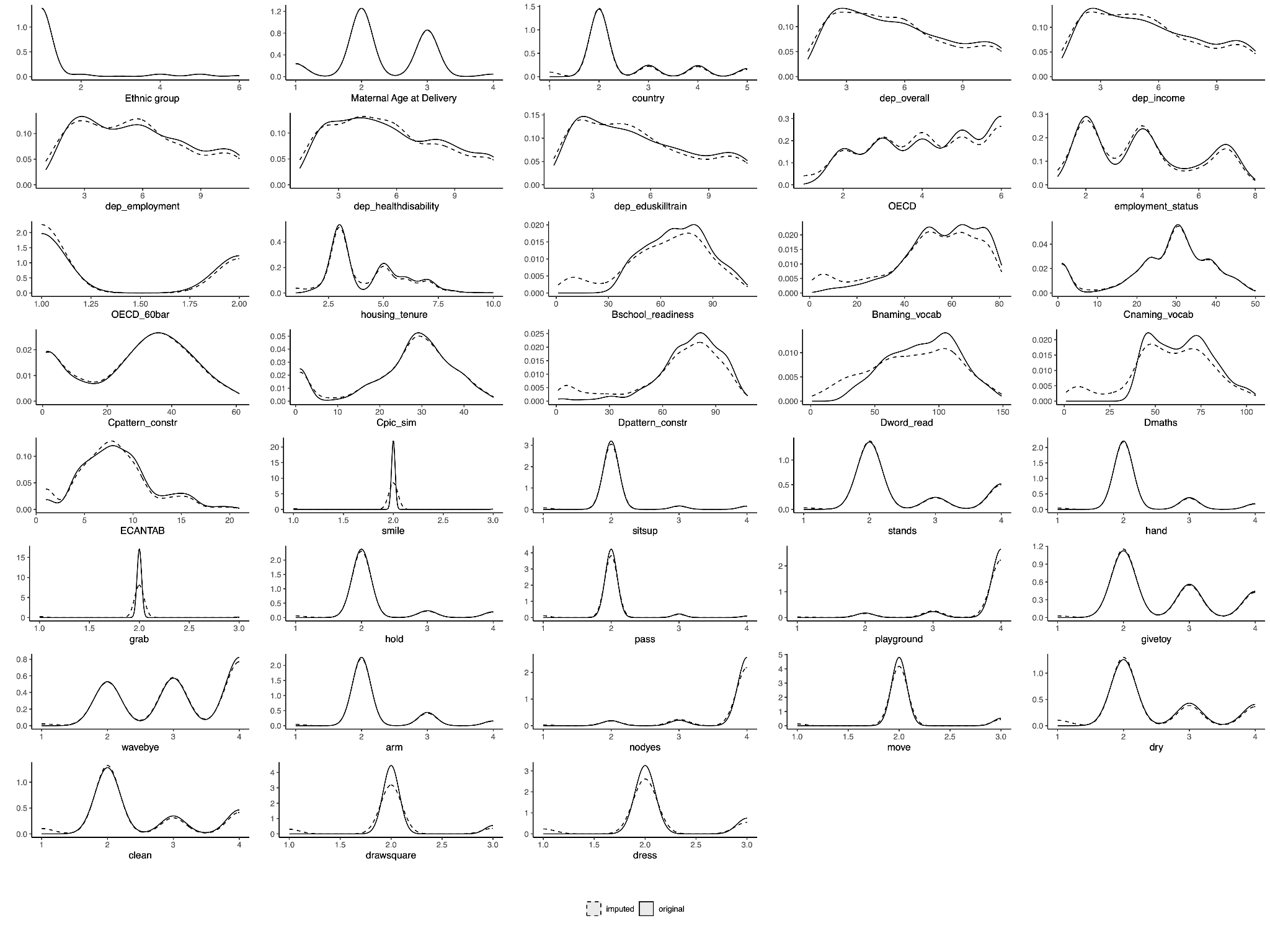

##### Note 2, Figure 2

*Density plots of raw and imputed auxiliary variables for imputation quality evaluation. Solid lines represent original raw data, while dashed lines indicate imputed data. dep_ = deprivation; OECD = Organisation for Economic Co-operation and Development score; OECD_60bar = OECD below 60% Poverty Indicator; naming_vocab = Naming Vocabulary; pattern_constr = Pattern Construction; pic_sim = Picture Similarity; word_read = Word Reading; CANTAB = Cambridge Neuropsychological Test Automated Battery Verbal Similarities Total Correct Responses. stands = stands up holding on; hands = puts hands together; grab = grabs objects; hold = holds small objects; pass = passes a toy; playground = Frequency take child to park or playground; givetoy = gives toy; wavebye = waves bye-bye; arm = Extend his arms for being picked up; nodyes = nods for yes; move = can move from place to place; dry = child dry during the day; clean = child clean during the day; drawsquare = can draw or copy a square; dress = can dress without help. Further details in* ***Supplementary Table 5****.*

### Note 3: GMM river plots and explanation

In the Growth Mixture Model (GMM) analyses in the expanded MCS sample including individuals with co-occurring ADHD and the imputed MCS autism sample, the optimal GMM identified more than two latent trajectory classes in some SDQ subscales (**Supplementary Tables 3 and 5**). However, the most prominent drop in BIC values was observed between models with one and two groups, indicating that the two-group solution captured the most substantial increase in model fit. This pattern was consistent across subscales, including SDQ total difficulties scores, where two-group GMMs were again identified as optimal and used in subsequent mediation analyses, in line with previous findings (**Supplementary Tables 5 and 8**).

River plots (**Note 3, Figures 3 and 8**) plotting changes in latent trajectory memberships between the two-class model and the optimal model (three or four latent) for different subscales (**Note 3, Figures 1, 2, and 4 - 7**) show that the additional latent trajectories were predominantly subsets of the original two latent trajectories. These are indicative that although two broad trajectories remain even with expanded sample sizes, these can be further decomposed into smaller groups with different trajectories.

For instance, in the GMM analyses of the SDQ Conduct problems subscale on the expanded MCS autism sample including participants with co-occurring ADHD, the two-group model identified two trajectories: one with stably decreasing conduct problems (green latent trajectory) and another with increasing conduct problems, particularly after age 7 (purple latent trajectory) (**Note 3, Figure 1A**). The optimal three-trajectory model further refined these trajectories. It identified a consistently low conduct problems group, largely composed of individuals from the green class in the two-trajectory model (**Note 3, Figure 3A**). Children in the late childhood emergent group predominantly remained in the overall increasing class (purple in both models). Stacked bar charts indicated that more children in the increasing groups (purple) were diagnosed at or after age 7 in both models (**Note 3, Figure 1**). Additionally, a distinct third group with initially high but progressively decreasing conduct problems was identified; their conduct problems declined to become the second-lowest (blue latent trajectory) (**Note 3, Figure 1B**).

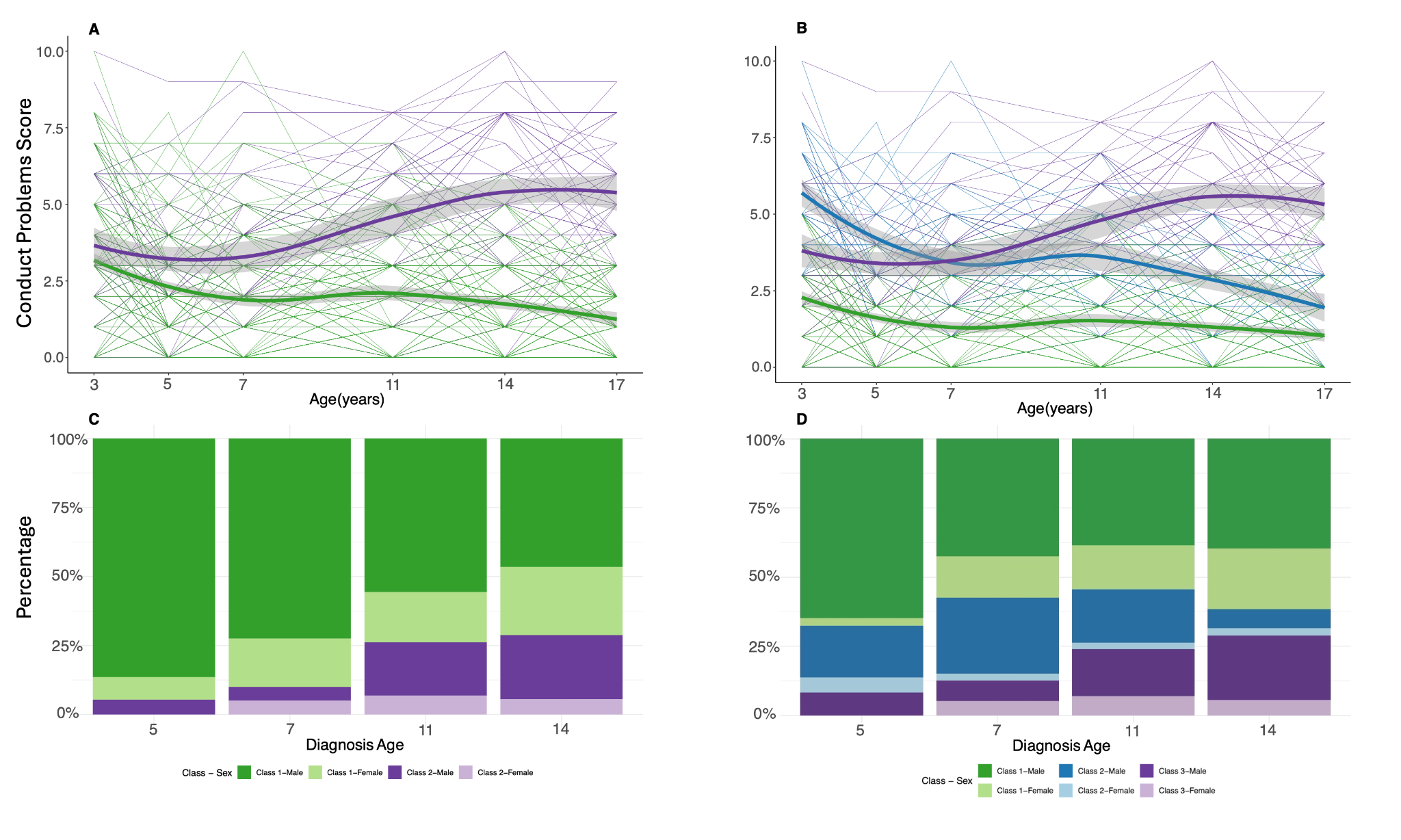

##### Note 3, Figure 1

*In the expanded MCS autism sample including children with co-occurring ADHD, A: Two-group longitudinal growth mixture model of SDQ Conduct Problems score . B: The optimal longitudinal growth mixture model of SDQ Conduct Problems score, demonstrating three latent trajectories. Shaded areas indicate 95% confidence intervals in A - B. C-D: Stacked bar charts providing proportion of individuals diagnosed as autistic at specific ages by the latent trajectory membership from the corresponding growth mixture models. Darker colours indicate males and lighter colours indicate females.*

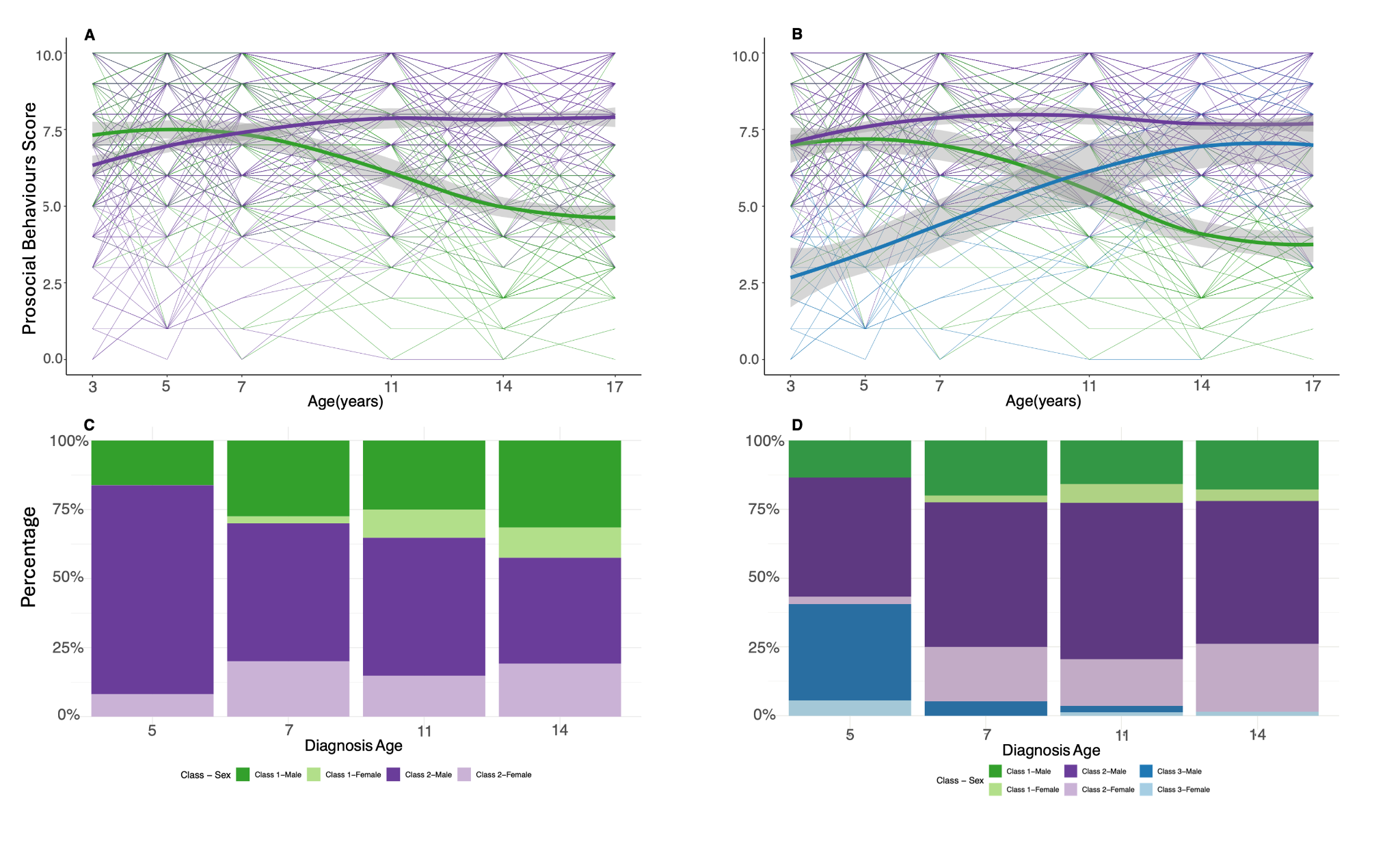

##### Note 3, Figure 2

*In the expanded MCS autism sample including children with co-occurring ADHD, A: Two-group longitudinal growth mixture model of SDQ Prosocial Behaviours score . B: The optimal longitudinal growth mixture model of SDQ Conduct Problems score , demonstrating three latent trajectories. Shaded areas indicate 95% confidence intervals in A - B. C-D: Stacked bar charts providing proportion of individuals diagnosed as autistic at specific ages by the latent trajectory membership from the corresponding growth mixture models. Darker colours indicate males and lighter colours indicate females.*

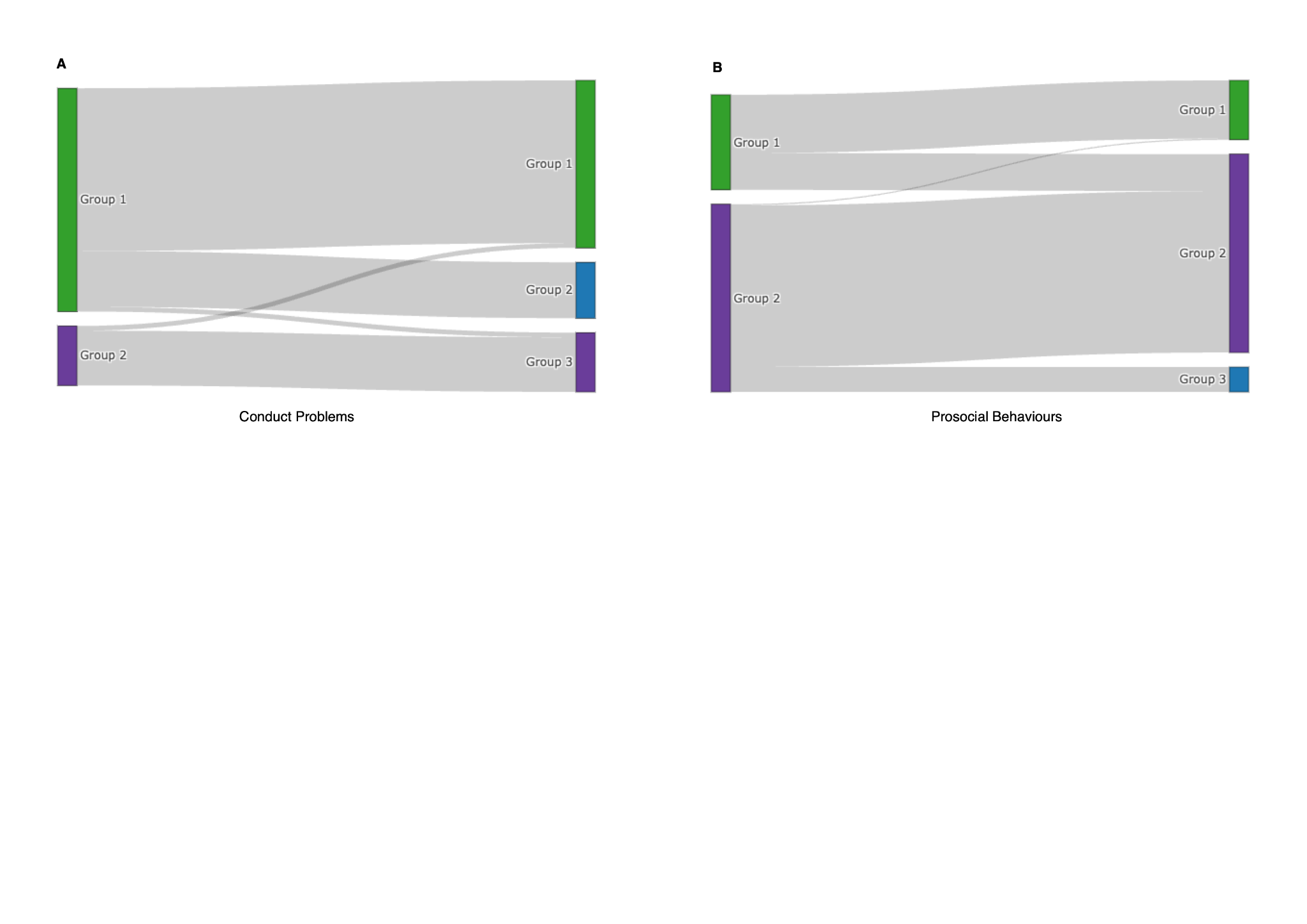

##### Note 3, Figure 3

*River plots (Sankey Plots) of Conduct Problems (A) and Prosocial Behaviours (B) for the expanded MCS autism sample including children with co-occurring ADHD, , illustrating the changes in latent trajectory group memberships between the two-group GMM and the optimal three-group GMM in this expanded sample. Colours are consistent with Note 3, Figures 1 and 2.*

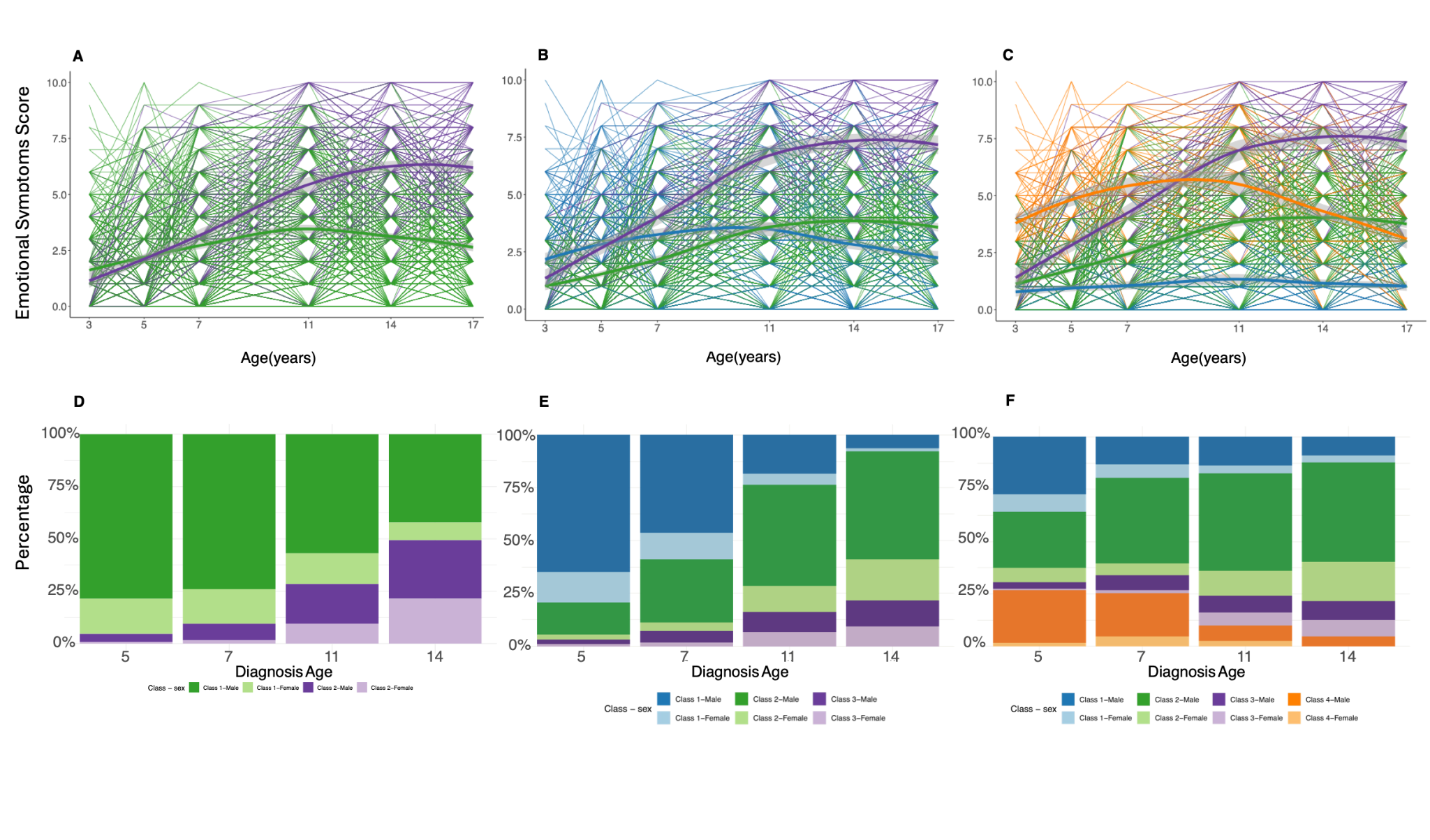

##### Note 3, Figure 4

*In the imputed MCS autism sample, A: Two-group longitudinal growth mixture model of SDQ Emotional Symptoms score. B: Three-group longitudinal growth mixture model of SDQ Emotional Symptoms score C: The optimal longitudinal growth mixture model of SDQ Emotional Symptoms score , demonstrating four latent trajectories. Shaded areas indicate 95% confidence intervals in A - C. D-F: Stacked bar charts providing proportion of individuals diagnosed as autistic at specific ages by the latent trajectory membership from the corresponding growth mixture models. Darker colours indicate males and lighter colours indicate females.*

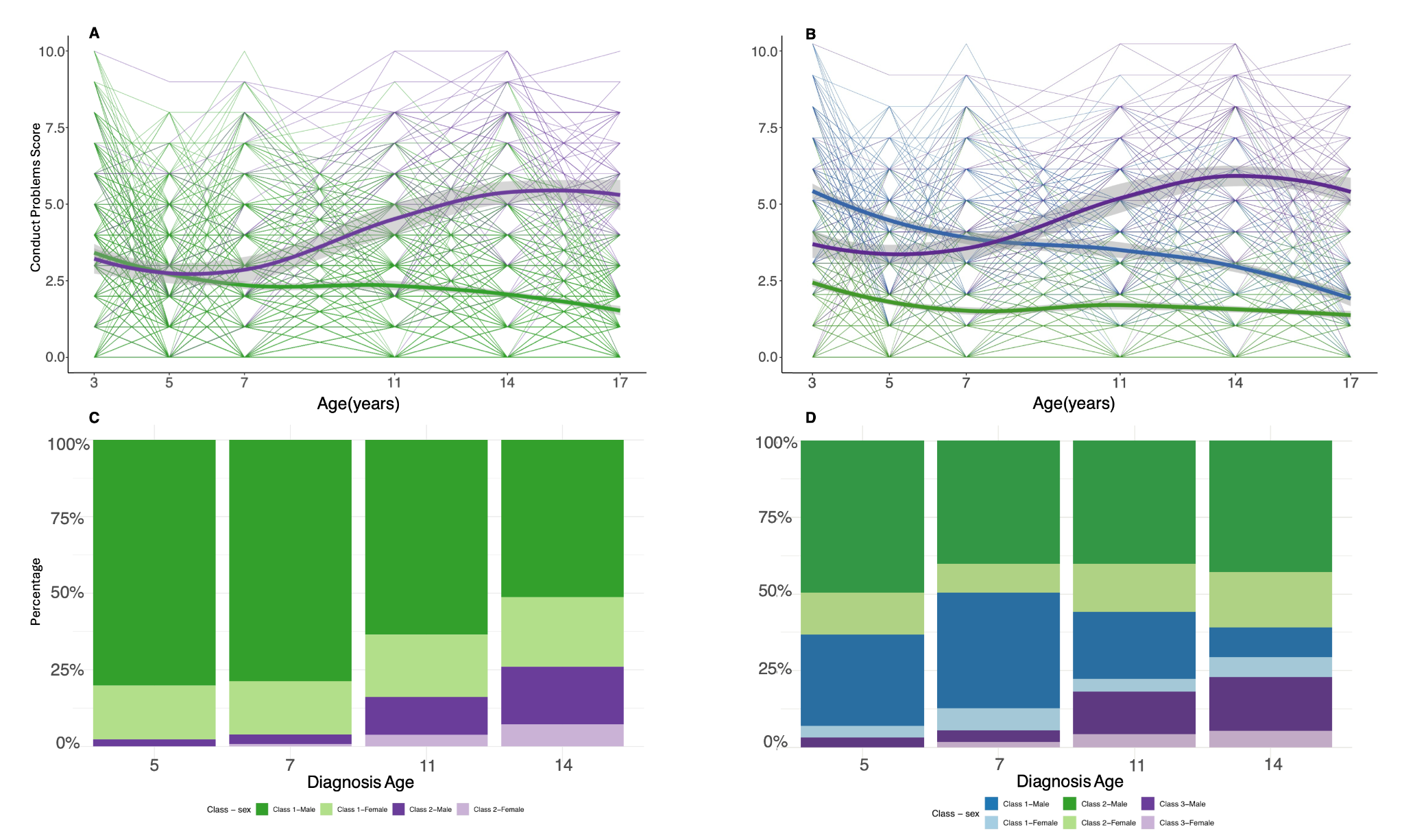

##### Note 3, Figure 5

*In the imputed MCS autism sample, A: Two-group longitudinal growth mixture model of SDQ Conduct Problems score B: The optimal longitudinal growth mixture model of SDQ Conduct Problems score, demonstrating three latent trajectories. C-D: Stacked bar charts providing proportion of diagnosed as autistic at specific ages by the latent trajectory membership from the corresponding growth mixture models. Darker colours indicate males and lighter colours indicate females.*

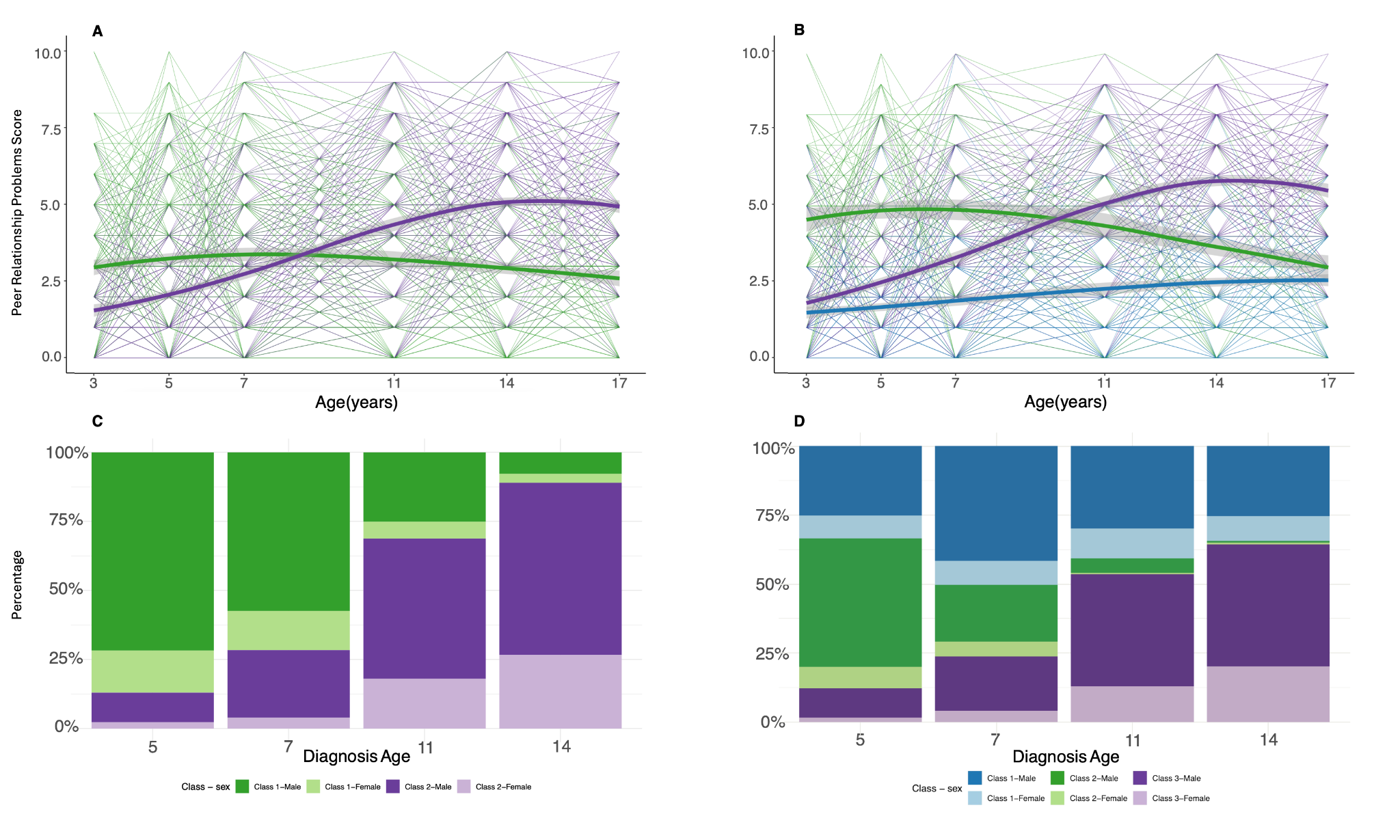

##### Note 3, Figure 6

*In the imputed MCS autism sample, A: Two-group longitudinal growth mixture model of SDQ Peer Relationship Problems score. B: The optimal longitudinal growth mixture model of SDQ Peer Relationship Problems score, demonstrating three latent trajectories. C-D: Stacked bar charts providing proportion of individuals diagnosed as autistic at specific ages by the latent trajectory membership from the corresponding growth mixture models. Darker colours indicate males and lighter colours indicate females.*

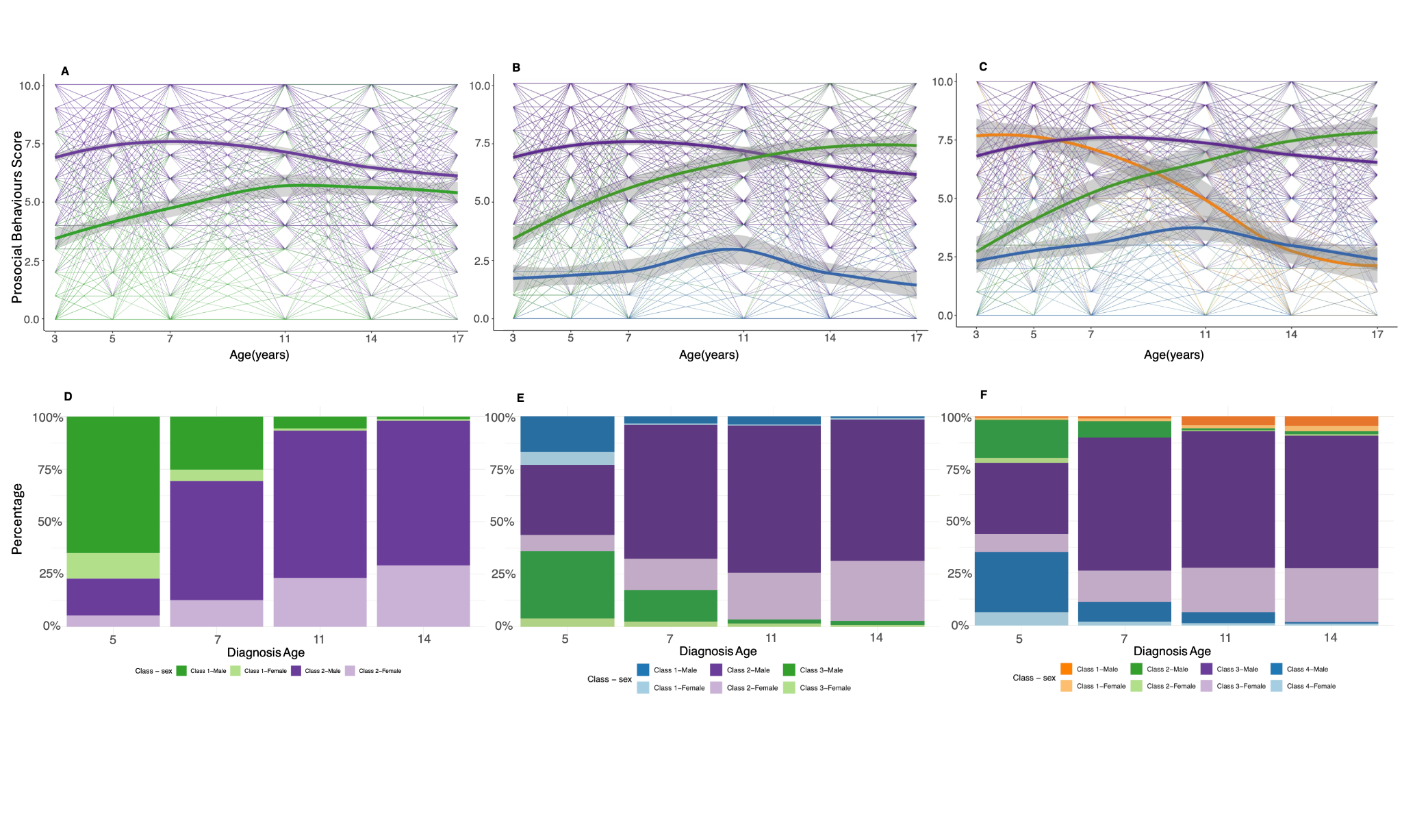

##### Note 3, Figure 7

*In the imputed MCS autism sample, A: Two-group longitudinal growth mixture model of SDQ Prosocial Behaviours score. B: Three-group longitudinal growth mixture model of SDQ Prosocial Behaviours score. C: The optimal longitudinal growth mixture model of SDQ Prosocial Behaviours score, demonstrating four latent trajectories. Shaded areas indicate 95% confidence intervals in A - C. D-F: Stacked bar charts providing proportion of individuals diagnosed as autistic at specific ages by the latent trajectory membership from the corresponding growth mixture models. Darker colours indicate males and lighter colours indicate females.*

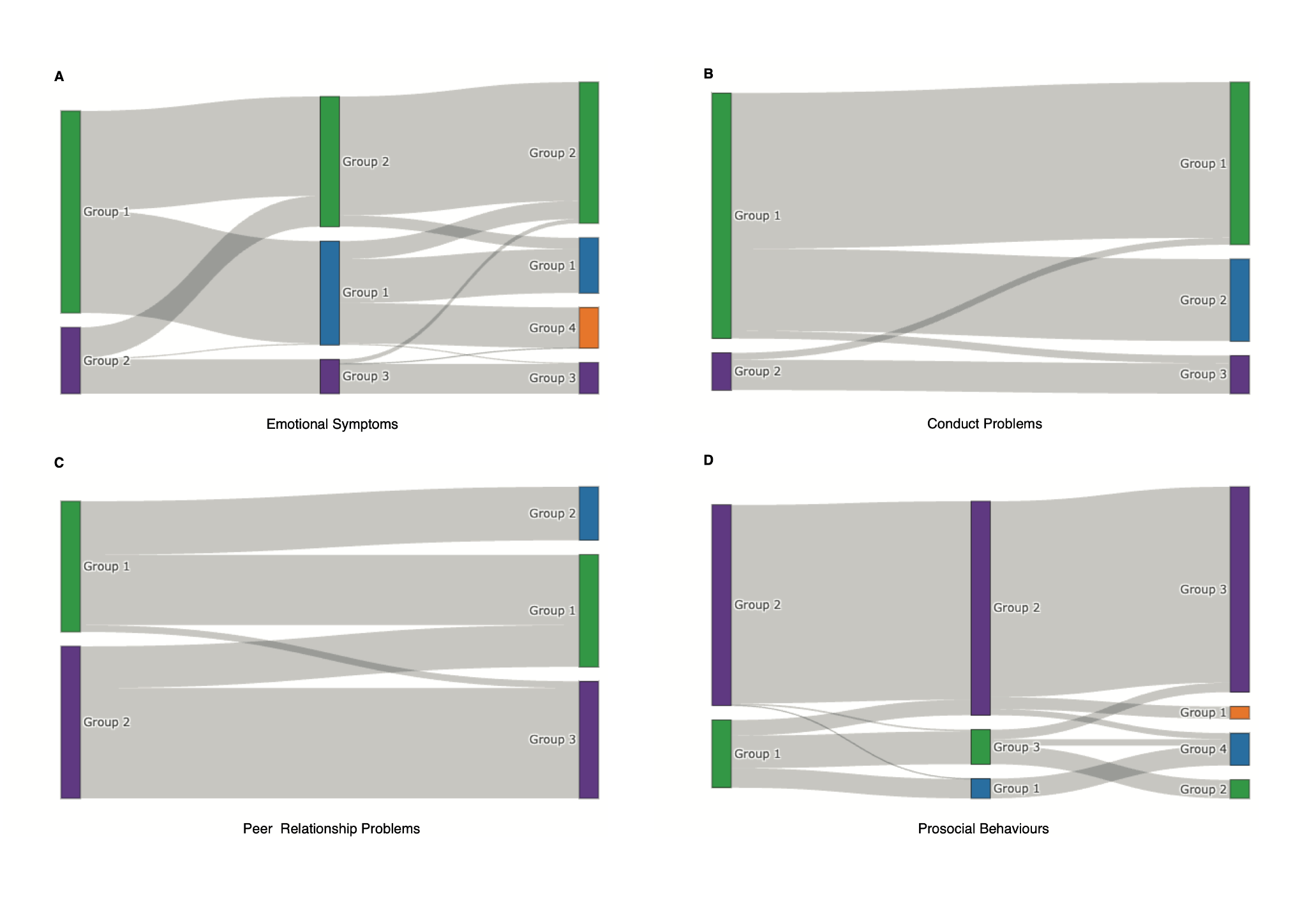

##### Note 3, Figure 8

*River plots (Sankey Plots) of Emotional Symptoms (A), Conduct Problems (B), Peer Relationship Problems (C) and Prosocial Behaviours (D) for the imputed MCS autism sample, illustrating changes in latent trajectory group memberships between the two-group GMM and the optimal GMM (three or four groups identified). Colours are in alignment with Note 3, Figures 4-7.*

### Note 4: Mediation analyses

We used multiple regression to assess the relationship between SDQ latent trajectories identified from the Growth Mixture Models and age at autism diagnosis in autistic individuals across MCS samples (primary, expanded sample to include participants with co-occurring ADHD, and imputed), LSAC-B, and LSAC-K (**Supplementary Table 8**). There were relatively modest effects of sex and other sociodemographic variables on age at autism diagnosis. To test whether the effects of sex and sociodemographic variables are mediated by SDQ latent trajectories, we conducted a mediation analysis with the latent trajectories serving as mediators. Specifically, using structural equation modelling, we modelled the direct effects of latent trajectories and sociodemographic variables on age at autism diagnosis, as well as indirect effects through affecting latent trajectory memberships of SDQ total difficulties and subscale scores. Analyses were conducted in R using the *lavaan* package (v. 0.6-19). Indirect effects were calculated as the product of the path coefficients from predictor to mediator and mediator to outcome; total effects were computed as the sum of direct and indirect effects. Standardised path coefficients are reported to enable comparison across variables. Confidence intervals and p-values for indirect effects were derived using nonparametric bootstrapping with 5,000 resamples^5^ (**Supplementary Table 8**).

Most sociodemographic variables did not have significant direct or indirect effects, underscoring the small effect sizes of these factors in explaining the age at autism diagnosis in our samples. Overall, results of both multiple regression and mediation analysis suggest that the latent trajectories explain a relatively large proportion of variance in age at autism diagnosis (**Supplementary Table 8**). Consistent results were obtained for the expanded autism sample with those with co-occurring ADHD and the imputed autism sample in MCS. In ADHD, latent trajectories and sociodemographic factors explained a much smaller proportion of variation in age at ADHD diagnosis compared to autism. The results were primarily driven by the hyperactivity/inattention and peer relationship problems SDQ subscales, demonstrating relative specificity of the findings to autism (**Supplementary Table 6**).

### Note 5: Latent growth curve modelling (LGCM)

In addition to Growth Mixture Models, which are data driven, we used Latent Growth Curve Modelling (LGCM) to investigate whether SDQ total difficulties and subscale score trajectories vary between autistic individuals based on age at autism diagnosis. Specifically, given the increasing number of autistic individuals being diagnosed in adolescence^6,7^, we tested whether there were broad differences in the trajectories of SDQ total difficulties and subscale scores among autistic individuals diagnosed before the ages of 9 - 11 (childhood diagnosed group, N = 39 - 118 across cohorts, **Supplementary Table 2**) and after (adolescent diagnosed group, N = 27 -73 across cohorts) (**Methods**). This age cutoff period corresponds to the onset of puberty, the transition from primary to secondary school, and the beginning of an increase in incidence of autism diagnosis in girls^8^^,^^9,10^. The specific cutoff age was cohort-dependent, as different birth cohorts collected information on autism diagnosis at different ages, further details of which are provided in the Methods.

We conducted LGCM in four cohorts: MCS, LSAC-B, LSAC-K, and GUI (**Extended Data Figure 3**). Across all four cohorts, LGCM identified different trajectories of SDQ total difficulties scores, peer relationship problems, and prosocial behaviours between the childhood and adolescent diagnosed groups (**Supplementary Figures 1 - 6, Supplementary Figure 9**). Compared with individuals without an autism diagnosis at any time point, the childhood diagnosed autistic group had higher difficulties in early childhood that remained relatively stable or gently declined in adolescence. Compared to the childhood diagnosed autistic group, the adolescent diagnosed autistic group had fewer difficulties during early childhood, but difficulties increased in later childhood and persisted into adolescence.

In MCS, we ran a series of sensitivity analyses to check the robustness of the above results. We obtained consistent results: (1) in an expanded sample of MCS that included autistic individuals with co-occurring Attention-Deficit/Hyperactivity Disorder (ADHD) and inconsistent reports of an autism diagnosis (**Methods**, **Supplementary Table 9**); (2) after imputing missing SDQ scores (**Supplementary Table 5, Supplementary Note 2**); and (3) when restricting to males. In all four birth cohorts, models stratified by age at diagnosis were generally a better fit to the data than sex-stratifed models (**Supplementary Table 9**), suggesting that the results do not primarily reflect sex differences in age at autism diagnosis.

While our primary analyses utilised cohort-specific thresholds corresponding to ages 9-11, we explored the robustness of this approach to age cutoff choices. In GUI, autism diagnosis assessment was unavailable at age 11 and only available at age 13. Conversely, LSAC-B had insufficient numbers of children diagnosed at age 13 and beyond, making latent growth curve models using a later age at diagnosis threshold underpowered. Nevertheless, we conducted sensitivity analyses in MCS and LSAC-B using various age at diagnosis cutoffs (**Supplementary Figure 10**). Consistent with previous analyses, groups with a higher proportion of later diagnoses exhibited lower baseline difficulties (intercepts) but steeper increases over time (slopes) across SDQ subscales. Models with smaller group sizes showed estimation issues in LSAC-B (e.g., negative latent variances for a parameter or a non-positive definite latent covariance matrix), supporting our choice of the age cut in dichotomous-group analysis.

To assess the specificity for autism, we ran LGCMs on SDQ total difficulties and subscale scores with children with an ADHD but without an autism diagnosis in the MCS cohort (N = 89). Children with ADHD diagnosed in childhood and adolescence differed only nominally in the slopes of the hyperactivity/inattention (P = 0.026) and prosocial behaviour subscales (P = 0.029) (**Supplementary Table 6 and Supplementary Figures 7 and 8**). Compared to adolescent diagnosed children with ADHD, adolescent diagnosed autistic children had a steeper increase in peer relationship problems (P = 5.77x10^-3^) and emotional symptoms (P = 0.012) across development. However, these results must be interpreted cautiously given the small sample size.

### Note 6: Genetic correlation among different age at autism diagnoses GWAS

In sensitivity analyses, we tested whether the moderate genetic correlation between the SPARK and iPSYCH age at autism diagnosis GWASs can be attributed to the differing median age at diagnosis. We tested this in two ways.

First, in the SPARK discovery cohort, we generated five overlapping subsets of autistic individuals (N = 9,650 - 9,704) with varying median age at autism diagnosis (**Note 6,** **Figure 1A**), and generated age at autism diagnosis GWAS in each of these subsets. We conducted genetic correlation using LDSC with the iPSYCH age at diagnosis GWAS, which had a median age at diagnosis of 10 years (MAD = 4). As there is no participant overlap between the two GWAS and because linear mixed effect models can account for fine-scale population stratification, we constrained the cross-trait LD score regression intercept to zero to reduce the standard errors of the genetic correlations. As the median age at diagnosis between the SPARK subset and iPSYCH became more similar, the genetic correlation approached 1 (**Note 6, Figure 1A)**.

Second, we stratified the iPSYCH autistic population into those diagnosed before age 11 and those diagnosed after age 10, and conducted age at diagnosis GWAS analyses in both these subsets. We calculated genetic correlation (LDSC, cross-trait intercept constrained to zero) with the SPARK (meta-analysed) age at diagnosis GWAS. As can be seen in **Note 6, Figure 1B**, as the median age at diagnosis between the iPSYCH subset and SPARK became more similar, the genetic correlation approached 1.

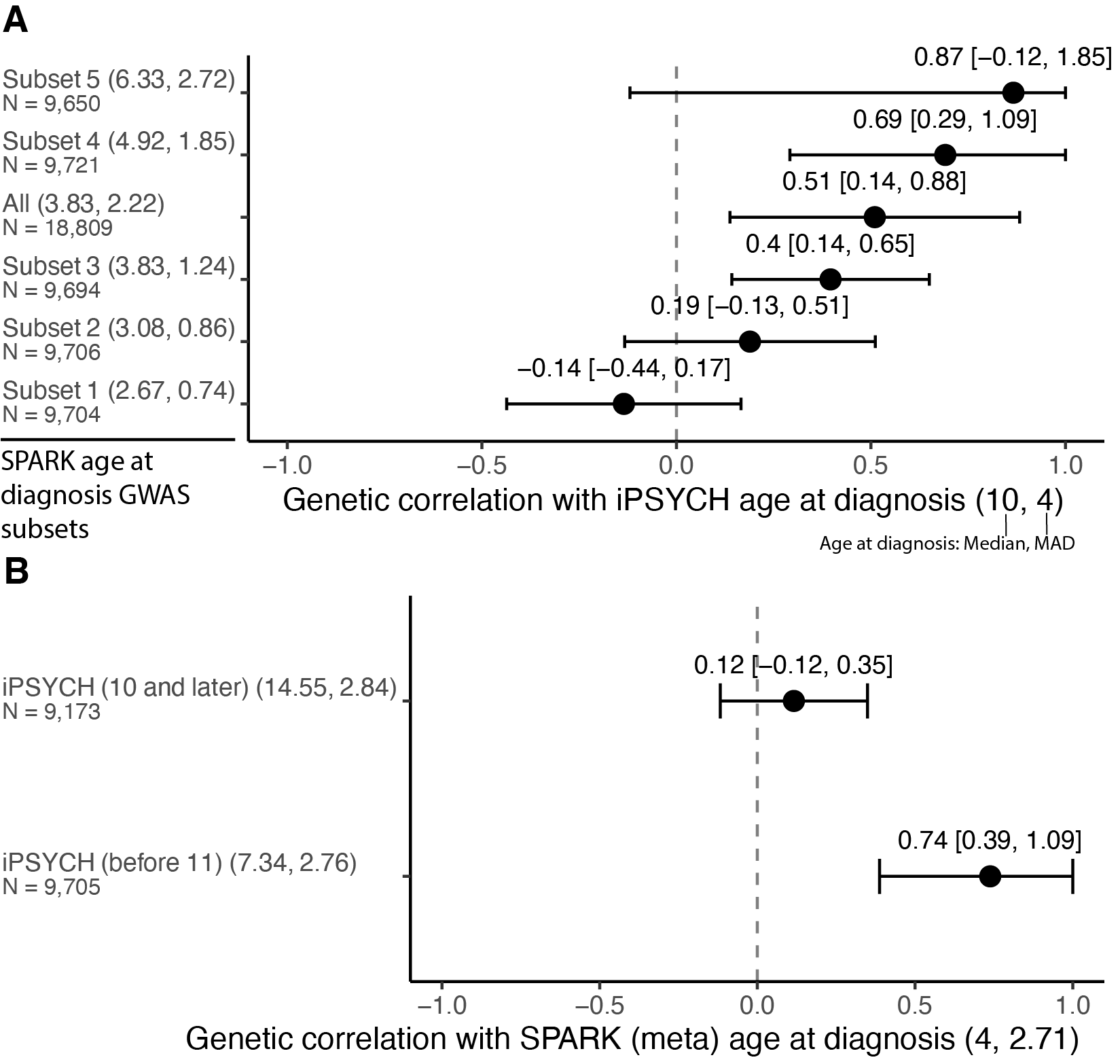

##### Note 6, Figure 1

*Genetic correlations among different age at autism diagnosis GWAS. A. Genetic correlations between age at autism diagnosis in iPSYCH (median age = 10, median absolute deviation = 4) and SPARK discovery subsets arranged by median age at diagnosis. B. Genetic correlation between age at autism diagnosis in SPARK (median age = 4, median absolute deviation = 2.71) and iPSYCH subsets. For both A and B, genetic correlations with 95% confidence intervals (CI) are shown as black points with horizontal error bars. For visualisation purposes, error bars are constrained to the range of -1 to 1, while text labels display the actual, unconstrained genetic correlation and 95% CI values. The vertical dashed line indicates zero correlation. Phenotypes are arranged by median age at diagnosis, with the median and median absolute deviation values in parentheses and sample size (N) below each label.*

### Note 7: Association between autism PGS and age at autism diagnosis

We conducted additional sensitivity analyses to test the hypothesis that there are two underlying polygenic autism factors that correlate with age at autism diagnosis. For these analyses, we used polygenic scores (PGS) derived from iPSYCH_before11_ and iPSYCH_after10_ as indices of earlier and later diagnosed autism respectively. We chose to use PGS generated from these two GWAS over PGS generated from the two autism polygenic factors because the iPSYCH cohort is independent of SPARK, in which we run several sensitivity analyses. For consistency, we used PGS generated from iPSYCH_before11_ and iPSYCH_after10_ in other cohorts as well.

*Within-family analyses*

We first investigated whether both sets of iPSYCH-derived PGS are over-transmitted from parents to their autistic children in the independent SPARK cohort and if this varies by sex and ID. Estimates of PGS association within populations may reflect several confounding factors including fine-scale population stratification, correlation between an individual’s genotypes and their environments, and parental indirect genetic effects wherein parental genes can impact their child’s traits independent of the child’s own genetics. A within-family approach can at least partly account for these three confounding issues, and an over-transmission of the iPSYCH-derived PGS would demonstrate that at least some of the common variant associations detected in the iPSYCH GWASs for earlier- and later-diagnosed autism are causal for autism. Furthermore, we stratified by sex and co-occurring ID: if the PGS are differentially over-transmitted in these subgroups, this would indicate that the iPSYCH-derived PGS have different effects on autism risk in these subgroups. In other words, the effects of the iPSYCH-derived PGS are not homogeneous across sex and/or ID.

We tested this using a polygenic Transmission Disequilibrium Test, which calculates the PGS in (autistic children) and compares them to the parental mean PGS^11^. A significant difference in the child’s PGS compared to the midparental mean is evidence of over- or under-transmission of PGS. Prior work has established an over-transmission of autism PGS to autistic children but not their non-autistic siblings from their parents^11,12^. We extend this here using the two iPSYCH-derived autism PGS.

In SPARK, both PGSs were over-transmitted from parents to their autistic children, with consistent results after stratifying by sex and ID (**Supplementary Table 15**). This confirms that PGS from age-stratified autism GWAS from iPSYCH are associated with autism in SPARK. However, we identified a modestly larger over-transmission of iPSYCH_before11_ compared to iPSYCH_after10_ PGS (P = 7.15x10^-3^) (**Supplementary Table 15, Note 7, Figure 1A**). As over 90% of autistic individuals in SPARK are diagnosed before age 10, this indicates that the effects of the PGS may vary by age at diagnosis.

*Within-cohort analyses*

The above analyses indicate an association between age-at-diagnosis stratified autism PGS and autism that does not reflect population stratification or other confounding factors. We next investigated whether both PGSs are associated with age at autism diagnosis in SPARK even after accounting for several additional clinical and sociodemographic factors. We used the quality controlled, genotyped SPARK cohort for this, conducting separate analyses in the SPARK Discovery and Replication cohorts and meta-analysing the estimates. We were unable to assess this using a within-family approach due to the relatively small number of complete trios with information on covariates.

In SPARK, iPSYCH_after10_ PGS was consistently associated with later age at autism diagnosis even after accounting for several additional clinical and sociodemographic factors (**Supplementary Table 16, Note 7, Figure 1B)**. The iPSYCH_before11_ PGS was associated with earlier age at autism diagnosis in some models, with consistently negative (i.e., associated with an earlier age at diagnosis) regression coefficients across all models. This suggests that genetic influences on age of autism diagnosis cannot be fully explained by these other factors.

Finally, in MCS, iPSYCH_before11_ but not iPSYCH_after10_ PGS was associated with autism diagnosed before age 11 (**Supplementary Table 17, Note 7, Figure 1C**). Taken together, this suggests that although both PGS are associated with autism, their effects on autism vary by age at diagnosis.

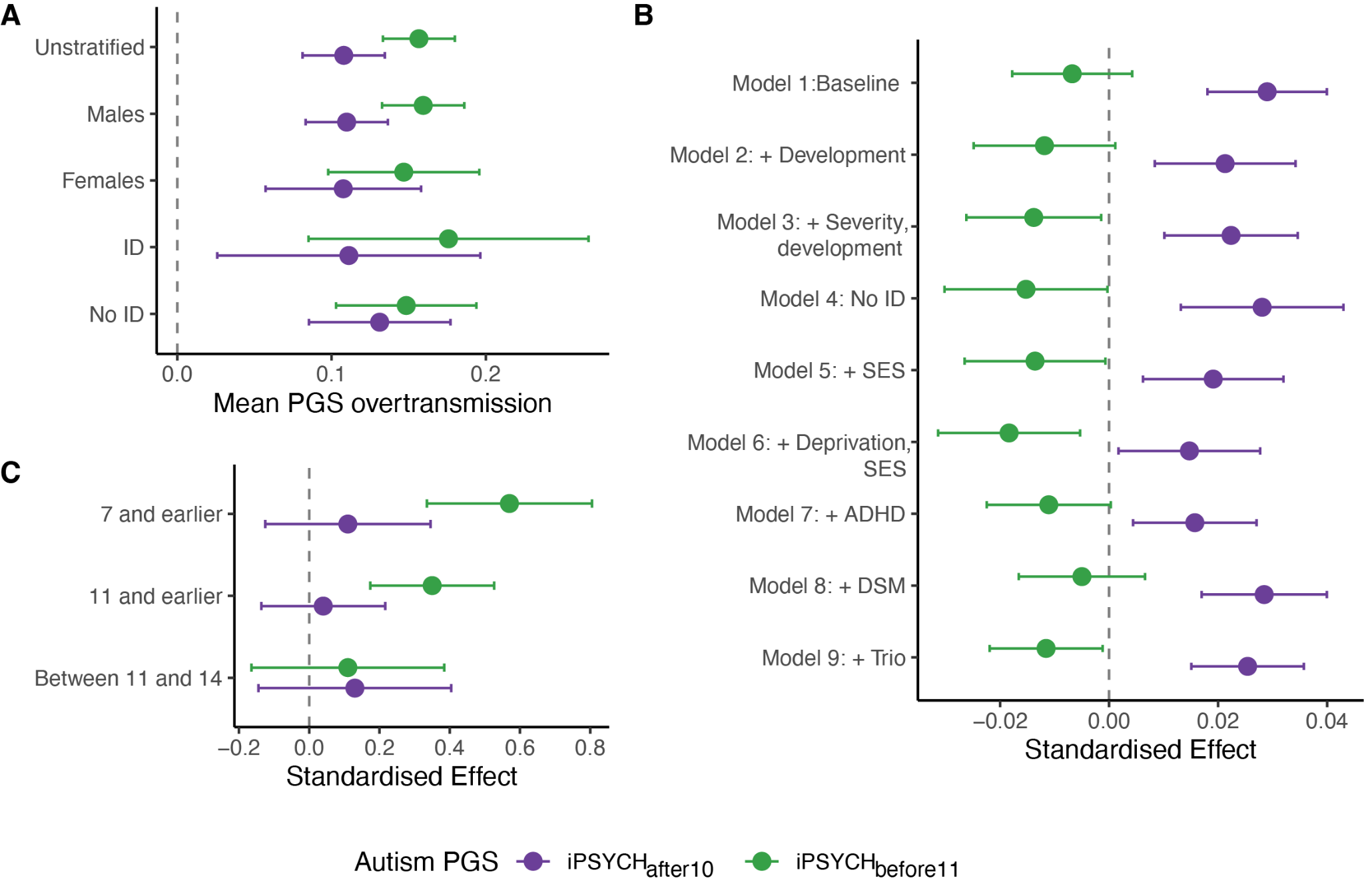

##### Note 7, Figure 1

*PGS association with age at autism diagnosis. A. Over-transmission of PGS for iPSYCH_before11_ and iPSYCH_after10_ from parents to autistic children in the SPARK cohort (meta-analysed estimates). Estimates provided for unstratified, sex-stratified, and ID-stratified analyses. Children’s PGS have been standardised to parental mean PGS, with the line at zero indicating no over-transmission. B. Association between age at autism diagnosis PGS for iPSYCH_before11_ and iPSYCH_after10_ in the SPARK cohort (meta-analysed estimates). Estimates provided after correcting for ID, sex and PCs (baseline), and additionally: Model 2: developmental (dev.) milestones, Model 3: autism severity indicating by SCQ, RBS-R scores, and developmental Milestones, developmental Regression, and IQ scores; Model 4: after excluding individuals with co-occurring ID; Model 5: dev. Milestones and parental socio-economic status (SES); and Model 6: dev. milestones, parental socioeconomic, and area deprivation. In Model 7, we controlled for ADHD, in Model 8 DSM-status (DSM-IV vs DSM-5), and in Model 9, trio status. + indicates that baseline covariates were also included. C. Association between autism diagnosed in childhood and adolescence and PGS for iPSYCH_befoere 11_ and iPSYCH_after10_ GWAS in the MCS cohort. For all plots, points indicate the estimate, whiskers indicate 95% confidence intervals. ID: Intellectual disability; SCQ: Social Communication Questionnaire; RBS-R: Repetitive Behaviours Scale-Revised; DSM: Diagnostic and Statistical Manual of Mental Disorders.*

### Note 8: Genetic decomposition of the autism effects

One framework to characterise the genetics of later diagnosed autism is that it is the additive genetic effects of autism (indexed by earlier diagnosed autism) and a series of other mental health conditions and ADHD, especially among adolescents and adults, where developmental history is difficult to ascertain^13^. This may occur either due to diagnostic misclassification, where someone is incorrectly diagnosed as autistic, or diagnostic overshadowing due to co-occurrence of autism and one or more other neurodevelopmental and psychiatric conditions. We illustrate this schematically in **Note 8, Figure 1**.

The Unitary Model (analogous to the first glass filled entirely with green beads) represents autism as emerging from a single, uniform polygenic etiology. In this model, earlier and later diagnosed individuals share the same fundamental genetic basis, with later diagnoses potentially resulting from having fewer autism-associated genetic variants or from non-genetic factors such as limited access to diagnostic services.

A special case of the Unitary Model (similar to the second glass, half green beads and half mixed colors) suggests that autism's genetic architecture comprises a core autism polygenic component plus various other mental health-related polygenic factors. In this Model, the same set of genetic variants underlie earlier and later diagnosed autism. However, in addition to genetic variants associated with autism, later diagnosed autistic individuals have elevated genetic predisposition to mental health conditions. Later diagnosed individuals might either have other additional co-occurring neurodevelopmental or may be misdiagnosed as autistic.

The Developmental Model (represented by the third glass with its three distinct layers) proposes two different genetic architectures for earlier and later diagnosed autism (shown by the green and purple beads), along with additional genetic factors related to other mental health conditions (the mixed-color beads). Unlike Model 2, this model suggests that later diagnosed autism's genetic architecture cannot be fully explained by the combination of the genetic effects of earlier diagnosed autism and other mental health conditions.

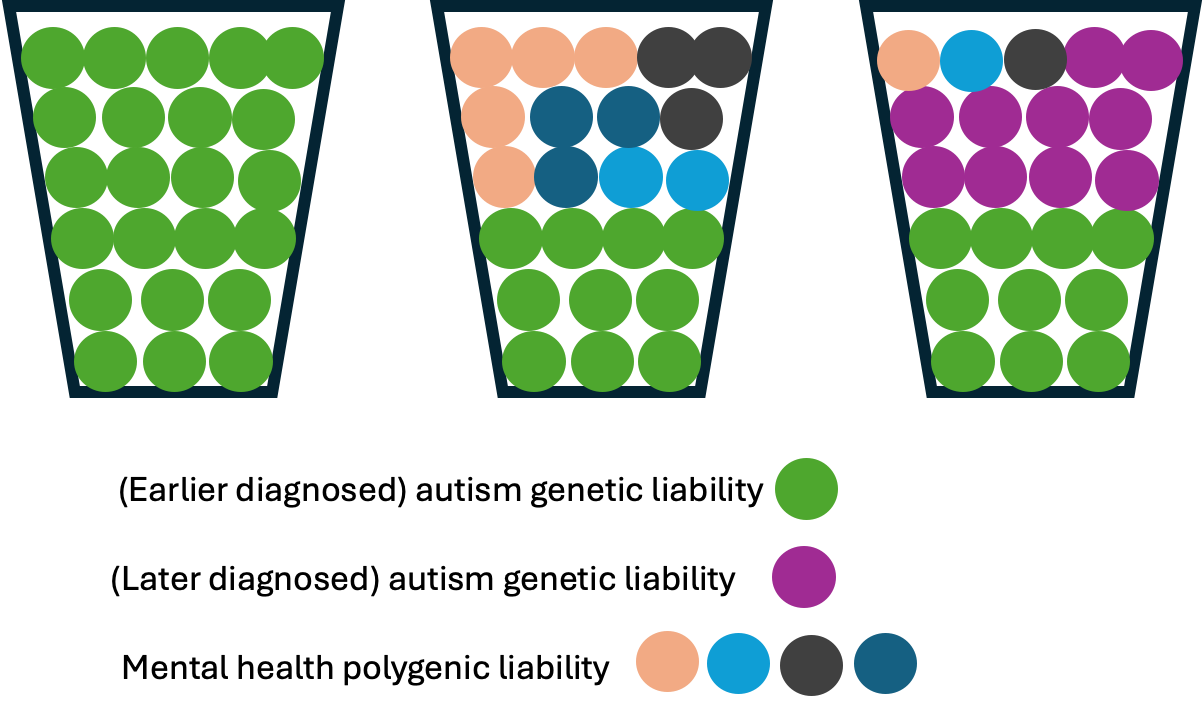

##### Note 8, Figure 1

*Illustrative example of the three models.*

To investigate the appropriateness of each model, we assumed that the PGC-2017 autism GWAS^14^ was a “gold-standard” GWAS of autism as autistic participants in this GWAS typically underwent rigorous assessment using tools such as ADOS^15^ and ADI-R^16^. We further assumed that autistic individuals included in the iPSYCH GWAS consisted of a fraction of autistic individuals with a true autism diagnosis and individuals who were diagnostically misclassified as autistic. If this were true then the total genetic variance of the iPSYCH autism GWAS could be explained by the PGC-2017 autism GWAS and other mental health conditions. In other words, the residual genetic variation would be statistically nonsignificant.

We tested this using genomicSEM after regressing the genetic effects of the iPSYCH GWAS simultaneously on the genetic effects of PGC-2017 autism GWAS^14^, ADHD^17^, depression^18^, schizophrenia^19^, bipolar disorder^20^, anorexia nervosa^21^, and PTSD^22^ (**SEM** **Model 1 in** **Note 8, Figure 2**). The residual genetic variance for iPSYCH autism GWAS was 0.36 (s.e.m = 0.10, P = 2.97x10^-4^). This suggests that just under 40% of the genetic variance in the iPSYCH autism was not explained by either the PGC-2017 autism GWAS or other mental health conditions. In the multiple regression framework within genomicSEM, we observed significant genetic correlation between the iPSYCH autism GWAS and the PGC 2017 autism GWAS (r_g_ = 0.58, s.e.m = 0.12, P = 1.64x10^-6^) and the ADHD GWAS (r_g_ = 0.31, s.e.m = 0.09, P = 1.80x10^-3^). We observed no significant genetic correlation between the iPSYCH autism GWAS and any of the other GWAS. Additionally, the genetic correlation between the iPSYCH and PGC-2017 without conditioning on the other GWAS was r_g_ = 0.61, s.e.m = 0.10, P = 2.64x10^-9^. The genetic correlation between the PGC-2017 and iPSYCH autism GWAS were statistically similar before and after conditioning on the genetic effects of ADHD and other mental health phenotypes.

We obtained consistent results when using the SPARK_before6_ autism GWAS instead of the PGC 2017 GWAS (**SEM** **Model 2 in Note 8, Figure 2**). Across Models 1 and 2, this suggests that the genetic architecture of autism captured in the iPSYCH cohort contains components that are distinct from both early-diagnosed autism and other psychiatric conditions.

In line with our findings, we hypothesised that this residual genetic variance likely represents unique genetic factors associated with later-diagnosed autism presentations. We test this by including SPARK_after10_ autism GWAS in the models as a measure of later diagnosed autism, and PGC 2017 (**SEM Model 3 in Note 8, Figure 2**) or SPARK_before6_ (**SEM Model 4 in Note 8, Figure 2**) as a measure of earlier diagnosed autism. In both models, we find that SPARK_after10_ significantly explains the residual variance in the iPSYCH autism GWAS (P = 0.0193 and P = 0.0365 for PGC and SPARK_before6_ models respectively). This confirms our hypothesis that the previously identified residual genetic variance is indeed associated with later-diagnosed autism presentations. Notably, in both models, ADHD genetic effects now fail to reach statistical significance (P = 0.620 and P = 0.939) after including SPARK_after10_, suggesting that the genetic architecture of later-diagnosed autism is not entirely explained by ADHD. The genetic effects of other psychiatric conditions also remain non-significant. These findings provide strong evidence for distinct genetic architectures between early and later-diagnosed autism, and that later autism diagnosis cannot be fully explained by the additive genetic effects of earlier diagnosed autism and other neurodevelopmental and mental health conditions.

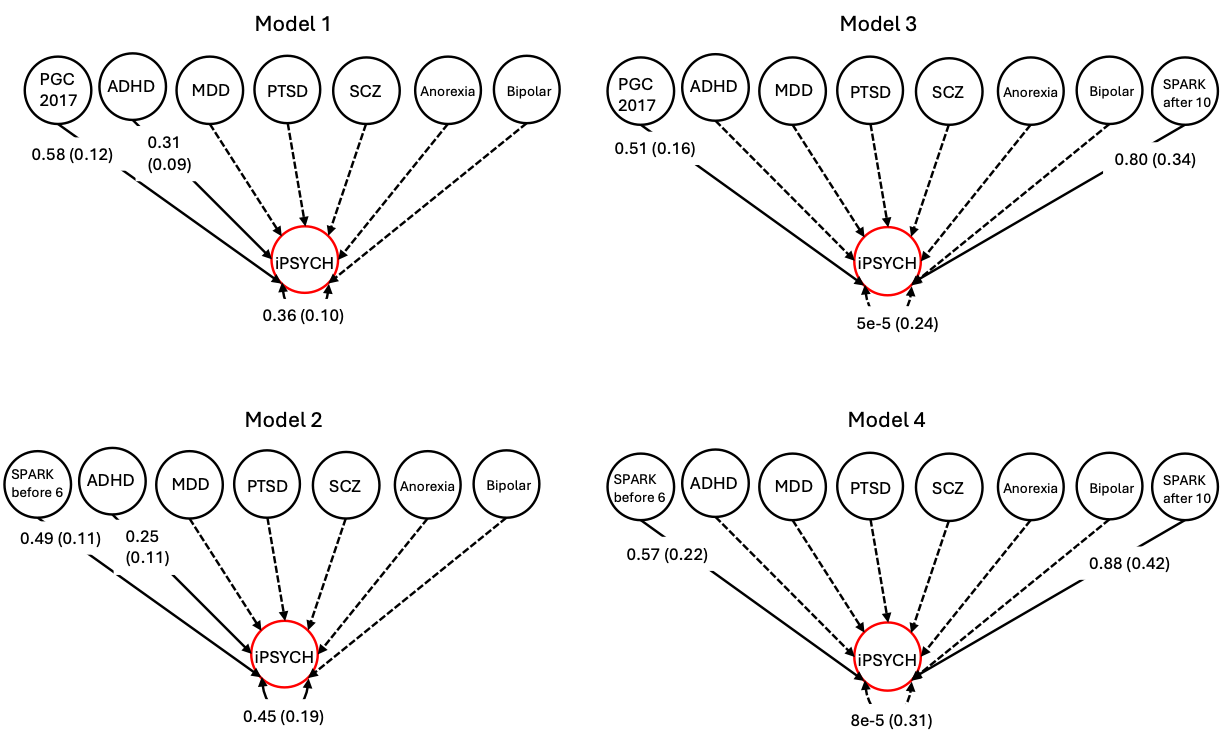

##### Note 8, Figure 2

*Path diagrams representing results from genomic multiple regression analyses using genomicSEM. The genetic effects of the iPSYCH autism GWAS were regressed on the genetic effects of the PGC-2017 autism GWAS (SEM Models 1 and 2) or SPARK_before6_ autism GWAS (SEM Models 2 and 4), attention-deficit/hyperactivity disorder (ADHD), major depressive disorder (MDD), posttraumatic stress disorder (PTSD), schizophrenia (SCZ), anorexia nervosa (Anorexia), and bipolar disorder (Bipolar) simultaneously. In Models 3 and 4, we additionally included SPARK_after10_ GWAS. Single-headed arrows indicate conditional genetic associations between the explanatory variables and the iPSYCH autism GWAS. Numbers represent standardised correlation coefficients, with standard errors in parentheses. Genetic associations between explanatory factors were accounted for in the analyses but are not shown on the graph for simplicity. The two-headed arrows connecting the genetic component of the iPSYCH autism GWAS to itself represent the residual genetic variance unexplained by the genetic influence of any of the GWAS included in the black circles. Solid lines indicate significant genetic associations, while dashed lines indicate non-significant associations. For simplicity, only the significant estimates and standard errors are shown, with the exception of the residual variance (and standard errors).*

### Note 9: Earlier and later diagnosed autism are differently associated with developmental trajectories

As delays in developmental milestones are the earliest indicator that a child may be autistic^23^, we investigated the genetic correlation between developmental traits and the two age-at-diagnosis-related autism polygenic factors. We tested the genetic correlation with: (1) GWASs of developmental phenotypes measured at age 3 in the Norwegian Mother, Father and Child Cohort (N = 41,708–58,630)^24^; (2) GWASs of infant and toddler vocabulary size, meta-analysed from multiple cohorts (N = 6,291–19,296)^25^; (3) GWAS of age at onset of walking, meta-analysed from multiple cohorts (N = 70,560)^26^; and (4) GWAS of rare neurodevelopmental conditions (N = 32,952)^27^.

Both earlier and later diagnosed autism factors were significantly genetically correlated with greater difficulties in social behaviour at age three (**Note 9,** **Figure 1A, Supplementary Table 20**) and the genetic correlations did not statistically differ between the polygenic factors. The genetic correlations of the two factors did differ with age at onset of walking and size of expressive vocabulary at 2 - 3 years: Earlier diagnosed autism factor was genetically correlated with later age at onset of walking and smaller vocabulary.

We extended these findings by investigating the associations between polygenic scores (PGS) derived from GWAS of autism in iPSYCH stratified by age at diagnosis and social and communication difficulties measured at 15 months in ALSPAC. As mentioned in **Supplementary Note 7**, we used PGS derived from iPSYCH_before11_ and iPSYCH_after10_ as indices of earlier and later diagnosed autism respectively. We chose to use PGS generated from these two GWAS over PGS generated from the two autism polygenic factors because the iPSYCH cohort is independent of SPARK, in which we run several sensitivity analyses. For consistency, we used PGS generated from iPSYCH_before11_ and iPSYCH_after10_ in other cohorts as well.

Supporting the genetic correlation findings with expressive vocabulary at 2 - 3 years, PGS for iPSYCH_before11_ but not iPSYCH_after10_ GWAS was associated with greater difficulties in social communication (gestures) at 15 months (**Supplementary Table 21, Note 9, Figure 1B**). However, the estimates did not statistically differ between the two PGS.

Finally, there was no association between either PGS and later attainment of developmental milestones among autistic individuals, although this may be due to collider bias as carriers of rare genetic variants within the cohort have lower autism PGS and substantially delayed developmental milestones^12^ (**Supplementary Table 22**).

Given the cross-sectional nature of the above analyses, we further investigated whether genetics from age-stratified GWAS supported trajectory modelling findings in the general population using the MCS cohort (N = 6,142 - 5,135) and ALSPAC (N = 7,172 - 4,977). We conducted multivariate linear mixed effect models to investigate if age-by-PGS interaction showed significant effects on the SDQ total difficulties and subscale scores. In both cohorts, the PGS*age interaction effects on SDQ total difficulties scores and peer relationship problems significantly differed between the two PGS (**Note 9, Figure 1C, Note 9, Figure 2, and Supplementary Table 23**). In MCS, the effect of iPSYCH_after10_ PGS increased with age for emotional symptoms, peer relationship problems and SDQ total difficulties scores, and decreased with age for prosocial behaviour. In ALSPAC, the effects of iPSYCH_before11_ on hyperactivity/inattention decreased with age, whilst the effect of iPSYCH_after10_ PGS on SDQ total difficulties scores and peer relationship problems nominally increased with age. These differences between the two cohorts are likely due to secular trends in mental health trajectories^28^ or variations in the developmental periods analysed rather than ascertainment differences, as entropy balancing did not alter these findings.

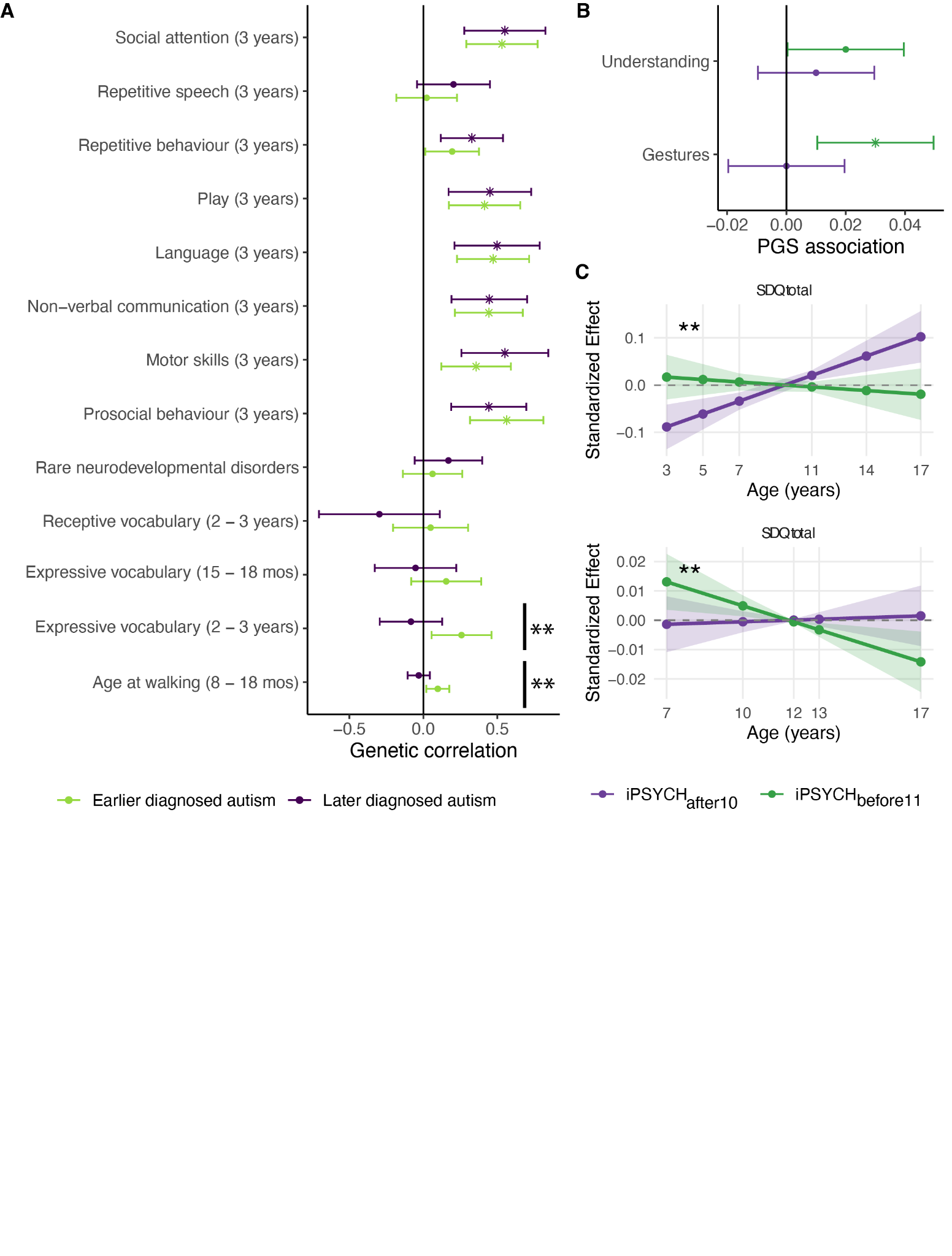

##### Note 9, Figure 1

*Association between age at diagnosis stratified autism and developmental milestones and trajectories. A. Genetic correlation between earlier and later diagnosed autism genetic factors and a range of developmental phenotypes. mos = months. B. Association between iPSYCH_before11_ PGS and iPSYCH_after10_ PGS and social communication skills at 15 months in ALSPAC. For A and B, points indicate the estimate, whiskers indicate 95% confidence intervals, and points with an asterisk (*) indicates significant associations after Benjamini-Yekutieli adjustment. Positive values in both plots A and B indicate greater difficulties/delays. C. Predicted interaction effects between age and autism PGS on SDQ total difficulties scores, derived from linear mixed effects models. Top panel shows results from the Millennium Cohort Study (MCS, ages 3-17 years, born 2000 - 2001) and bottom panel shows results from the Avon Longitudinal Study of Parents and Children (ALSPAC, ages 7-17 years, born 1991 - 1992). Lines represent the predicted age-dependent effects of the PGS with 95% confidence intervals shown as shaded regions. Points indicate ages at which SDQ was measured. Effects are standardized, representing predicted standard deviation changes in SDQ scores per standard deviation increase in PGS at different ages. The dashed horizontal line at zero represents no age-dependent effect. Double asterisk (**) indicates significant difference in genetic correlation between earlier and later diagnosed autism (Plot A), or significant difference in age interaction effects between the two PGS (Plot B).*

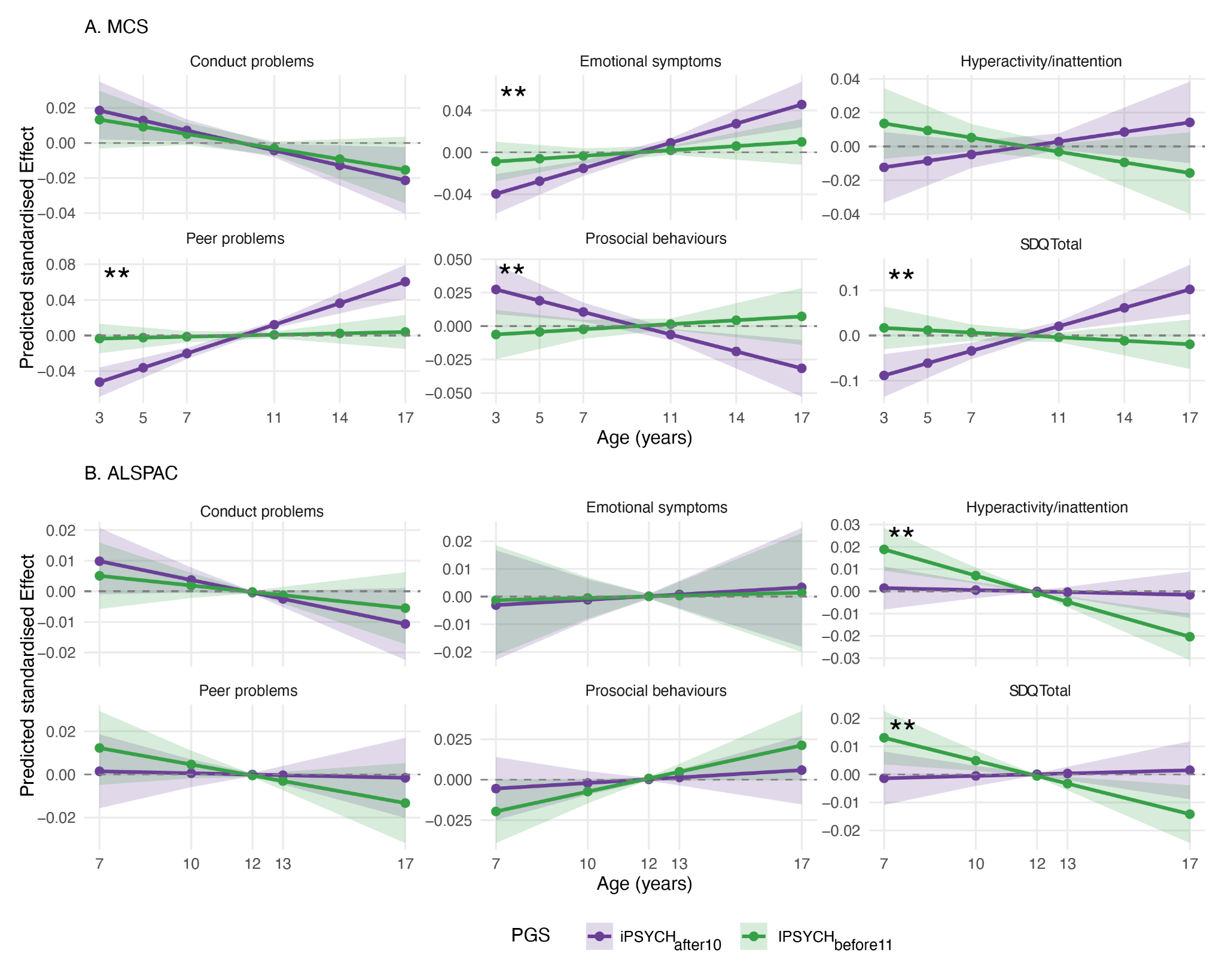

##### Note 9, Figure 2

*Predicted age-dependent effects of autism polygenic scores (PGS) on child behavioral problems across development in two British birth cohorts. The plots show the predicted interaction effects between age and autism PGS on SDQ total difficulties and subscale scores, derived from linear mixed effects models. Panel A shows results from the Millennium Cohort Study (MCS, ages 3-17 years) and Panel B shows results from the Avon Longitudinal Study of Parents and Children (ALSPAC, ages 7-17 years). Lines represent the predicted age-dependent effects of two versions of autism PGS (iPSYCH_after10_ and iPSYCH_before11_) with 95% confidence intervals shown as shaded regions. Points indicate ages at which SDQ was measured. Effects are standardised, representing predicted standard deviation changes in SDQ scores per standard deviation increase in PGS at different ages. The dashed horizontal line at zero represents no age-dependent effect. Two asterisks (**) in the top left of each plot indicates interaction effects that are significantly different between the two sets of PGS (P < 0.05, two-tailed Z test).*

### Note 10: Impact of various demographic and clinical characteristics on age at autism diagnosis

Age at autism diagnosis is complex and likely impacted by several socio-biological factors that vary across time and geography. Some of these factors can confound our results. We assessed the impact of five factors on our findings. These include: 1. Changes in diagnostic criteria over time; 2. Co-occurring intellectual disability and developmental delays in the child; 3. Country-level differences in diagnostic practices; 4. Sex; and 5. Parental factors influencing age at autism diagnosis.

Our primary aim was to investigate whether genetic and longitudinal results we obtained primarily reflect these five factors. We do not exclude the possibility of these factors impacting age at autism diagnosis through other mechanisms.

#### 1. Clinical factors: Changes in diagnostic criteria over time

The following cohorts primarily used an ICD based system for diagnosis: iPSYCH, MCS, GUI. The following cohorts primarily used a DSM based system for diagnosis: LSAC-B, LSAC-K, SPARK.

Details of when the birth cohorts were collected and the diagnostic changes that affect them are provided in **Supplementary Table 2**.

We considered the impact of the following changes to the diagnostic criteria in our analyses.

(1) DSM - III (1980) to DSM - IV (1994)

- Does not impact any of the longitudinal analyses as all the participants in the birth cohorts were born after the introduction of DSM - IV.
- To minimise the impact on the genetic analyses in SPARK^29^ we restricted it to autistic individuals who were 22 years or younger, as these autistic individuals were born after 1994 (diagnosed using DSM - IV or DSM5).

(2) ICD - 9 (1977) to ICD - 10 (1994)

- Does not impact any of the longitudinal analyses as all the birth cohorts were born after the introduction of ICD - 10.
- To minimise the impact on the genetic analyses, in iPSYCH^30^, we ran sensitivity analyses by conducting GWAS of autism diagnosed before age 9 (iPSYCH_before9_) and after age 12 (iPSYCH_after11_), and after restricting it to individuals born in 1994 or after. We then compared these results to the original GWAS (iPSYCH_before9_ and iPSYCH_after11_) that included all birth years. The genetic correlation between the GWAS with and without individuals born before 1994 was high (r_g_ > 0.95) and not statistically different from 1, indicating strong consistency in the genetic findings regardless of birth year restrictions.

(3) DSM - IV (1994) to DSM - 5 (2013)

- Affects diagnosis in the Australian cohorts (LSAC-K and LSAC-B). However, it does not affect diagnosis in MCS or GUI from the UK and Ireland respectively, where ICD was predominantly used. We observe similar results between the Australian cohorts and cohorts in Ireland and UK, suggesting that the results observed are not primarily due to changes in the diagnostic criteria, or differences between DSM and ICD.
- Does not affect iPSYCH analyses. However, in SPARK, approximately 60% of the participants included in the study were diagnosed after 2013, implying that they were diagnosed using the DSM-5 criteria. The remaining 40% were diagnosed using the DSM-IV criteria. The PGS for iPSYCH_after10_ was associated with age at autism diagnosis in SPARK even after controlling for the DSM edition (DSM-IV vs DSM-5). Notably, we observed no significant interaction effect between iPSYCH_after10_ PGS and DSM edition, suggesting that iPSYCH_after10_ PGS has similar effects on age at autism diagnosis regardless of which DSM edition was used to diagnose autism.

(4) ICD - 10 (1994 in Denmark) to ICD - 11 (2018)

- This does not impact any of the longitudinal analyses as children in the MCS and GUI cohorts were diagnosed before ICD-11 has been used by the UK and Ireland respectively .
- Does not impact any genetic analyses as the iPSYCH participants were all diagnosed before 2015.

#### 2. Clinical factors: Intellectual disability and co-occurring developmental delays

Several sensitivity analyses indicate that the findings are not driven by co-occurring intellectual disability (ID) and developmental delays.

- Longitudinal analyses: No autistic children in the MCS, GUI, or LSAC cohorts who had measures of cognitive aptitude met the criteria for ID, likely due to participation bias. Furthermore, in regression models, the first principal component of a child's cognitive aptitude does not explain a significant proportion of the variance in age at autism diagnosis, yet again likely due to participation bias and relatively small sample size. In the general literature, the variance explained by IQ/cognitive ability is typically less than 10%.
- Genetic analyses: First, in SPARK, we observed no significant attenuation of the SNP-based heritability in age at autism diagnosis when accounting for co-occurring ID, parent-reported IQ scores, and age at attaining developmental milestones. Second, in SPARK, we find similar over-transmission of the polygenic scores (PGS) for iPSYCH_before11_ and iPSYCH_after10_ among autistic individuals with and without ID. Third, in SPARK, PGS for iPSYCH_after10_ has a similar positive association with age at autism diagnosis even after excluding individuals with ID and limited verbal ability. Fourth, the GWAS for age at autism diagnosis and age at diagnosis stratified autism GWAS were all controlled for ID. Finally, we find no genetic correlation between the earlier and later diagnosed autism genetic factors and a GWAS for rare neurodevelopmental conditions, the majority of whom have co-occurring intellectual disability.

#### 3. Clinical factors: Country/continent level differences in diagnosis

Our analyses indicate that the results do not primarily reflect differences in diagnostic practices across countries.

- Longitudinal analyses: We find similar results in cohorts from the UK, Ireland, and Australia. Notably, the UK uses the ICD, and Australia primarily uses the DSM.
- Genetic analyses: First, the pattern of genetic correlation among different autism GWAS was not explained by country level/cohort differences. Second, we further tested this using genomicSEM. In the genomicSEM analyses, a geography model wherein we tested the hypothesis that cohorts from Europe will load onto one factor and cohorts predominantly from North America will load onto a second factor had poor fit statistics. Third, we find consistent genetic correlation between age at diagnosis stratified GWAS from both SPARK and iPSYCH and other mental health conditions.

#### 4. Demographic factors: Sex

We ran several sensitivity analyses to confirm that the results were not primarily picking up sex differences.

- Longitudinal analyses: First, in the longitudinal analyses of birth cohorts, both Latent Growth Curve Models and Growth Mixture Models conducted only in autistic males identified consistent results to the analyses conducted in both males and females. Second, in the latent growth curve analyses, age of diagnosis stratified models better fit the data compared to sex-stratified models. Finally, in birth cohorts, multiple regression and mediation analyses indicated that trajectories explained a larger proportion of the variance in age at autism diagnosis than sex. This is consistent with findings from wider literature that suggests that sex explains a small proportion of the variance in age at autism diagnosis, as provided in Extended Data Figure 2.
- Genetic analyses: First, we find a significant SNP-based heritability for age at autism diagnosis even after accounting for sex as a covariate. Second, the pattern of genetic correlations among the different autism GWAS was not explained by sex differences. For example, the female-stratified autism GWAS in iPSYCH had higher genetic correlation with the male-stratified autism GWAS (r_g_ = 0.80, s.e.m = 0.08) than with the sex-unstratified PGC-2017 (r_g_ = 0.48, s.e.m = 0.12) or the SPARK autism GWAS (r_g_ = 0.63, s.e.m = 0.17). Finally, the iPSYCH_after10_ PGS was associated with age at autism diagnosis in the SPARK cohort in both males and females.

#### 5. Demographic factors: Parental characteristics

We ran a few analyses to understand if parental characteristics can impact age at autism diagnosis, primarily through gene-environment correlations.

- Longitudinal analyses: In the longitudinal analyses of birth cohorts, accounting for ethnic minority status^31^, parental socio-economic status, material deprivation, and maternal age at birth does not impact the variance explained by the GMM latent trajectories on age at autism diagnosis.
- Genetic analyses: In SPARK, controlling for parental socio-economic status and neighbourhood deprivation does not significantly attenuate the SNP heritability for age at autism diagnosis.

### Supplementary Figure 1

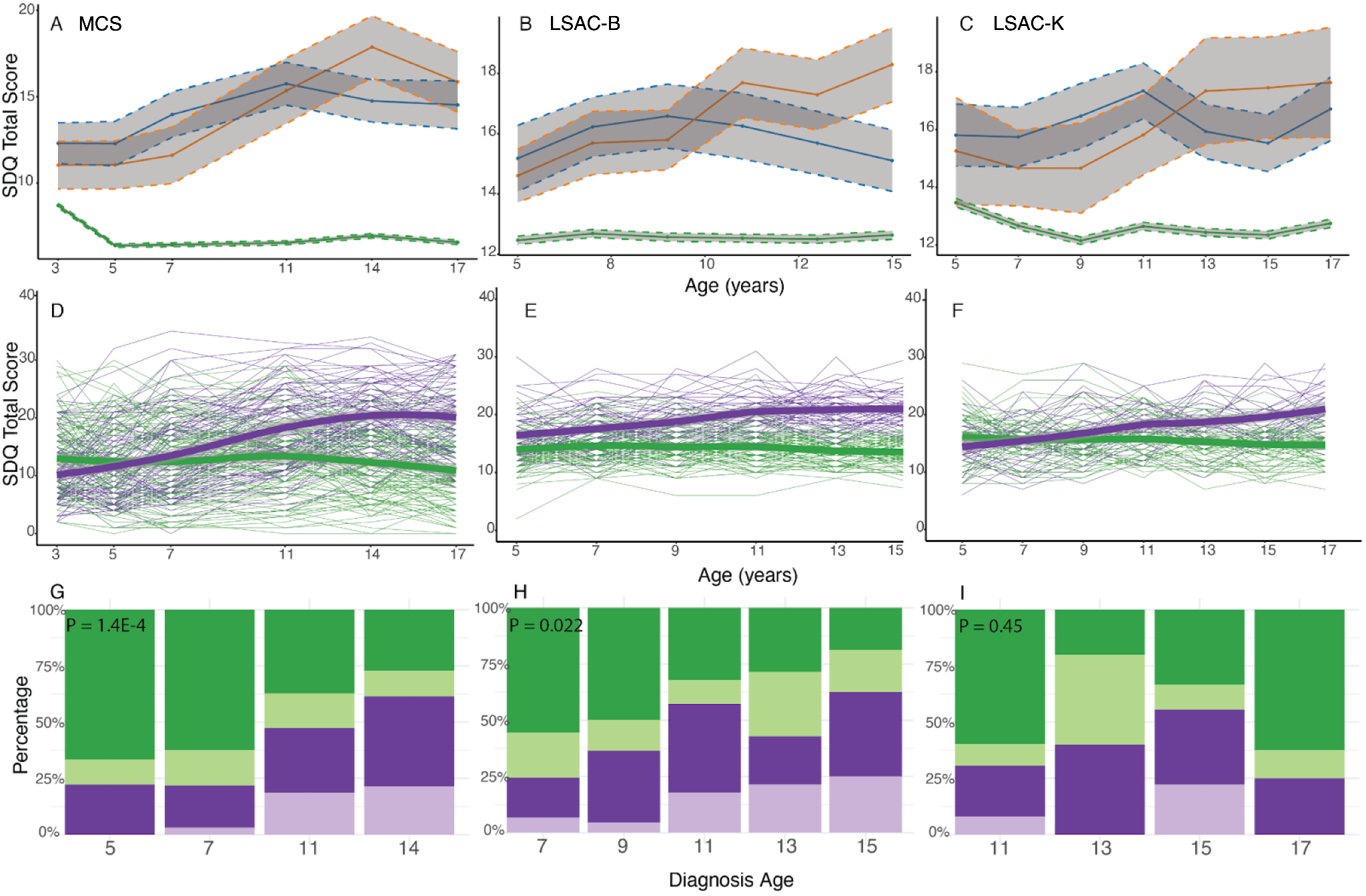

*Mean trajectories of SDQ total difficulties scores by autism diagnosis timing in three birth cohorts. A - C: Mean SDQ total difficulties scores in autistic individuals diagnosed in childhood (blue) and adolescence (orange), and individuals without an autism diagnosis (green) in the MCS (A), LSAC-B (B), and LSAC-K (C) cohorts. Grey regions indicate 95% confidence intervals. For completeness, we also plot the results of GMM in plots D - I. D - F: Longitudinal growth mixture models of SDQ total difficulties scores among autistic individuals, demonstrating the presence of two groups (green indicating early childhood emergent latent trajectory and purple indicating late childhood emergent latent trajectory) in the MCS (D), LSAC-B (E) and LSAC-K (F) cohorts. Shaded areas indicate 95% confidence intervals in D - F. G - I: Stacked bar charts providing the proportion of individuals who had been diagnosed as autistic at specific ages, categorised by membership in the latent trajectories identified from the growth mixture models in MCS (G), LSAC-B (H), and LSAC-K (I) cohorts. Darker colours indicate males and lighter colours indicate females. P-values (inset) are from chi-square tests comparing the distribution of age at autism diagnosis between the two latent trajectories.*

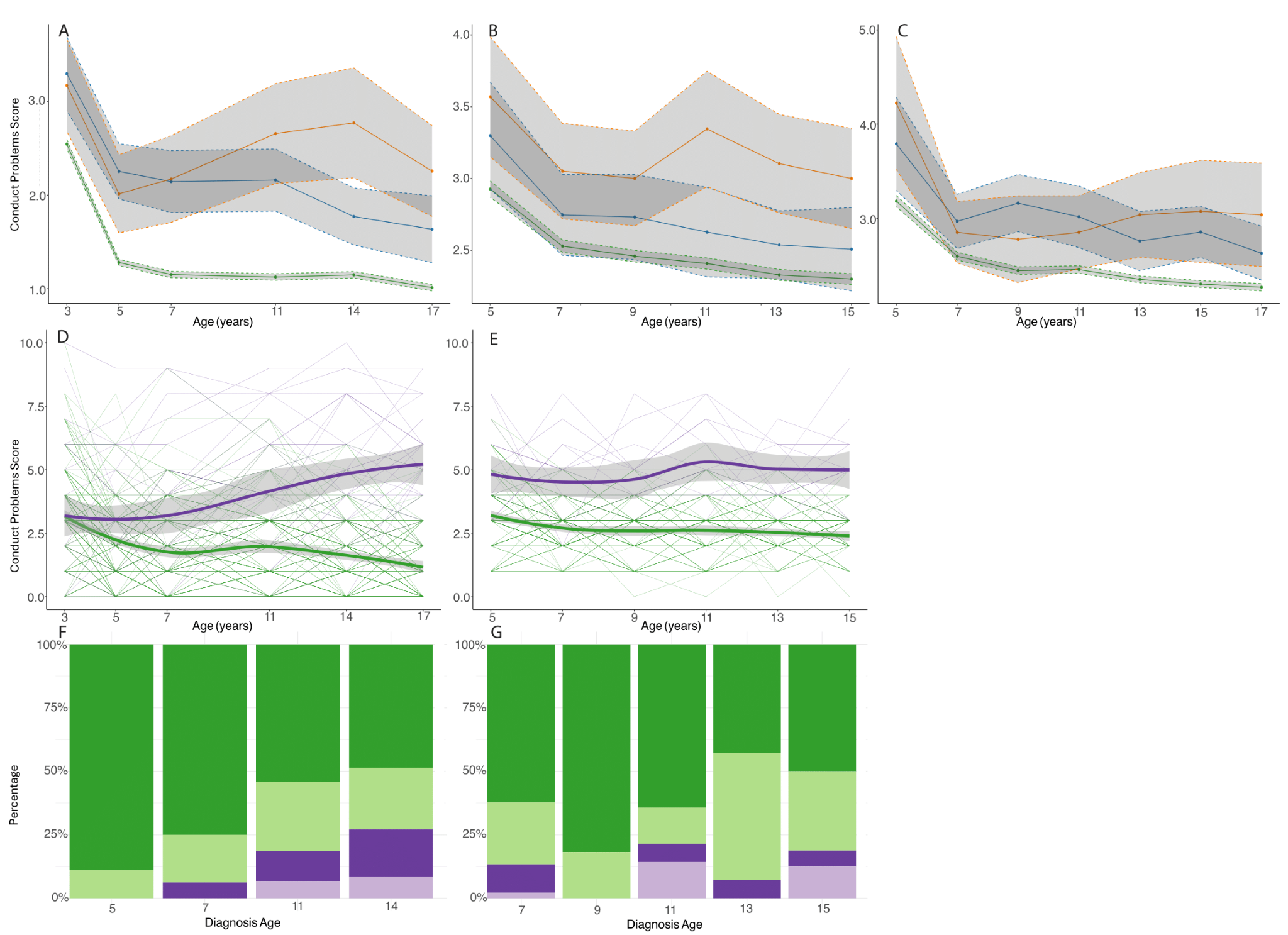

### Supplementary Figure 2

*Trajectory analysis of conduct problems score. A - C: Mean conduct problem scores in autistic individuals diagnosed in childhood (blue) and adolescence (orange), and individuals without an autism diagnosis (green) in the MCS (A), LSAC-B (B), and LSAC-K (C) cohorts. Grey regions indicate 95% confidence intervals. D - E: Longitudinal growth mixture models of conduct problem scores among autistic individuals, demonstrating the presence of two groups (green indicating early childhood emergent latent trajectory and purple indicating late childhood emergent latent trajectory) in the MCS (D), LSAC-B (E). Only a single latent group was identified in LSAC-K and hence not plotted. Shaded areas indicate 95% confidence intervals in D - E. F-G: Stacked bar charts providing proportion of individuals who had been diagnosed as autistic at specific ages by the latent trajectory membership from the growth mixture models in MCS (F) and LSAC-B (G).*

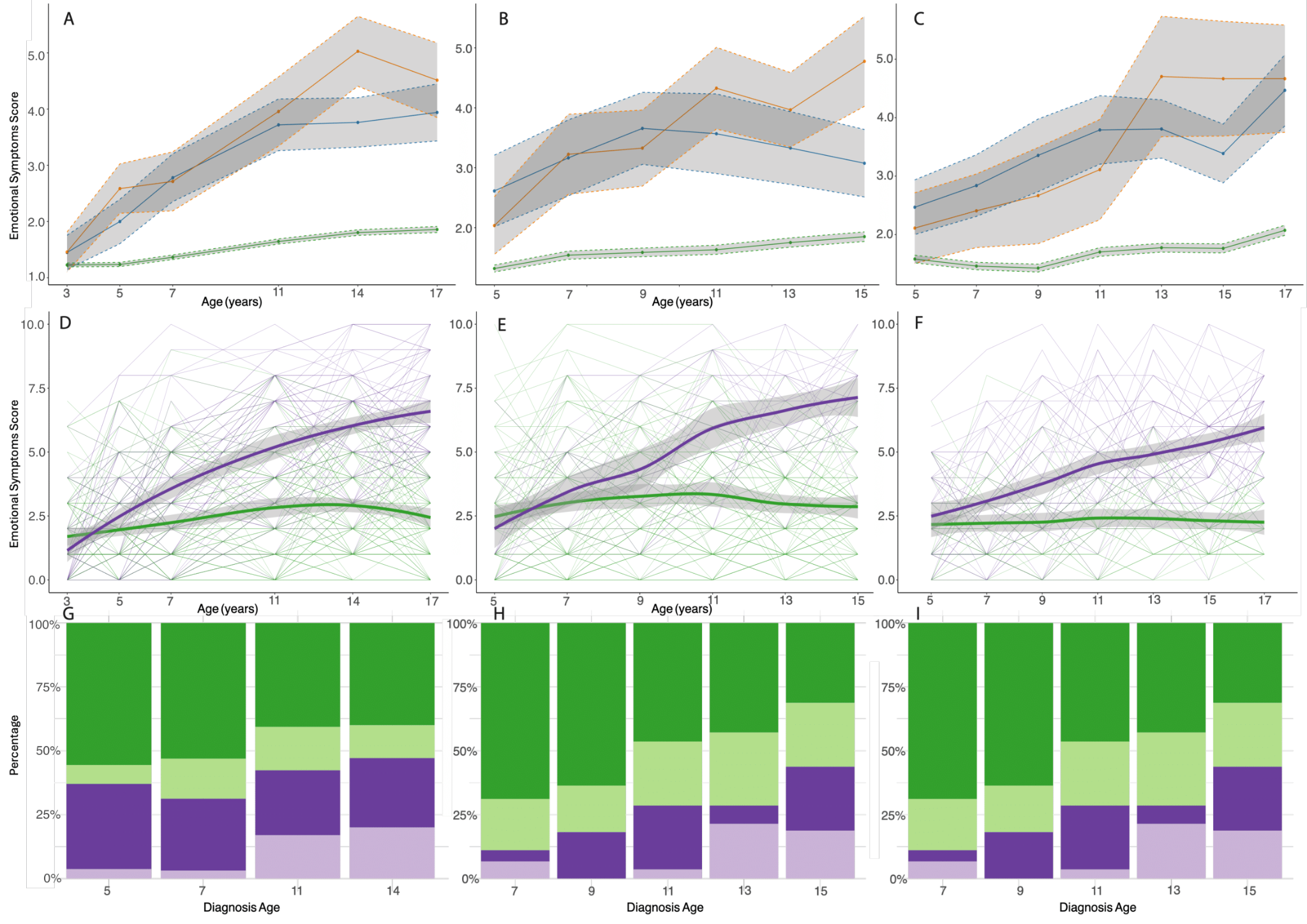

### Supplementary Figure 3

*Trajectory analysis of emotional symptoms score. A - C: Mean emotional symptom scores in autistic individuals diagnosed in childhood (blue) and adolescence (orange), and individuals without an autism diagnosis (green) in the MCS (A), LSAC-B (B), and LSAC-K (C) cohorts. Grey regions indicate 95% confidence intervals. D - F: Longitudinal growth mixture models of emotional symptoms scores among autistic individuals, demonstrating the presence of two groups (green indicating early childhood emergent latent trajectory and purple indicating late childhood emergent latent trajectory) in the MCS (D), LSAC-B (E) and LSAC-K (F) cohorts. Shaded areas indicate 95% confidence intervals in D - F. G - I: Stacked bar charts providing proportion of individuals who had been diagnosed as autistic at specific ages by the latent trajectory membership from the growth mixture models in MCS (G), LSAC-B (H), and LSAC-K (I) cohorts. Darker colours indicate males and lighter colours indicate females.*

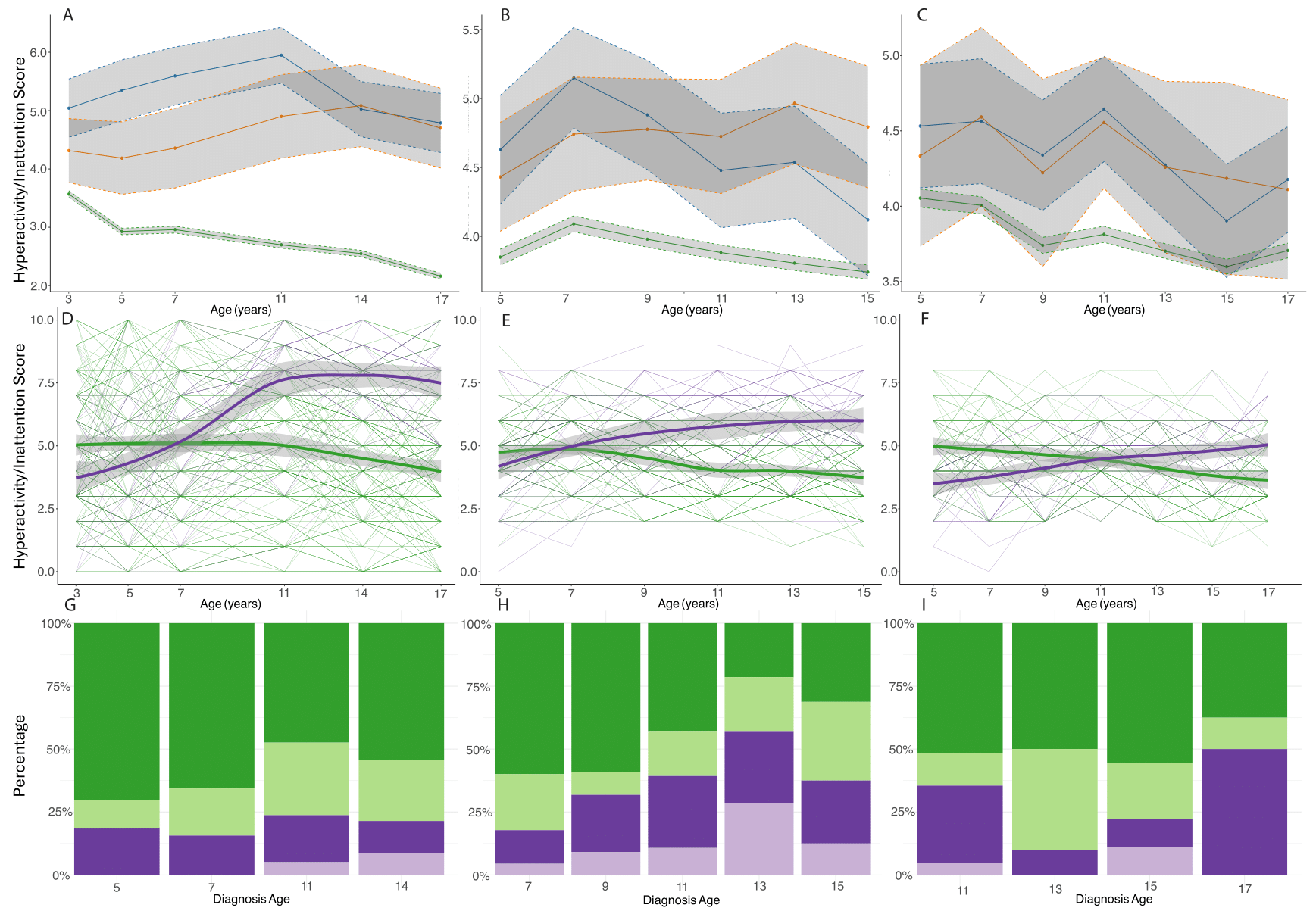

### Supplementary Figure 4

*Trajectory analysis of hyperactivity/Inattention score. A - C: Mean emotional symptom scores in autistic individuals diagnosed in childhood (blue) and adolescence (orange), and individuals without an autism diagnosis (green) in the MCS (A), LSAC-B (B), and LSAC-K (C) cohorts. Grey regions indicate 95% confidence intervals. D - F: Longitudinal growth mixture models of emotional symptoms scores among autistic individuals, demonstrating the presence of two groups (green indicating early childhood emergent latent trajectory and purple indicating late childhood emergent latent trajectory) in the MCS (D), LSAC-B (E) and LSAC-K (F) cohorts. Shaded areas indicate 95% confidence intervals in D - F. G - I: Stacked bar charts providing proportion of individuals who had been diagnosed as autistic at specific ages by the latent trajectories membership from the growth mixture models in MCS (G), LSAC-B (H), and LSAC-K (I) cohorts. Darker colours indicate males and lighter colours indicate females.*

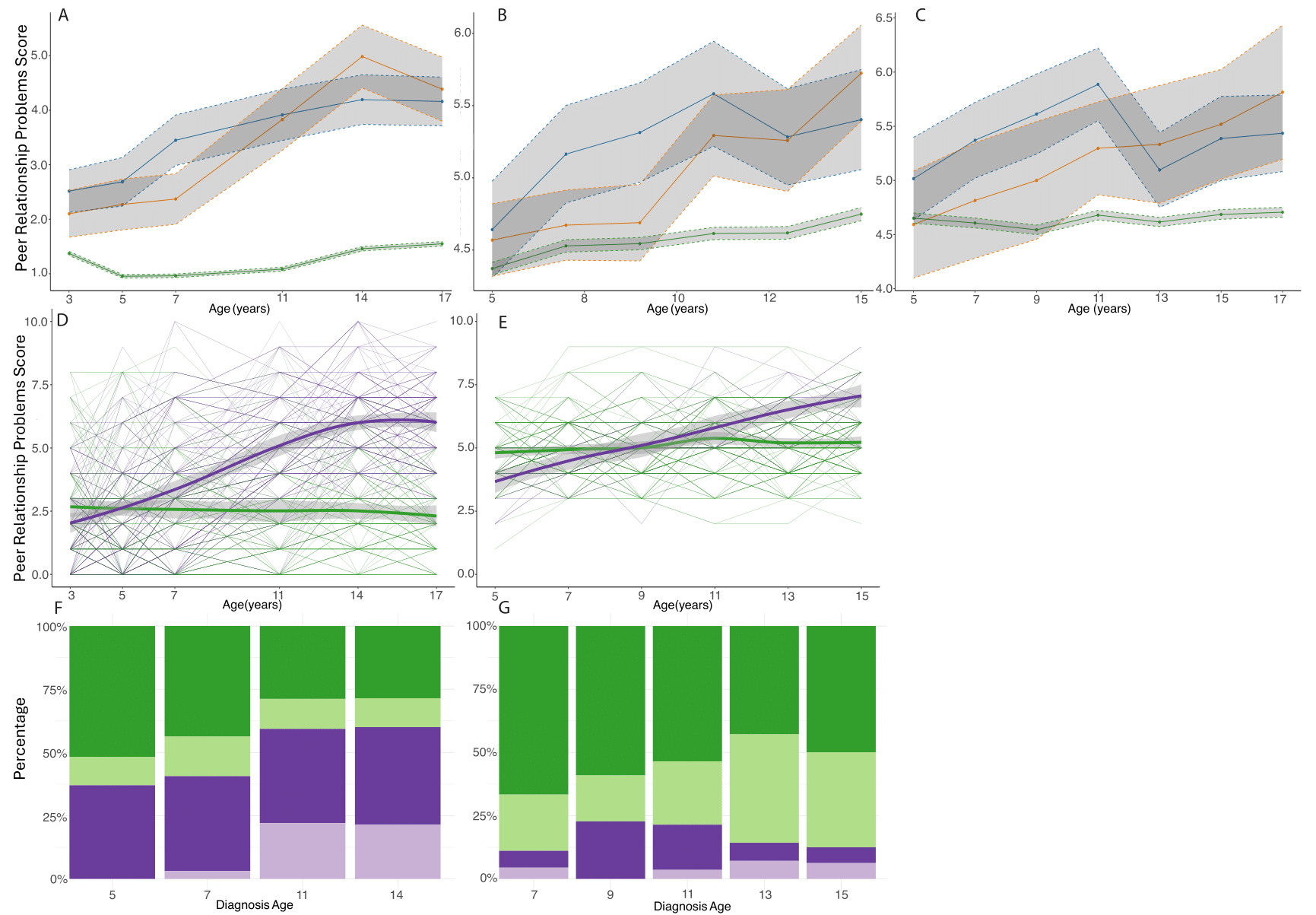

### Supplementary Figure 5

*Trajectory analysis of peer relationship problems score. A - C: Mean peer relationship scores in autistic individuals diagnosed in childhood (blue) and adolescence (orange), and individuals without an autism diagnosis (green) in the MCS (A), LSAC-B (B), and LSAC-K (C) cohorts. Grey regions indicate 95% confidence intervals. D - E: Longitudinal growth mixture models of peer relationship scores among autistic individuals, demonstrating the presence of two groups (green indicating early childhood emergent latent trajectory and purple indicating late childhood emergent latent trajectory) in the MCS (D), LSAC-B (E). Only a single latent group was identified in LSAC-K and hence not plotted. Shaded areas indicate 95% confidence intervals in D - E. F-G: Stacked bar charts providing proportion of individuals who had been diagnosed as autistic at specific ages by the latent trajectory membership from the growth mixture models in MCS (F) and LSAC-B (G).*

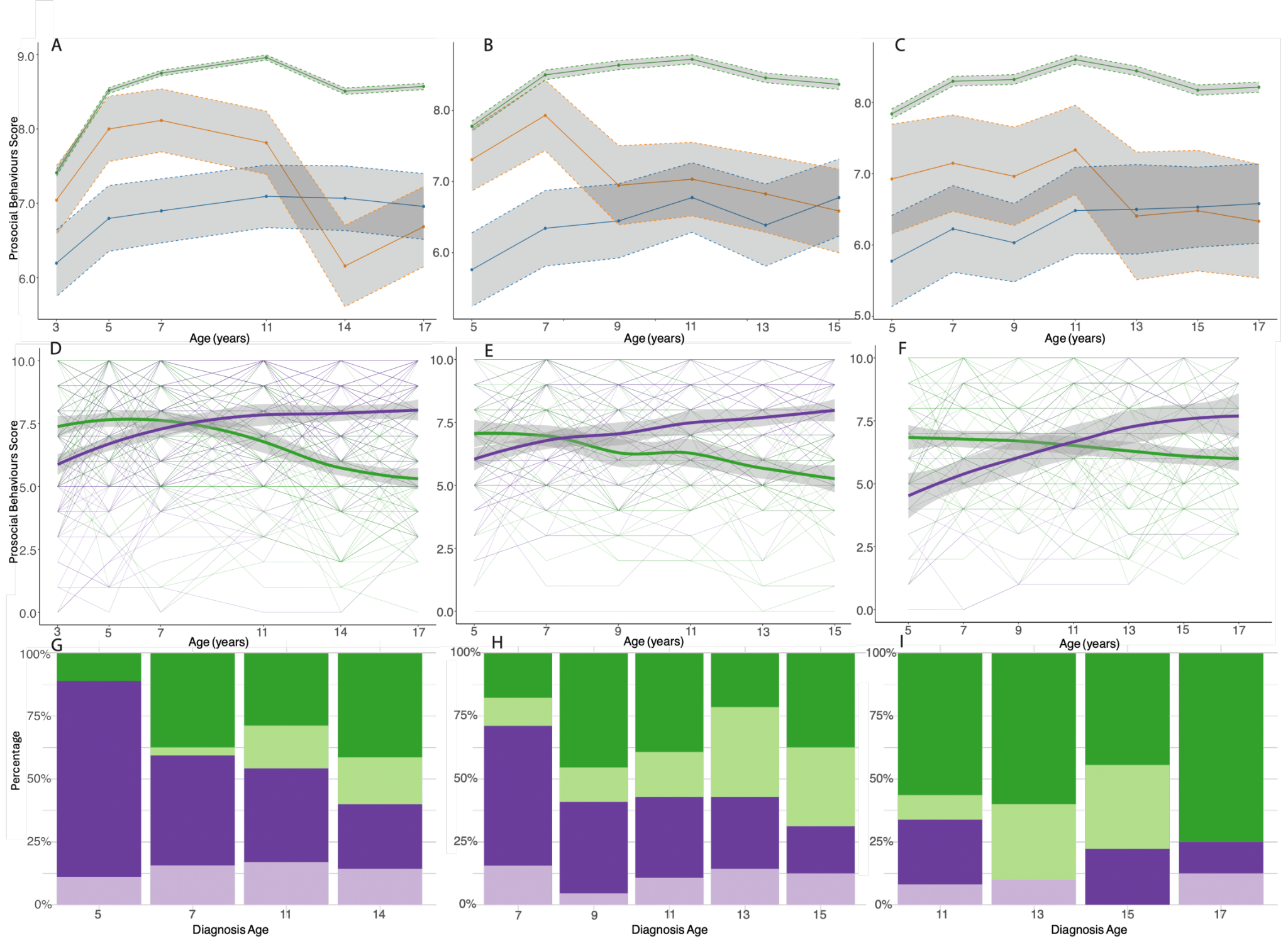

### Supplementary Figure 6

*Trajectory analysis of prosocial behaviour score. A - C: Mean prosocial behaviour scores in autistic individuals diagnosed in childhood (blue) and adolescence (orange), and individuals without an autism diagnosis (green) in the MCS (A), LSAC-B (B), and LSAC-K (C) cohorts. Grey regions indicate 95% confidence intervals. D - F: Longitudinal growth mixture models of prosocial behaviour scores among autistic individuals, demonstrating the presence of two groups (green indicating early childhood emergent latent trajectory and purple indicating late childhood emergent latent trajectory) in the MCS (D), LSAC-B (E) and LSAC-K (F) cohorts. Shaded areas indicate 95% confidence intervals in D - F. G - I: Stacked bar charts providing proportion of individuals who had been diagnosed as autistic at specific ages by the latent trajectory membership from the growth mixture models in MCS (G), LSAC-B (H), and LSAC-K (I) cohorts. Darker colours indicate males and lighter colours indicate females.*

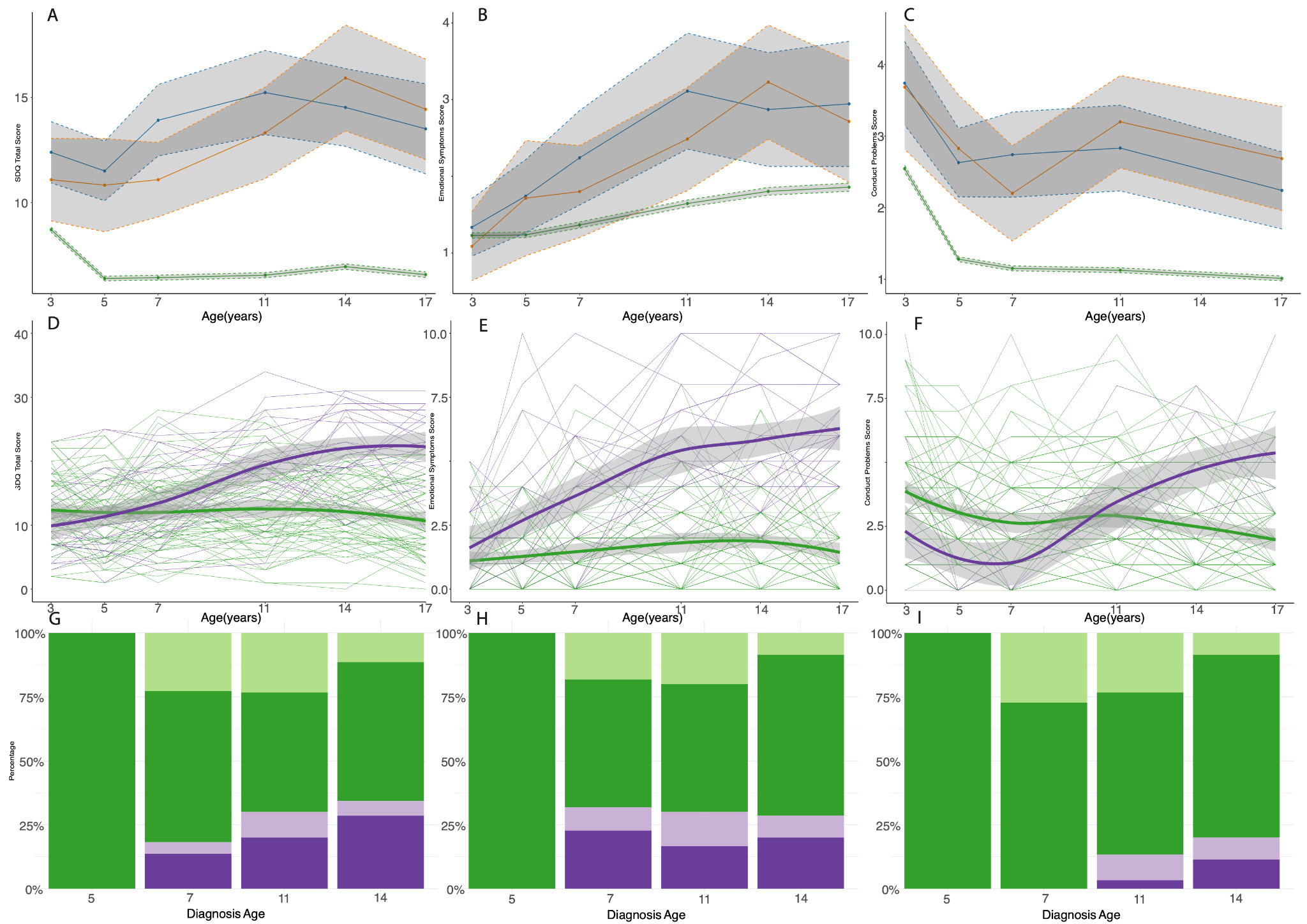

### Supplementary Figure 7

*Sensitivity analyses of trajectory analysis on children with ADHD in MCS. A - C: Mean SDQ total difficulties (A), emotional symptoms (B), and conduct problem (C) scores in individuals diagnosed with ADHD in childhood (blue) and adolescence (orange), and individuals without an ADHD diagnosis (green) in the MCS cohort. Grey regions indicate 95% confidence intervals. D - F: Longitudinal growth mixture models of SDQ total difficulties (D), emotional symptoms (E), and conduct problem (F) scores among individuals with an ADHD diagnosis, demonstrating the presence of two groups (green indicating early childhood emergent latent trajectory and purple indicating late childhood emergent latent trajectory) in the MCS. Shaded areas indicate 95% confidence intervals in D - F. G - I: Stacked bar charts providing proportion of individuals who had been diagnosed with ADHD at specific ages by the latent trajectory membership from the growth mixture models of SDQ total difficulties (G), emotional symptoms (H) and conduct problem (I) scores in the MCS. Darker colours indicate males and lighter colours indicate females.*

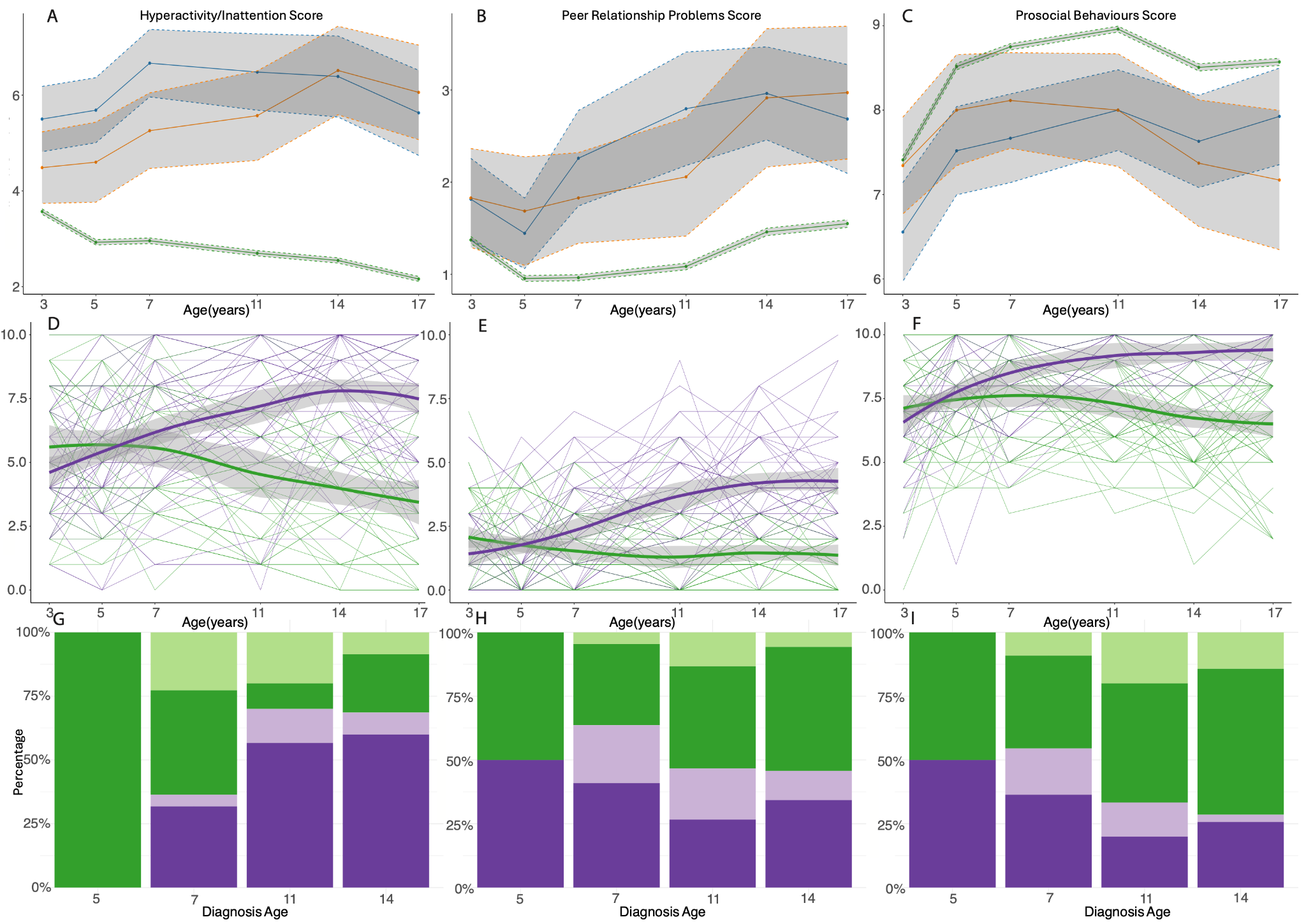

### Supplementary Figure 8

*Sensitivity analyses of trajectory analysis on children with ADHD in MCS (cont.). A - C: Mean hyperactivity/inattention (A), peer relationship problems (B), and prosocial behaviours (C) scores in individuals diagnosed with ADHD in childhood (blue) and adolescence (orange), and individuals without an ADHD diagnosis (green) in the MCS cohort. Grey regions indicate 95% confidence intervals. D - F: Longitudinal growth mixture models of hyperactivity/inattention (D), peer problems (E), and prosocial behaviour (F) scores among individuals with an ADHD diagnosis, demonstrating the presence of two groups (green indicating early childhood emergent latent trajectory and purple indicating late childhood emergent latent trajectory) in the MCS. Shaded areas indicate 95% confidence intervals in D - F. G - J: Stacked bar charts providing proportion of individuals who had been diagnosed with ADHD at specific ages by the latent trajectory membership from the growth mixture models of hyperactivity/inattention (G), peer problems (H) and prosocial behaviour (I) scores in the MCS. Darker colours indicate males and lighter colours indicate females.*

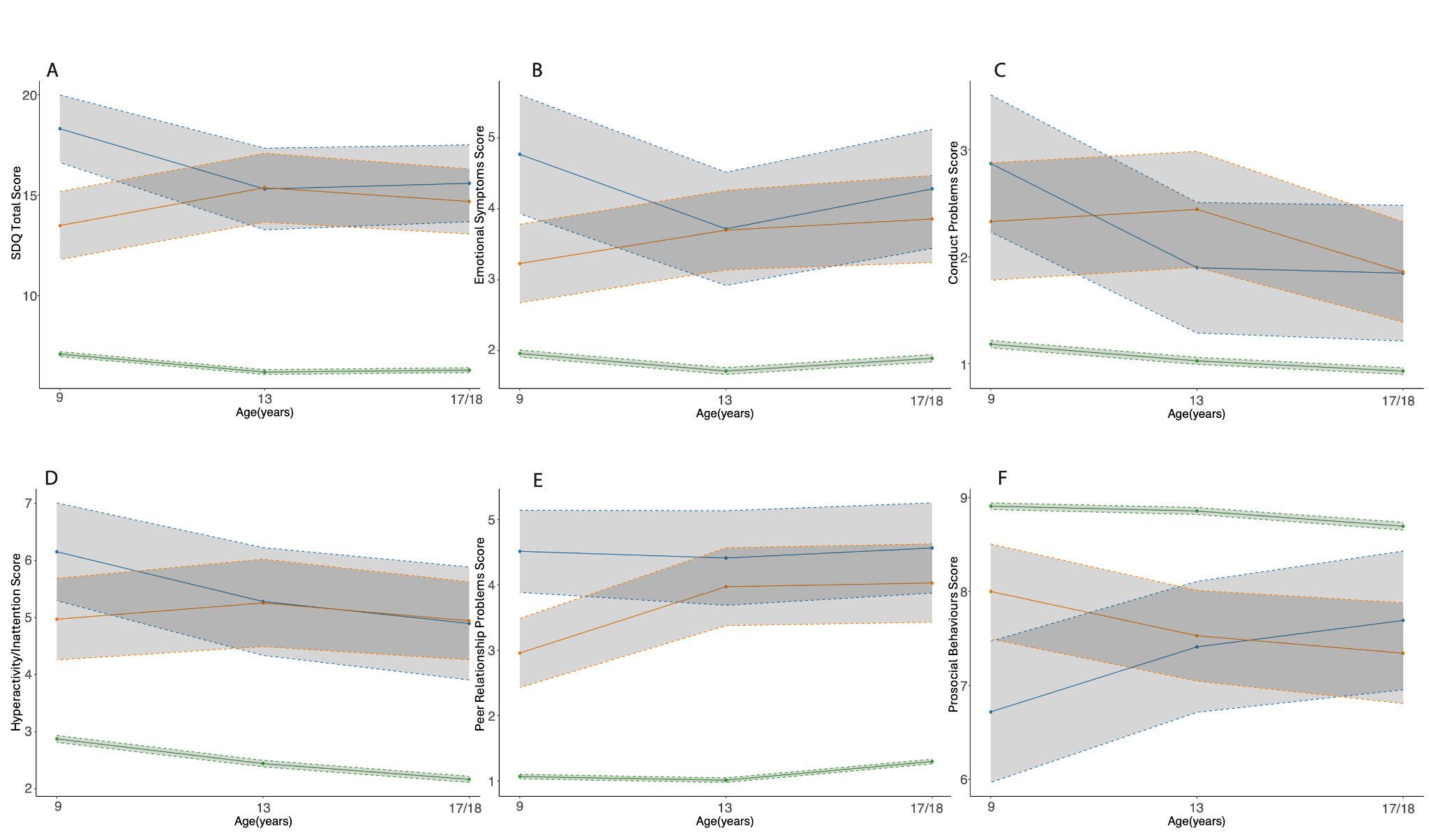

### Supplementary Figure 9

*Mean SDQ trajectories by age at autism diagnosis in GUI. Mean SDQ total difficulties and subscale scores in autistic individuals diagnosed in childhood (blue) and adolescence (orange), and individuals without an autism diagnosis (green) in the GUI cohorts. (A) SDQ total difficulties scores; (B) Emotional symptoms scores; (C) Conduct problem scores; (D) Hyperactivity/inattention scores; (E) Peer problem scores; (F) Prosocial behaviour scores.*

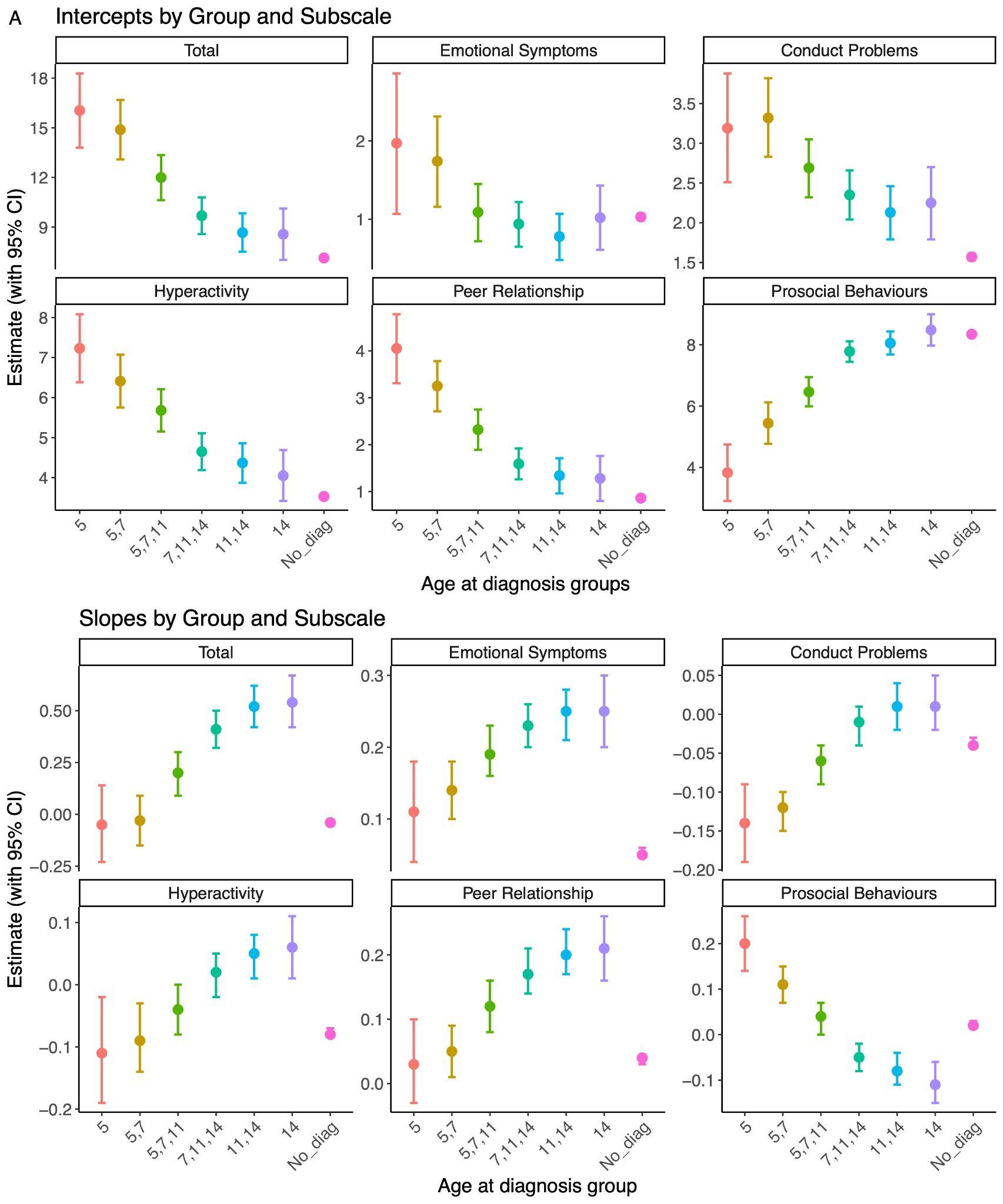

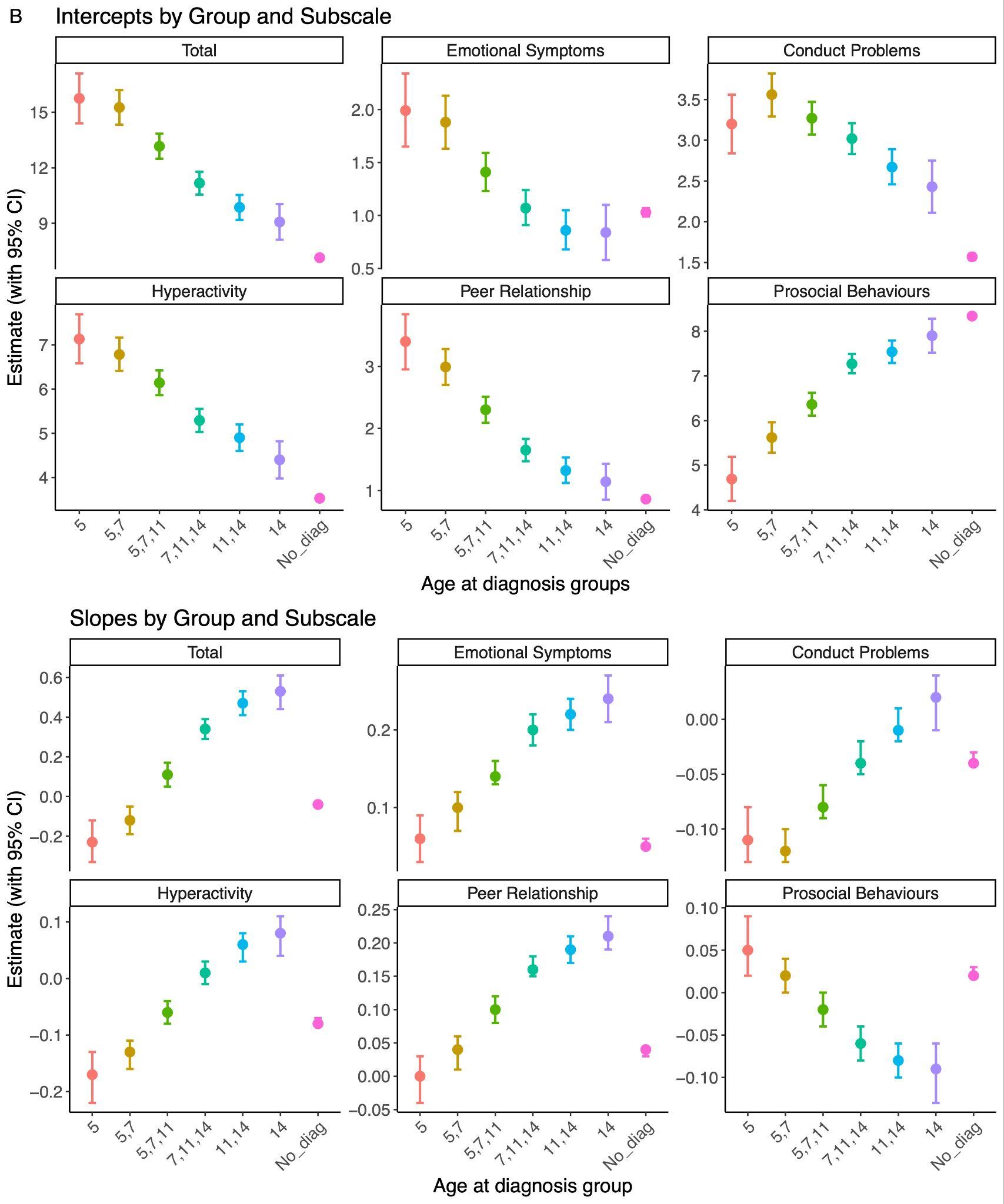

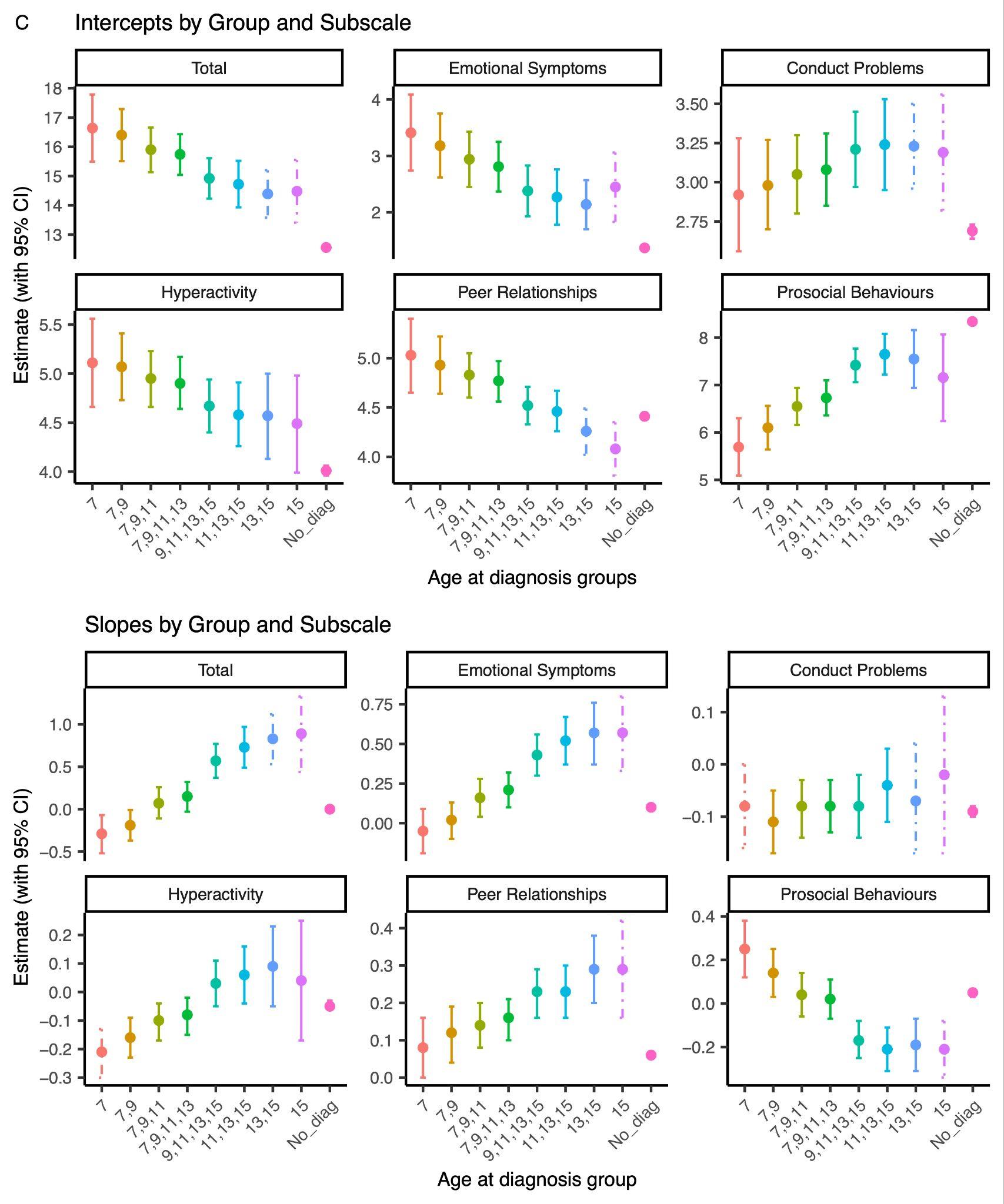

### Supplementary Figure 10

*Latent growth curve model (LGCM) estimates for SDQ total difficulties and subscale scores by age at autism diagnosis group (including a non-autistic comparison group) for: A. MCS core sample; B. MCS imputed autism sample; and C. LSAC-B. For each panel, the* ***intercept*** *(top) and* ***slope*** *(bottom) estimates are presented with 95% confidence intervals. Dashed lines indicate estimates with negative latent variances or non-positive definite latent covariance matrices, likely due to small sample sizes.*

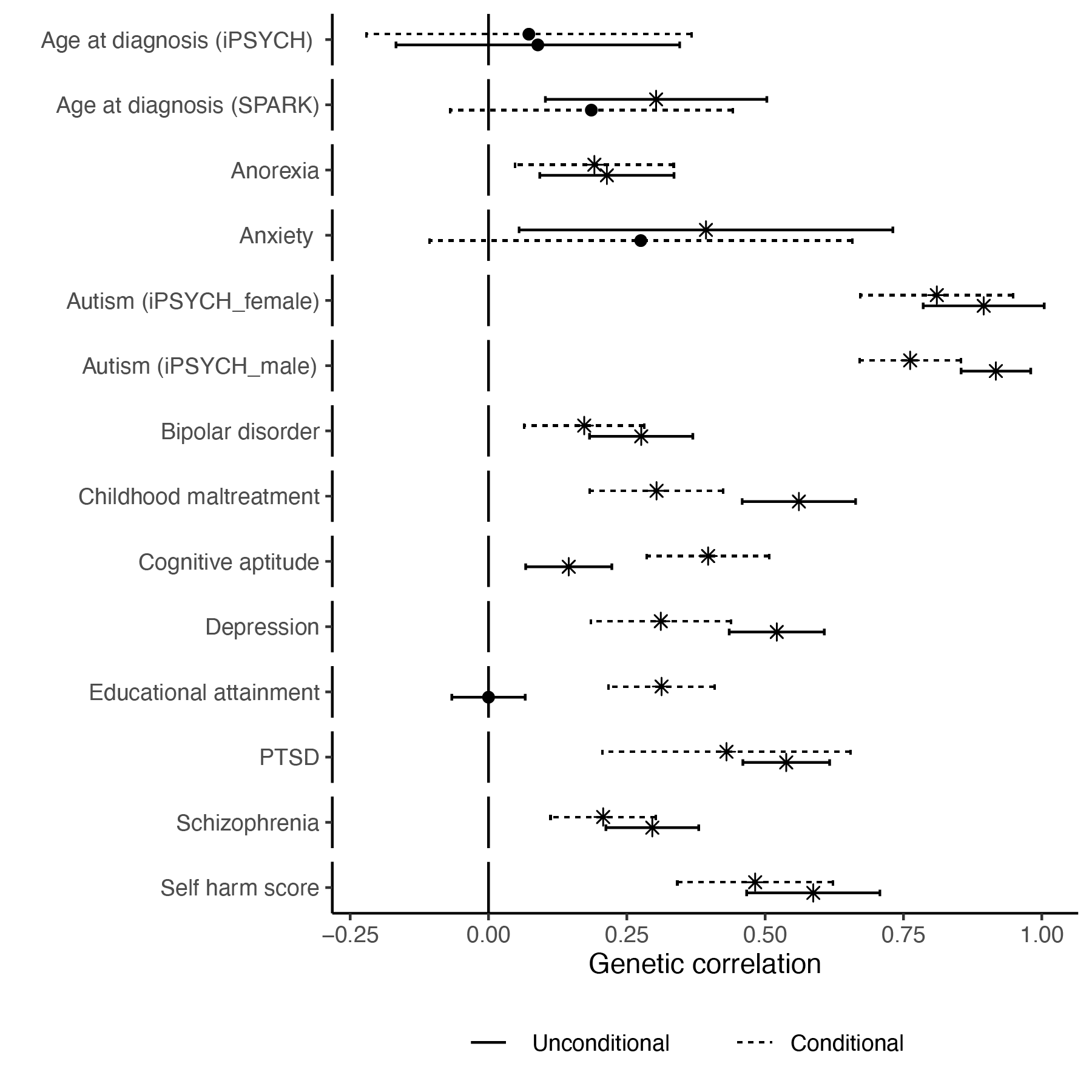

### Supplementary Figure 11

*Genetic correlation between iPSYCH_after10_ autism GWAS and other traits before and after conditioning on the genetic effects of ADHD. Points indicate the estimate, whiskers indicate 95% confidence intervals, and points with an asterisk (*) indicate significant associations after Benjamini-Yekutieli adjustment. Solid lines represent genetic correlation estimates from iPSYCH_after10_ that have not been conditioned on the genetic effects of ADHD. Dotted lines represent genetic correlation estimates from iPSYCH_after10_ after conditioning on the genetic effects of ADHD.*

### Summary

We wanted to understand why some autistic people receive a diagnosis only in late childhood or afterwards. We knew, from previous research, that one reason for variable age at autism diagnosis is because behavioural features that typically lead to an autism diagnosis change from infancy to adolescence.

To better understand how changes in behavioural features can impact when someone receives an autism diagnosis, we analysed data from four long-term studies that followed children from birth through adolescence. In all four studies, children were born around the same time. We measured behavioural features using a parent/caregiver reported questionnaire called the Strengths and Difficulties Questionnaire, measured at multiple time points as the children grew up.

We found that autistic children tended to follow one of two different behavioural trajectories as they grew up. One group showed higher behavioural difficulties from a very young age that remained relatively stable over time. Children in this group were more likely to be diagnosed as autistic at an earlier age.

The other group did not show as many difficulties in early childhood, but began to struggle more with social skills, emotional problems, and peer relationships in late childhood and adolescence. Those in this second group were more commonly diagnosed as autistic later, often in adolescence.

We then looked at genetic factors and found that a person's genetic profile correlates with their age of autism diagnosis. We identified two correlated genetic profiles, or "polygenic factors", associated with autism that seem to correspond to the two behavioural trajectories.

One genetic factor was linked to being diagnosed earlier and having more social communication difficulties in infancy. The other genetic factor was associated with a later autism diagnosis, more emotional and peer problems in adolescence, and higher rates of other conditions like ADHD and trauma.

These findings suggest there are different genetic influences that can predispose some people to show clear autism traits from a very young age, leading to an earlier diagnosis. For others, genetic influences may alter which autism features emerge and when. Some of these children may have features that are not picked up by parents or caregivers until they cause significant distress in late childhood or adolescence.

Our study suggests that the age when someone is diagnosed with autism seems to depend on a combination of their genetic factors influencing developmental trajectories, socio-behavioural challenges emerging at different points, and other factors still to be fully understood. Recognising this diversity and variability in autism can help more timely diagnosis and provide more personalised support for autistic people.

### FAQs

**About the Study**

**Why did you do this study?**

We wanted to understand why some autistic people receive a diagnosis only in late childhood or later. We knew that there is some epidemiological and some qualitative studies on the topic. In parallel, we were also aware of studies that show that social, emotional, and behavioural features change over the course of one's life, especially during childhood and adolescence. Parents and caregivers typically seek an autism diagnosis when they observe social, emotional, and behavioural features in their children that need support. We wondered if changes in these features during early life partly contribute to when someone receives an autism diagnosis. Given that how social, emotional, and behavioural features change over a young person’s life is partly due to their genetics, we also wanted to understand how genetic variants can impact when these features emerge, leading to an autism diagnosis.

Simply put, the model we studied was this:

Genetic variants → Varying developmental trajectories over time → Varying emergence of autism features → Varying age at autism diagnosis

**What did you find?**

The main findings of this study are:

- There are at least two behavioural trajectories that autistic children tend to follow - one group shows elevated social, emotional, and behavioural difficulties from very early childhood, while the other group does not show as many issues until late childhood or adolescence.
- A person's age of being diagnosed with autism is partly explained by genetic factors and their neurodevelopmental trajectories. However, we still do not know many factors that explain when someone receives an autism diagnosis.
- We identified two correlated genetic profiles or "polygenic factors" that correspond to the two behavioural trajectories. One is linked to earlier autism diagnosis, and social communication difficulties in infancy. The other is associated with later diagnosis, increasing emotional/peer problems in adolescence, and higher rates of co-occurring conditions like ADHD and PTSD.

**What are the implications of this study?**

The findings suggest there is diversity in how autism manifests and emerges across childhood and adolescence, influenced by a person's unique genetic makeup. This has implications for:

- Improving our understanding of the diversity and variability seen in autism.
- Recognising that there is no one route to diagnosing autism. Multiple developmental pathways, some of which may fully emerge only later in childhood, can lead to an autism diagnosis.
- The need for personalised support and intervention approaches tailored to different developmental trajectories.
- Considering age of autism diagnosis as a factor in research on sex differences, co-occurring conditions, etc. in autism.

**Why did you run genetic analyses?**

We ran genetic analyses for a few key reasons:

- The behavioural trajectory data suggested that age of autism diagnosis is influenced by developmental factors. Since many developmental traits have a genetic component, we wanted to test whether age of diagnosis itself is partly influenced by their genetics.
- We further sought to understand what specific genetic influences underlying age at autism diagnosis are shared with different behavioural trajectories and developmental milestones in the general population.
- We wanted to understand whether studying the genetics of age at autism diagnosis could help explain some of the heterogeneity and diversity seen in the autism genetics studies, including the differential shared genetics with mental health conditions.
- Finally, genetic methods help to provide additional support to results observed from studying the changes in behavioural trajectories among autistic individuals in four birth cohorts.

**What genetic analyses did you run?**

We used a few different genetic methods. Some of the analyses we ran are:

- Estimating the proportion of the variance in age at autism diagnosis explained by the genetic variants studied. This is called genetic heritability or SNP heritability
- Looking at the shared genetics (called genetic correlation) between age at diagnosis and various mental health conditions. In other words, do the genetic variants that increase or decrease the age at autism diagnosis also increase or decrease the genetic propensity for various mental health conditions?
- Using a method called polygenic scores to create a summed index of the genetic likelihood for autism and test their association with age at autism diagnosis.
- Running a statistical method called genetic structural equation model that looks at the relationship between different genetic studies of autism to identify two correlated genetic factors linked to early vs later diagnosis.
- Analysing very rare genetic variants and their links to age at autism diagnosis.

**What are the limitations of the study?**

Some key limitations of this study are:

- The findings do not fully explain all of the variance in age at diagnosis, suggesting other contributing factors.
- We were unable to measure and study the effect of potentially non-genetic influences such as camouflaging/masking, stigma, healthcare access, waiting lists etc. on age at autism diagnosis.
- Analyses were limited by sample sizes for some datasets therefore we cannot assume that all our results apply to the entire population. For example, the number of autistic individuals in the birth cohorts were around a hundred in each cohort.
- We observed some differences in the variance in age at autism diagnosis explained by the factors studied across cohorts. This suggests that there are cultural and geographic features across cohorts that may impact these findings.
- We still do not fully understand what are the early developmental features that lead to a diagnosis of autism in late childhood/adolescence.
- Although we used the largest genetic datasets to date, it is possible that there may be more than two underlying genetic latent traits that contribute to when features leading to an autism diagnosis emerge, contributing to differences in when an autism diagnosis is made.
- All datasets we used were from developed/western countries. It is unclear if we will find similar findings in countries from other parts of the world.

Although the study does not provide the full picture of the factors that contribute to age at autism diagnosis, it is an important first step towards understanding all the different factors that may impact when someone would receive an autism diagnosis.

**Implications of the study**

**What is the impact of factors like camouflaging, waiting time, stigma on age at autism diagnosis?**

The study did not directly measure the impact of factors like camouflaging autistic traits, delays in assessment/diagnosis waitlists, or stigma on age at autism diagnosis. However, we acknowledge that these likely play a role, in addition to the genetic, demographic, and developmental factors that we have studied.

It is important to note that in this study, genetics explain only about 11% of the total variation in when someone receives an autism diagnosis. Similarly, developmental and demographic factors in this study explained between 10 - 60% of the variance in age at autism diagnosis across cohorts, with considerable variation among the cohorts studied. Taken together, it is clear that there are several other unmeasured factors that contribute to when someone receives an autism diagnosis.

More research is needed to understand how much these social/environmental variables influence the timing of when someone receives an autism diagnosis.

**Are earlier and later diagnosed autism two different types of autism?**

No, our findings do not suggest that earlier and later diagnosed autism are completely distinct conditions. Rather, the findings indicate there are at least two main genetic and developmental profiles. One of these profiles predispose some autistic individuals to showing clear traits from very early childhood leading to earlier diagnosis, while autistic individuals with the second profile may not exhibit prominent characteristics until adolescence resulting in later diagnosis. However, the polygenic factors identified were moderately correlated.

**I am/my child is potentially autistic, what are the implications of this study?**

For individuals who may be autistic or have an autistic child, our findings underscore that there can be diversity in how and when autism traits manifest across development. Some key implications are:

- Closely monitoring a child's social/emotional development is important, as difficulties can potentially emerge later even if not apparent in early years.
- Consider getting an evaluation if difficulties arise - although some of these difficulties may be transient, they may be linked to other difficulties that emerge later on.
- Understanding that a later autism diagnosis does not mean someone's autistic traits are any less valid.
- Different support approaches may be helpful depending on the child's unique profile and trajectory.

**Can you use the findings from this study to diagnose/predict autism?**

No, the findings from this study alone cannot be used to diagnose or definitively predict autism in individuals. The genetic scores and developmental trajectory patterns provide insights at a population level, but there is still substantial variability across individuals. Formal autism diagnostic evaluations by qualified professionals are still required.

**What are some potential misinterpretations of the study?**

Some potential misinterpretations of the study's findings include:

- Assuming all autistic children will follow one of just two rigid trajectories, when there is more variability.
- Families delaying seeking evaluation based on expectation that difficulties may naturally resolve.
- Oversimplifying the results to suggest there are two completely distinct "types" of autism.
- Overemphasising genetics while minimising environmental/social influences on age at autism diagnosis.
- Trying to use genetic analyses to self-diagnose or make predictions at an individual level.
- Assuming that autistic individuals can be subgrouped based on when they were diagnosed. Age at diagnosis is merely a proxy for different emergence of developmental trajectories. Several other factors can influence the age of diagnosis in a person.

### Glossary of terms

*Trajectories*: Trajectories are the different paths or ways that things can develop over time. In this study, we found two different paths for how autistic children's behaviours and social skills changed as they got older.

*Genetic variant*: A genetic variant is a difference in someone's genes or DNA code compared to other people. These small variations can sometimes affect things like a person's traits or health.

*GWAS*: This is a way for scientists to look at all of a person's genes to try and find which genetic variants are linked to certain traits or conditions, like autism.

*Polygenic score*: This is a number that shows how many genetic variants linked to a trait, like autism, a person has. A higher number means more of those variants.

*De novo varian*t: These are new genetic variants that weren't inherited from the person's parents, but appeared newly in a person’s DNA.

*Genetically inferred ancestry*: This refers to using a person's genes to figure out what broad geographic regions their ancestors likely came from a long time ago.

*Latent trait:* This is an unseen characteristic that cannot be directly measured but can be calculated using other correlated measured characteristics.

### Consortia membership

**APEX Consortium Authors & Affiliations**

Simon Baron-Cohen^1^, Carrie Allison^1^, Varun Warrier^1^, Alex Tsompanidis^1^, Deep Adhya, Rosemary Holt^1^, Yuanjun Gu^1^, Xinhe Zhang^1^, Omar Al-Rubaie^1^, Daniel H Geschwind^2^, Ramin Ali Marandi Ghoddousi^2^, Alexander EP Heazell^3^, Jonathan Mill^4^, Alice Franklin^4^, Rosie Bamford^4^ , Matthew E Hurles^5^, Hilary C Martin^5^, Mahmoud Mousa^5^, David H Rowitch^6^ , Kathy K Niakan^7^, Graham J Burton^7^, Tereza Cindrova-Davies^7^, Deepak P Srivastava^8^ , Lucia Dutan-Polit^8^, Adam Pavlinek^8^, Laura Sichlinger^8^, Roland Nagy^8^, Madeline A Lancaster^9^, Jose Gonzalez-Martinez^9^, Tal Biron-Shental^10^, Lidia V Gabis^11- 12^, Dorothea Floris^13- 14^, Richard Bethlehem^15^, Michael V Lombardo^16^, Marcin Radecki^1^, Meng-Chuan Lai^1,17-20^, Yeshaya David Greenberg^1^, Elizabeth Weir^1^, Florina Uzefovsky^21^, Yumnah T Khan^1^, Juan Pablo Del Rio^22^

1. Autism Research Centre, University of Cambridge, UK
2. UCLA, Los Angeles, USA
3. Obstetrics and Director Tommy’s Maternal and Fetal Research Centre, University of Manchester
4. University of Exeter Medical School, College of Medicine & Health, University of Exeter
5. Wellcome Trust Sanger Institute, Hinxton, UK
6. Department of Pediatrics, University of Cambridge
7. Centre for Trophoblast Research, University of Cambridge
8. MRC Centre for Neurodevelopmental Disorders, King’s College London, London
9. MRC Laboratory of Molecular Biology, University of Cambridge
10. Department of Obstetrics and Gynecology, Meir Medical Center, Israel
11. Faculty of Medical & Health Sciences, Tel-Aviv University, Israel
12. Maccabi Healthcare, Israel
13. Department of Psychology, University of Zurich, Switzerland
14. Donders Institute for Brain, Cognition, and Behavior, Radboud University, The Netherlands
15. Department of Psychology, University of Cambridge
16. Laboratory for Autism and Neurodevelopmental Disorders, Center for Neuroscience and Cognitive Systems, Istituto Italiano Di Tecnologia, Rovereto, Italy
17. Margaret and Wallace McCain Centre for Child, Youth & Family Mental Health and Azrieli Adult Neurodevelopmental Centre, Campbell Family Mental Health Research Institute, Centre for Addiction and Mental Health, Toronto, ON, Canada
18. Department of Psychiatry, The Hospital for Sick Children, Toronto, ON, Canada
19. Department of Psychiatry, Temerty Faculty of Medicine, University of Toronto, Toronto, ON, Canada
20. Department of Psychiatry, National Taiwan University Hospital and College of Medicine, Taipei, Taiwan
21. Psychology Department, Ben-Gurion University of the Negev, 1 David Ben-Gurion Blvd, Be’er Sheva, Israel
22. Department of Child and Adolescent Psychiatry and Mental Health, University of Chile

**iPSYCH Autism Consortium Authors & Affiliations**

Anders Borglum^1-3^, Jonas Bybjerg-Grauholm^4^, Jakob Grove^1-3,5^, David M. Hougaard^4^, Ole Mors^6^, Preben Bo Mortensen^7^, Merete Nordentoft^8,9^ & Thomas Werge^9,10^

1. The Lundbeck Foundation Initiative for Integrative Psychiatric Research, iPSYCH, Aarhus, Denmark
2. Center for Genomics and Personalized Medicine (CGPM), Aarhus University, Aarhus, Denmark
3. Department of Biomedicine (Human Genetics) and iSEQ Center, Aarhus University, Aarhus, Denmark
4. Danish Center for Neonatal Screening, Department of Congenital Disorders, Statens Serum Institut, Copenhagen, Denmark
5. Bioinformatics Research Centre, Aarhus University, Aarhus, Denmark
6. Aarhus University Hospital—Psychiatry, Psychosis Research Unit, Aarhus, Denmark
7. NCRR, National Centre for Register-Based Research, Aarhus University, Aarhus, Denmark
8. CORE - Copenhagen Research Center for Mental Health, Mental Health Centre Copenhagen, Copenhagen University Hospital, Copenhagen, Denmark
9. Department of Clinical Medicine, Faculty of Health and Medical Sciences, University of Copenhagen, Copenhagen, Denmark
10. Institute of Biological Psychiatry, Mental Health Services, Copenhagen University Hospital, Roskilde, Denmark
    Thomas Werge

**PGC-PTSD (Freeze 3) Consortium Authors & Affiliations**

Caroline M Nievergelt ^1,2,3,*^, Adam X Maihofer ^1,2,3,*^, Elizabeth G Atkinson ^4^, Chia-Yen Chen ^5^, Karmel W Choi ^6,7^, Jonathan R I Coleman ^8,9^, Nikolaos P Daskalakis ^10,11,12^, Laramie E Duncan ^13^, Renato Polimanti ^14,15^, Cindy Aaronson ^16^, Ananda B Amstadter ^17^, Soren B Andersen ^18^, Ole A Andreassen ^19,20^, Paul A Arbisi ^21,22^, Allison E Ashley-Koch ^23^, S Bryn Austin ^24,25,26^, Esmina Avdibegoviç ^27^, Dragan Babić ^28^, Silviu-Alin Bacanu ^29^, Dewleen G Baker ^1,2,30^, Anthony Batzler ^31^, Jean C Beckham ^32,33,34^, Sintia Belangero ^35,36^, Corina Benjet ^37^, Carisa Bergner ^38^, Linda M Bierer ^39^, Joanna M Biernacka ^31,40^, Laura J Bierut ^41^, Jonathan I Bisson ^42^, Marco P Boks ^43^, Elizabeth A Bolger ^11,44^, Amber Brandolino ^45^, Gerome Breen ^9,46^, Rodrigo Affonseca Bressan ^47,48^, Richard A Bryant ^49^, Angela C Bustamante ^50^, Jonas Bybjerg-Grauholm ^51,52^, Marie Bækvad-Hansen ^51,52^, Anders D Børglum ^52,53,54^, Sigrid Børte ^55,56^, Leah Cahn ^16^, Joseph R Calabrese ^57,58^, Jose Miguel Caldas-de-Almeida ^59^, Chris Chatzinakos ^10,11,60^, Sheraz Cheema ^61^, Sean A P Clouston ^62,63^, Lucía Colodro-Conde ^64^, Brandon J Coombes ^31^, Carlos S Cruz-Fuentes ^65^, Anders M Dale ^66^, Shareefa Dalvie ^67^, Lea K Davis ^68^, Jürgen Deckert ^69^, Douglas L Delahanty ^70^, Michelle F Dennis ^32,33,34^, Frank Desarnaud ^16^, Christopher P DiPietro ^10,60^, Seth G Disner ^71,72^, Anna R Docherty ^73,74^, Katharina Domschke ^75,76^, Grete Dyb ^20,77^, Alma Džubur Kulenović ^78^, Howard J Edenberg ^79,80^, Alexandra Evans ^42^, Chiara Fabbri ^9,81^, Negar Fani ^82^, Lindsay A Farrer ^83,84,85,86,87^, Adriana Feder ^16^, Norah C Feeny ^88^, Janine D Flory ^16^, David Forbes ^89^, Carol E Franz ^1^, Sandro Galea ^90^, Melanie E Garrett ^23^, Bizu Gelaye ^6^, Joel Gelernter ^91,92^, Elbert Geuze ^93,94^, Charles F Gillespie ^82^, Slavina B Goleva ^68,95^, Scott D Gordon ^64^, Aferdita Goçi ^96^, Lana Ruvolo Grasser ^97^, Camila Guindalini ^98^, Magali Haas ^99^, Saskia Hagenaars ^8,9^, Michael A Hauser ^32^, Andrew C Heath ^100^, Sian M J Hemmings ^101,102^, Victor Hesselbrock ^103^, Ian B Hickie ^104^, Kelleigh Hogan ^1,2,3^, David Michael Hougaard ^51,52^, Hailiang Huang ^10,105^, Laura M Huckins ^106^, Kristian Hveem ^55^, Miro Jakovljević ^107^, Arash Javanbakht ^97^, Gregory D Jenkins ^31^, Jessica Johnson ^108^, Ian Jones ^109^, Tanja Jovanovic ^82^, Karen-Inge Karstoft ^18,110^, Milissa L Kaufman ^11,44^, James L Kennedy ^111,112,113,114^, Ronald C Kessler ^115^, Alaptagin Khan ^11,44^, Nathan A Kimbrel ^32,34,116^, Anthony P King ^117^, Nastassja Koen ^118^, Roman Kotov ^119^, Henry R Kranzler ^120,121^, Kristi Krebs ^122^, William S Kremen ^1^, Pei-Fen Kuan ^123^, Bruce R Lawford ^124^, Lauren A M Lebois ^11,12^, Kelli Lehto ^122^, Daniel F Levey ^14,15^, Catrin Lewis ^42^, Israel Liberzon ^125^, Sarah D Linnstaedt ^126^, Mark W Logue ^86,127,128^, Adriana Lori ^82^, Yi Lu ^129^, Benjamin J Luft ^130^, Michelle K Lupton ^64^, Jurjen J Luykx ^94,131^, Iouri Makotkine^16^, Jessica L Maples-Keller ^82^, Shelby Marchese ^132^, Charles Marmar ^133^, Nicholas G Martin ^134^, Gabriela A Martínez-Levy ^65^, Kerrie McAloney ^64^, Alexander McFarlane ^135^, Katie A McLaughlin ^136^, Samuel A McLean ^126,137^, Sarah E Medland ^64^, Divya Mehta ^124,138^, Jacquelyn Meyers ^139^, Vasiliki Michopoulos ^82^, Elizabeth A Mikita ^1,2,3^, Lili Milani ^122^, William Milberg ^140^, Mark W Miller ^127,128^, Rajendra A Morey ^141^, Charles Phillip Morris ^124^, Ole Mors ^52,142^, Preben Bo Mortensen ^52,53,143,144^, Mary S Mufford ^145^, Elliot C Nelson ^41^, Merete Nordentoft ^52,146^, Sonya B Norman ^1,2,147^, Nicole R Nugent ^148,149,150^, Meaghan O'Donnell ^151^, Holly K Orcutt ^152^, Pedro M Pan ^153^, Matthew S Panizzon ^1^, Gita A Pathak ^14,15^, Edward S Peters ^154^, Alan L Peterson ^155,156^, Matthew Peverill ^157^, Robert H Pietrzak ^15,158^, Melissa A Polusny ^21,72,159^, Bernice Porjesz ^139^, Abigail Powers ^82^, Xue-Jun Qin ^23^, Andrew Ratanatharathorn ^6,160^, Victoria B Risbrough ^1,2,3^, Andrea L Roberts ^161^, Alex O Rothbaum ^162,163^, Barbara O Rothbaum ^82^, Peter Roy-Byrne ^164^, Kenneth J Ruggiero ^165^, Ariane Rung ^166^, Heiko Runz ^167^, Bart P F Rutten ^168^, Stacey Saenz de Viteri ^169^, Giovanni Abrahão Salum ^170,171^, Laura Sampson ^6,87^, Sixto E Sanchez ^172^, Marcos Santoro ^173^, Carina Seah ^132^, Soraya Seedat ^174,175^, Julia S Seng ^176,177,178,179^, Andrey Shabalin ^74^, Christina M Sheerin ^17^, Derrick Silove ^180^, Alicia K Smith ^82,181^, Jordan W Smoller ^7,10,182^, Scott R Sponheim ^21,183^, Dan J Stein ^118^, Synne Stensland ^56,77^, Jennifer S Stevens ^82^, Jennifer A Sumner ^184^, Martin H Teicher ^11,185^, Wesley K Thompson ^186,187^, Arun K Tiwari ^111,112,113^, Edward Trapido ^166^, Monica Uddin ^188^, Robert J Ursano ^189^, Unnur Valdimarsdóttir ^190,191^, Miranda Van Hooff ^192^, Eric Vermetten ^193,194,195^, Christiaan H Vinkers ^196,197,198^, Joanne Voisey ^124,138^, Yunpeng Wang ^199^, Zhewu Wang ^200,201^, Monika Waszczuk ^202^, Heike Weber ^69^, Frank R Wendt ^203^, Thomas Werge ^52,204,205,206^, Michelle A Williams ^6^, Douglas E Williamson ^32,33^, Bendik S Winsvold ^55,56,207^, Sherry Winternitz ^11,44^, Christiane Wolf ^69^, Erika J Wolf ^128,208^, Yan Xia ^10,105^, Ying Xiong ^129^, Rachel Yehuda ^16,209^, Keith A Young ^210,211^, Ross McD Young ^212,213^, Clement C Zai ^10,111,112,113,114,214^, Gwyneth C Zai ^111,112,113,114,215^, Mark Zervas ^99^, Hongyu Zhao ^216^, Lori A Zoellner ^157^, John-Anker Zwart ^20,55,56^, Terri deRoon-Cassini ^45^, Sanne J H van Rooij ^82^, Leigh L van den Heuvel ^101,102^; AURORA Study, Estonian Biobank Research Team, FinnGen Investigators, HUNT All-In Psychiatry, Murray B Stein ^1,30,217^, Kerry J Ressler ^11,44,82^, Karestan C Koenen ^6,10,182^

^1^University of California San Diego, Department of Psychiatry, La Jolla, California, United States of America.

^2^Veterans Affairs San Diego Healthcare System, Center of Excellence for Stress and Mental Health, San Diego, California, United States of America.

^3^Veterans Affairs San Diego Healthcare System, Research Service, San Diego, California, United States of America.

^4^Baylor College of Medicine, Department of Molecular and Human Genetics, Houston, Texas, United States of America.

^5^Biogen Inc., Translational Sciences, Cambridge, Massachusetts, United States of America.

^6^Harvard T.H. Chan School of Public Health, Department of Epidemiology, Boston, Massachusetts, United States of America.

^7^Massachusetts General Hospital, Department of Psychiatry, Boston, Massachusetts, United States of America.

^8^King's College London, National Institute for Health and Care Research Maudsley Biomedical Research Centre, South London and Maudsley NHS Foundation Trust, London, United Kingdom.

^9^King's College London, Social, Genetic and Developmental Psychiatry Centre, Institute of Psychiatry, Psychology and Neuroscience, London, United Kingdom.

^10^Broad Institute of MIT and Harvard, Stanley Center for Psychiatric Research, Cambridge, Massachusetts, United States of America.

^11^Harvard Medical School, Department of Psychiatry, Boston, Massachusetts, United States of America.

^12^McLean Hospital, Center of Excellence in Depression and Anxiety Disorders, Belmont, Massachusetts, United States of America.

^13^Stanford University, Department of Psychiatry and Behavioral Sciences, Stanford, California, United States of America.

^14^VA Connecticut Healthcare Center, West Haven, Connecticut, United States of America.

^15^Yale University School of Medicine, Department of Psychiatry, New Haven, Connecticut, United States of America.

^16^Icahn School of Medicine at Mount Sinai, Department of Psychiatry, New York, New York, United States of America.

^17^Virginia Institute for Psychiatric and Behavioral Genetics, Department of Psychiatry, Richmond, Virginia, United States of America.

^18^The Danish Veteran Centre, Research and Knowledge Centre, Ringsted, Sjaelland, Denmark.

^19^Oslo University Hospital, Division of Mental Health and Addiction, Oslo, Norway.

^20^University of Oslo, Institute of Clinical Medicine, Oslo, Norway.

^21^Minneapolis VA Health Care System, Mental Health Service Line, Minneapolis, Minnesota, United States of America.

^22^University of Minnesota, Department of Psychiatry, Minneapolis, Minnesota, United States of America.

^23^Duke University, Duke Molecular Physiology Institute, Durham, North Carolina, United States of America.

^24^Boston Children's Hospital, Division of Adolescent and Young Adult Medicine, Boston, Massachusetts, United States of America.

^25^Harvard Medical School, Department of Pediatrics, Boston, Massachusetts, United States of America.

^26^Harvard T.H. Chan School of Public Health, Department of Social and Behavioral Sciences, Boston, Massachusetts, United States of America.

^27^University Clinical Center of Tuzla, Department of Psychiatry, Tuzla, Bosnia and Herzegovina.

^28^University Clinical Center of Mostar, Department of Psychiatry, Mostar, Bosnia and Herzegovina.

^29^Virginia Commonwealth University, Department of Psychiatry, Richmond, Virginia, United States of America.

^30^Veterans Affairs San Diego Healthcare System, Psychiatry Service, San Diego, California, United States of America.

^31^Mayo Clinic, Department of Quantitative Health Sciences, Rochester, Minnesota, United States of America.

^32^Duke University School of Medicine, Department of Psychiatry and Behavioral Sciences, Durham, North Carolina, United States of America.

^33^Durham VA Health Care System, Research, Durham, North Carolina, United States of America.

^34^VA Mid-Atlantic Mental Illness Research, Education, and Clinical Center (MIRECC), Genetics Research Laboratory, Durham, North Carolina, United States of America.

^35^Universidade Federal de São Paulo, Department of Morphology and Genetics, São Paulo, São Paulo, Brazil.

^36^Universidade Federal de São Paulo, Laboratory of Integrative Neuroscience, Department of Psychiatry, São Paulo, São Paulo, Brazil.

^37^Instituto Nacional de Psiquiatraía Ramón de la Fuente Muñiz, Center for Global Mental Health, Mexico City, Mexico City, Mexico.

^38^Medical College of Wisconsin, Comprehensive Injury Center, Milwaukee, Wisconsin, United States of America.

^39^James J. Peters VA Medical Center, Department of Psychiatry, Bronx, New York, United States of America.

^40^Mayo Clinic, Department of Psychiatry and Psychology, Rochester, Minnesota, United States of America.

^41^Washington University in Saint Louis School of Medicine, Department of Psychiatry, Saint Louis, Missouri, United States of America.

^42^Cardiff University, National Centre for Mental Health, MRC Centre for Psychiatric Genetics and Genomics, Cardiff, South Glamorgan, United Kingdom.

^43^Brain Center University Medical Center Utrecht, Department of Psychiatry, Utrecht, Utrecht, The Netherlands.

^44^McLean Hospital, Belmont, Massachusetts, United States of America.

^45^Medical College of Wisconsin, Department of Surgery, Division of Trauma & Acute Care Surgery, Milwaukee, Wisconsin, United States of America.

^46^King's College London, NIHR Maudsley BRC, London, United Kingdom.

^47^Universidade Federal de São Paulo, Department of Psychiatry, São Paulo, São Paulo, Brazil.

^48^Universidade Federal de São Paulo, Laboratory of Integrative Neuroscience, Department of Psychiatry, São Paulo, São Paulo, Brazil.

^49^University of New South Wales, School of Psychology, Sydney, New South Wales, Australia.

^50^University of Michigan Medical School, Division of Pulmonary and Critical Care Medicine, Department of Internal Medicine, Ann Arbor, Michigan, United States of America.

^51^Statens Serum Institut, Department for Congenital Disorders, Copenhagen, Denmark.

^52^The Lundbeck Foundation Initiative for Integrative Psychiatric Research, iPSYCH, Aarhus, Denmark.

^53^Aarhus University, Centre for Integrative Sequencing, iSEQ, Aarhus, Denmark.

^54^Aarhus University, Department of Biomedicine - Human Genetics, Aarhus, Denmark.

^55^Norwegian University of Science and Technology, K. G. Jebsen Center for Genetic Epidemiology, Department of Public Health and Nursing, Faculty of Medicine and Health Sciences, Trondheim, Norway.

^56^Oslo University Hospital, Department of Research, Innovation and Education, Division of Clinical Neuroscience, Oslo, Norway.

^57^Case Western Reserve University, School of Medicine, Cleveland, Ohio, United States of America.

^58^University Hospitals, Department of Psychiatry, Cleveland, Ohio, United States of America.

^59^Chronic Diseases Research Centre (CEDOC), Lisbon Institute of Global Mental Health, Lisbon, Portugal.

^60^McLean Hospital, Division of Depression and Anxiety Disorders, Belmont, Massachusetts, United States of America.

^61^University of Toronto, CanPath National Coordinating Center, Toronto, Ontario, Canada.

^62^Stony Brook University, Family, Population, and Preventive Medicine, Stony Brook, New York, United States of America.

^63^Stony Brook University, Public Health, Stony Brook, New York, United States of America.

^64^QIMR Berghofer Medical Research Institute, Mental Health & Neuroscience Program, Brisbane, Queensland, Australia.

^65^Instituto Nacional de Psiquiatraía Ramón de la Fuente Muñiz, Department of Genetics, Mexico City, Mexico City, Mexico.

^66^University of California San Diego, Department of Radiology, Department of Neurosciences, La Jolla, California, United States of America.

^67^University of Cape Town, Division of Human Genetics, Department of Pathology, Cape Town, Western Province, South Africa.

^68^Vanderbilt University Medical Center, Vanderbilt Genetics Institute, Nashville, Tennessee, United States of America.

^69^University Hospital of Würzburg, Center of Mental Health, Psychiatry, Psychosomatics and Psychotherapy, Würzburg, Denmark.

^70^Kent State University, Department of Psychological Sciences, Kent, Ohio, United States of America.

^71^Minneapolis VA Health Care System, Research Service Line, Minneapolis, Minnesota, United States of America.

^72^University of Minnesota Medical School, Department of Psychiatry & Behavioral Sciences, Minneapolis, Minnesota, United States of America.

^73^Huntsman Mental Health Institute, Salt Lake City, Utah, United States of America.

^74^University of Utah School of Medicine, Department of Psychiatry, Salt Lake City, Utah, United States of America.

^75^University of Freiburg, Faculty of Medicine, Centre for Basics in Neuromodulation, Freiburg, Denmark.

^76^University of Freiburg, Faculty of Medicine, Department of Psychiatry and Psychotherapy, Freiburg, Denmark.

^77^Norwegian Centre for Violence and Traumatic Stress Studies, Oslo, Norway.

^78^University Clinical Center of Sarajevo, Department of Psychiatry, Sarajevo, Bosnia and Herzegovina.

^79^Indiana University School of Medicine, Biochemistry and Molecular Biology, Indianapolis, Indiana, United States of America.

^80^Indiana University School of Medicine, Medical and Molecular Genetics, Indianapolis, Indiana, United States of America.

^81^University of Bologna, Department of Biomedical and Neuromotor Sciences, Bologna, Italy.

^82^Emory University, Department of Psychiatry and Behavioral Sciences, Atlanta, Georgia, United States of America.

^83^Boston University Chobanian & Avedisian School of Medicine, Department of Medicine (Biomedical Genetics), Boston, Massachusetts, United States of America.

^84^Boston University Chobanian & Avedisian School of Medicine, Department of Neurology, Boston, Massachusetts, United States of America.

^85^Boston University Chobanian & Avedisian School of Medicine, Department of Ophthalmology, Boston, Massachusetts, United States of America.

^86^Boston University School of Public Health, Department of Biostatistics, Boston, Massachusetts, United States of America.

^87^Boston University School of Public Health, Department of Epidemiology, Boston, Massachusetts, United States of America.

^88^Case Western Reserve University, Department of Psychological Sciences, Cleveland, Ohio, United States of America.

^89^University of Melbourne, Department of Psychiatry, Melbourne, Victoria, Australia.

^90^Boston University School of Public Health, Boston, Massachusetts, United States of America.

^91^VA Connecticut Healthcare Center, Psychiatry Service, West Haven, Connecticut, United States of America.

^92^Yale University School of Medicine, Department of Genetics and Neuroscience, New Haven, Connecticut, United States of America.

^93^Netherlands Ministry of Defence, Brain Research and Innovation Centre, Utrecht, Utrecht, The Netherlands.

^94^UMC Utrecht Brain Center Rudolf Magnus, Department of Psychiatry, Utrecht, Utrecht, The Netherlands.

^95^National Institutes of Health, National Human Genome Research Institute, Bethesda, Maryland, United States of America.

^96^University Clinical Centre of Kosovo, Department of Psychiatry, Prishtina, Kosovo.

^97^Wayne State University School of Medicine, Psychiatry and Behavioral Neurosciences, Detroit, Michigan, United States of America.

^98^Gallipoli Medical Research Foundation, Greenslopes Private Hospital, Greenslopes, Queensland, Australia.

^99^Cohen Veterans Bioscience, New York, New York, United States of America.

^100^Washington University in Saint Louis School of Medicine, Department of Genetics, Saint Louis, Missouri, United States of America.

^101^Stellenbosch University, Department of Psychiatry, Faculty of Medicine and Health Sciences, Cape Town, Western Cape, South Africa.

^102^Stellenbosch University, SAMRC Genomics of Brain Disorders Research Unit, Cape Town, Western Cape, South Africa.

^103^University of Connecticut School of Medicine, Psychiatry, Farmington, Connecticut, United States of America.

^104^University of Sydney, Brain and Mind Centre, Sydney, New South Wales, Australia.

^105^Massachusetts General Hospital, Analytic and Translational Genetics Unit, Department of Medicine, Boston, Massachusetts, United States of America.

^106^Yale University, Department of Psychiatry, New Haven, Connecticut, United States of America.

^107^University Hospital Center of Zagreb, Department of Psychiatry, Zagreb, Croatia.

^108^Icahn School of Medicine at Mount Sinai, Genetics and Genomic Sciences, New York, New York, United States of America.

^109^Cardiff University, National Centre for Mental Health, Cardiff University Centre for Psychiatric Genetics and Genomics, Cardiff, South Glamorgan, United Kingdom.

^110^University of Copenhagen, Department of Psychology, Copenhagen, Denmark.

^111^Centre for Addiction and Mental Health, Neurogenetics Section, Molecular Brain Science Department, Campbell Family Mental Health Research Institute, Toronto, Ontario, Canada

^112^Centre for Addiction and Mental Health, Tanenbaum Centre for Pharmacogenetics, Toronto, Ontario, Canada.

^113^University of Toronto, Department of Psychiatry, Toronto, Ontario, Canada

^114^University of Toronto, Institute of Medical Sciences, Toronto, Ontario, Canada.

^115^Harvard Medical School, Department of Health Care Policy, Boston, Massachusetts, United States of America.

^116^Durham VA Health Care System, Mental Health Service Line, Durham, North Carolina, United States of America.

^117^The Ohio State University, College of Medicine, Institute for Behavioral Medicine Research, Columbus, Ohio, United States of America.

^118^University of Cape Town, Department of Psychiatry & Neuroscience Institute, SA MRC Unit on Risk & Resilience in Mental Disorders, Cape Town, Western Province, South Africa.

^119^Stony Brook University, Department of Psychiatry, Stony Brook, New York, United States of America.

^120^Mental Illness Research, Education and Clinical Center, Crescenz VAMC, Philadelphia, Pennsylvania, United States of America.

^121^University of Pennsylvania Perelman School of Medicine, Department of Psychiatry, Philadelphia, Pennsylvania, United States of America.

^122^University of Tartu, Institute of Genomics, Estonian Genome Center, Tartu, Estonia.

^123^Stony Brook University, Department of Applied Mathematics and Statistics, Stony Brook, New York, United States of America.

^124^Queensland University of Technology, School of Biomedical Sciences, Kelvin Grove, Queensland, Australia.

^125^Texas A&M University College of Medicine, Department of Psychiatry and Behavioral Sciences, Bryan, Texas, United States of America.

^126^UNC Institute for Trauma Recovery, Department of Anesthesiology, Chapel Hill, North Carolina, United States of America.

^127^Boston University School of Medicine, Psychiatry, Biomedical Genetics, Boston, Massachusetts, United States of America.

^128^VA Boston Healthcare System, National Center for PTSD, Boston, Massachusetts, United States of America.

^129^Karolinska Institutet, Department of Medical Epidemiology and Biostatistics, Stockholm, Sweden.

^130^Stony Brook University, Department of Medicine, Stony Brook, New York, United States of America.

^131^UMC Utrecht Brain Center Rudolf Magnus, Department of Translational Neuroscience, Utrecht, Utrecht, The Netherlands.

^132^Icahn School of Medicine at Mount Sinai, Department of Genetic and Genomic Sciences, New York, New York, United States of America.

^133^New York University, Grossman School of Medicine, New York, New York, United States of America.

^134^QIMR Berghofer Medical Research Institute, Genetics, Brisbane, Queensland, Australia.

^135^University of Adelaide, Discipline of Psychiatry, Adelaide, South Australia, Australia.

^136^Harvard University, Department of Psychology, Boston, Massachusetts, United States of America.

^137^UNC Institute for Trauma Recovery, Department of Emergency Medicine, Chapel Hill, North Carolina, United States of America.

^138^Queensland University of Technology, Centre for Genomics and Personalised Health, Kelvin Grove, Queensland, Australia.

^139^SUNY Downstate Health Sciences University, Department of Psychiatry and Behavioral Sciences, Brooklyn, New York, United States of America.

^140^VA Boston Healthcare System, GRECC/TRACTS, Boston, Massachusetts, United States of America.

^141^Duke University School of Medicine, Duke Brain Imaging and Analysis Center, Durham, North Carolina, United States of America.

^142^Aarhus University Hospital - Psychiatry, Psychosis Research Unit, Aarhus, Denmark.

^143^Aarhus University, Centre for Integrated Register-based Research, Aarhus, Denmark.

^144^Aarhus University, National Centre for Register-Based Research, Aarhus, Denmark.

^145^University of Cape Town, Division of Human Genetics, Department of Pathology, Cape Town, Western Province, South Africa.

^146^University of Copenhagen, Mental Health Services in the Capital Region of Denmark, Copenhagen, Denmark.

^147^National Center for Post Traumatic Stress Disorder, Executive Division, White River Junction, Vermont, United States of America.

^148^Alpert Brown Medical School, Department of Emergency Medicine, Providence, Rhode Island, United States of America.

^149^Alpert Brown Medical School, Department of Pediatrics, Providence, Rhode Island, United States of America.

^150^Alpert Brown Medical School, Department of Psychiatry and Human Behavior, Providence, Rhode Island, United States of America.

^151^University of Melbourne, Phoenix Australia, Department of Psychiatry, Melbourne, Victoria, Australia.

^152^Northern Illinois University, Department of Psychology, DeKalb, Illinois, United States of America.

^153^Universidade Federal de São Paulo, Psychiatry, São Paulo, São Paulo, Brazil.

^154^University of Nebraska Medical Center, College of Public Health, Omaha, Nebraska, United States of America.

^155^South Texas Veterans Health Care System, Research and Development Service, San Antonio, Texas, United States of America.

^156^University of Texas Health Science Center at San Antonio, Department of Psychiatry and Behavioral Sciences, San Antonio, Texas, United States of America.

^157^University of Washington, Department of Psychology, Seattle, Washington, United States of America.

^158^U.S. Department of Veterans Affairs National Center for Posttraumatic Stress Disorder, West Haven, Connecticut, United States of America.

^159^Center for Care Delivery and Outcomes Research (CCDOR), Minneapolis, Minnesota, United States of America.

^160^Columbia University Mailmain School of Public Health, Department of Epidemiology, New York, New York, United States of America.

^161^Harvard T.H. Chan School of Public Health, Department of Environmental Health, Boston, Massachusetts, United States of America.

^162^Emory University, Department of Psychological Sciences, Atlanta, Georgia, United States of America.

^163^Skyland Trail, Department of Research and Outcomes, Atlanta, Georgia, United States of America.

^164^University of Washington, Department of Psychiatry, Seattle, Washington, United States of America.

^165^Medical University of South Carolina, Department of Nursing and Department of Psychiatry, Charleston, South Carolina, United States of America.

^166^Louisiana State University Health Sciences Center, School of Public Health and Department of Epidemiology, New Orleans, Louisiana, United States of America.

^167^Biogen Inc., Research & Development, Cambridge, Massachusetts, United States of America.

^168^Maastricht Universitair Medisch Centrum, School for Mental Health and Neuroscience, Department of Psychiatry and Neuropsychology, Maastricht, Limburg, The Netherlands.

^169^SUNY Downstate Health Sciences University, School of Public Health, Brooklyn, New York, United States of America.

^170^Child Mind Institute, New York, New York, United States of America.

^171^Instituto Nacional de Psiquiatria de Desenvolvimento, São Paulo, São Paulo, Brazil.

^172^Universidad Peruana de Ciencias Aplicadas, Department of Medicine, Lima, Lima, Peru.

^173^Universidade Federal de São Paulo, Departamento de Bioquímica - Disciplina de Biologia Molecular, São Paulo, São Paulo, Brazil.

^174^Stellenbosch University, Department of Psychiatry, Faculty of Medicine and Health Sciences, Stellenbosch University, Cape Town, Western Cape, South Africa.

^175^Stellenbosch University, SAMRC Extramural Genomics of Brain Disorders Research Unit, Cape Town, Western Cape, South Africa.

^176^University of Michigan, Department of Obstetrics and Gynecology, Ann Arbor, Michigan, United States of America.

^177^University of Michigan, Department of Women's and Gender Studies, Ann Arbor, Michigan, United States of America.

^178^University of Michigan, Institute for Research on Women and Gender, Ann Arbor, Michigan, United States of America.

^179^University of Michigan, School of Nursing, Ann Arbor, Michigan, United States of America.

^180^University of New South Wales, Department of Psychiatry, Sydney, New South Wales, Australia.

^181^Emory University, Department of Gynecology and Obstetrics; Department of Psychiatry and Behavioral Sciences; Department of Human Genetics, Atlanta, Georgia, United States of America.

^182^Massachusetts General Hospital, Psychiatric and Neurodevelopmental Genetics Unit (PNGU), Boston, Massachusetts, United States of America.

^183^University of Minnesota Medical School, Department of Psychiatry and Behavioral Sciences, Minneapolis, Minnesota, United States of America.

^184^University of California, Los Angeles, Department of Psychology, Los Angeles, California, United States of America.

^185^McLean Hospital, Developmental Biopsychiatry Research Program, Belmont, Massachusetts, United States of America.

^186^Mental Health Centre Sct. Hans, Institute of Biological Psychiatry, Roskilde, Denmark.

^187^University of California San Diego, Herbert Wertheim School of Public Health and Human Longevity Science, La Jolla, California, United States of America.

^188^University of South Florida College of Public Health, Genomics Program, Tampa, Florida, United States of America.

^189^Uniformed Services University, Department of Psychiatry, Bethesda, Maryland, United States of America.

^190^Karolinska Institutet, Unit of Integrative Epidemiology, Institute of Environmental Medicine, Stockholm, Sweden.

^191^University of Iceland, Faculty of Medicine, Center of Public Health Sciences, School of Health Sciences, Reykjavik, Iceland.

^192^University of Adelaide, Adelaide Medical School, Adelaide, South Australia, Australia.

^193^ARQ Nationaal Psychotrauma Centrum, Psychotrauma Reseach Expert Group, Diemen, North Holland, The Netherlands

^194^Leiden University Medical Center, Department of Psychiatry, Leiden, South Holland, The Netherlands.

^195^New York University School of Medicine, Department of Psychiatry, New York, New York, United States of America.

^196^Amsterdam Neuroscience, Mood, Anxiety, Psychosis, Sleep & Stress Program, Amsterdam, North Holland, The Netherlands.

^197^Amsterdam UMC location Vrije Universiteit Amsterdam, Department of Anatomy and Neurosciences, Amsterdam, North Holland, The Netherlands.

^198^Amsterdam UMC location Vrije Universiteit Amsterdam, Department of Psychiatry, Amsterdam, North Holland, The Netherlands.

^199^University of Oslo, Lifespan Changes in Brain and Cognition (LCBC), Department of Psychology, Oslo, Norway.

^200^Medical University of South Carolina, Department of Psychiatry and Behavioral Sciences, Charleston, South Carolina, United States of America.

^201^Ralph H Johnson VA Medical Center, Department of Mental Health, Charleston, South Carolina, United States of America.

^202^Rosalind Franklin University of Medicine and Science, Department of Psychology, North Chicago, Illinois, United States of America.

^203^University of Toronto, Dalla Lana School of Public Health, Department of Anthropology, Toronto, Ontario, Canada.

^204^Copenhagen University Hospital, Institute of Biological Psychiatry, Mental Health Services, Copenhagen, Denmark.

^205^University of Copenhagen, Department of Clinical Medicine, Copenhagen, Denmark.

^206^University of Copenhagen, The Globe Institute, Lundbeck Foundation Center for Geogenetics, Copenhagen, Denmark.

^207^Oslo University Hospital, Department of Neurology, Oslo, Norway.

^208^Boston University Chobanian & Avedisian School of Medicine, Department of Psychiatry, Boston, Massachusetts, United States of America.

^209^James J. Peters VA Medical Center, Department of Mental Health, Bronx, New York, United States of America.

^210^Central Texas Veterans Health Care System, Research Service, Temple, Texas, United States of America.

^211^Texas A&M University School of Medicine, Department of Psychiatry and Behavioral Sciences, Bryan, Texas, United States of America.

^212^Queensland University of Technology, School of Clinical Sciences, Kelvin Grove, Queensland, Australia.

^213^University of the Sunshine Coast, The Chancellory, Sippy Downs, Queensland, Australia.

^214^University of Toronto, Department of Laboratory Medicine and Pathology, Toronto, Ontario, Canada.

^215^Centre for Addiction and Mental Health, General Adult Psychiatry and Health Systems Division, Toronto, Ontario, Canada.

^216^Yale University, Department of Biostatistics, New Haven, Connecticut, United States of America.

^217^University of California San Diego, School of Public Health, La Jolla, California, United States of America.

^*^Contributed equally

Author Contributions

PGC-PTSD writing group: E.G.A., S.-A.B., C.-Y.C., K.W.C., J.R.I.C., N.P.D., L.E.D., K.C.K., A.X.M., R.A.M., C.M.N., R.P., K.J.R., and M.B.S.

Study PI or co-PI: A.B.A., S.B. Andersen, P.A.A., A.E.A.-K., S.B. Austin, E.A., D.B., D.G.B., J.C.B., S. Belangero, C. Benjet, J.M.B., L.J.B., J.I.B., G.B., R.B., A.D.B., J.R.C., C.S.C., L.K.B., J.D., D.L.D., T.d.-C., K.D., G.D., A.D.-K., N.F., L.A.F., A.F., N.C.F., B.G., J.G., E.G., C.F.G., A.G.U., M.A.H., A.C.H., V.H., I.B.H., D.M.H., K. Hveem, M. Jakovljevic, A.J., I.J., T.J., K.-I.K., M.L.K., R.C.K., N.A.K., K.C.K., R.K., H.R.K., W.S.K., B.R.L., K.L., I.L., B.L., C.M., N.G.M., K.A.M., S.A.M., S.E.M., D.M., W.P.M., M.W.M., C.P.M., O.M., P.B.M, E.C.N., C.M.N., M.N., S.B.N., N.R.N., P.M.P., A.L.P., R.H.P., M.A.P., B.P., A.P., K.J.R., V.R., P.R.B., K.R., H.R., G.S., S. Seedat, J.S. Seng, A.K.S., S.R.S., D.J.S., M.B.S., R.J.U., U.V., S.J.H.v.R., E.V., J.V., Z.W., M.W., H.W., T.W., M.A.W., D.E.W., C.W., R.M.Y., H.Z., L.A.Z., and J.-A.Z.

Obtained funding for studies: A.B.A., P.A.A., A.E.A.-K., S.B. Austin, J.C.B., S. Belangero, C. Benjet, J.M.B., L.J.B., G.B., A.D.B., C.S.C., J.D., T.d.-C., A.F., N.C.F., J.D.F., C.E.F., E.G., C.F.G., M.H., M.A.H., A.C.H., V.H., I.B.H., D.M.H., K. Hveem, T.J., N.A.K., K.C.K., R.K., W.S.K., B.R.L., B.L., C.M., N.G.M., K.A.M., S.A.M., S.E.M., J.M., W.P.M., M.W.M., C.P.M., O.M., P.B.M, E.C.N., C.M.N., M.N., N.R.N., H.K.O., M.A.P., B.P., K.J.R., B.O.R., G.S., M.S., A.K.S., S.R.S., M.H.T., R.J.U., U.V., E.V., J.V., Z.W., M.W., T.W., M.A.W., D.E.W., R.Y., R.M.Y., and L.A.Z.

Clinical: C.A., P.A.A., E.A., D.B., D.G.B., J.C.B., L.B., L.J.B., E.A.B., R.B., A.C.B., A.D.B., S. Børte, L.C., J.R.C., K.W.C., L.K.B., M.F.D., T.d.-C., S.G.D., G.D., A.D.-K., N.F., N.C.F., J.D.F., C.E.F., S.G., E.G., A.G.U., S.B.G., L.G., C.G., V.H., D.M.H., M. Jakovljevic, A.J., G.D.J., M.L.K., A.K., N.A.K., N.K., R.K., W.S.K., B.R.L., L.A.M.L., K.L., C.E.L., B.L., J.L.M.-K., S.A.M., P.B.M., H.K.O., P.M.P., M.S.P., E.S.P., A.L.P., M.P., R.H.P., M.A.P., B.P., A.P., B.O.R., A.O.R., G.S., L.S., J.S. Seng, C.M.S., S. Stensland, M.H.T., W.K.T., E.T., M.U., U.V., L.L.v.d.H., E.V., Z.W., Y.W., T.W., D.E.W., B.S.W., S.W., E.J.W., R.Y., K.A.Y., and L.A.Z.

Contributed data: O.A.A., P.A.A., S.B. Austin, D.G.B., S. Belangero, L.J.B., R.B., R.A.B., A.D.B., J.R.C., J.M.C.-d.-A., S.Y.C., S.A.P.C., A.M.D., L.K.B., D.L.D., A.E., N.C.F., D.F., C.E.F., S.G., B.G., S.M.J.H., D.M.H., L.M.H., K. Hveem, A.J., I.J., M.L.K., J.L.K., R.C.K., A.P.K., R.K., W.S.K., L.A.M.L., K.L., D.F.L., C.E.L., I.L., B.L., M.K.L., S.M., G.A.M., K.M., A.M., K.A.M., S.E.M., J.M., L.M., O.M., P.B.M., M.N., S.B.N., N.R.N., M.O., P.M.P., M.S.P., E.S.P., A.L.P., M.P., R.H.P., M.A.P., K.J.R., V.R., P.R.B., A. Rung, G.S., L.S., S.E.S., M.S., C.S., S. Seedat, J.S. Seng, D. Silove, J.W.S., S.R.S., M.B.S., A.K.T., E.T., U.V., L.L.v.d.H., M.V.H., M.W., T.W., D.E.W., S.W., K.A.Y., C.C.Z., G.C.Z., L.A.Z., and J.-A.Z.

Statistical analysis: A.E.A.-K., A. Batzler, C. Bergner, A. Brandolino, S. Børte, C.C., C.-Y.C., S.A.P.C., J.R.I.C., L.C.-C., B.J.C., S.D., S.G.D., A.D., L.E.D., C.F., M.E.G., B.G., S.B.G., S.D.G., C.G., S.H., E.M.H., K. Hogan, H.H., G.D.J., K.K., P.-F.K., D.F.L., M.W.L., A.L, Y.L., A.X.M., S.M., C.M., D.M., J.M., V.M., E.A.M., M.S.M., C.M.N., G.A.P., M.P., X.-J.Q., A.R., A.L.R., S.S.d.V., C.S., A.S., C.M.S., S. Stensland, J.S.S., J.A.S., F.R.W., B.S.W., Y. Xia, Y. Xiong, and C.C.Z.

Bioinformatics: A.E.A.-K., A. Batzler, M.P.B., S. Børte, C.C., C.-Y.C., J.R.I.C., N.P.D., C.D.P., S.G.D., A.D., H.E., M.E.G., K. Hogan, H.H., K.K., P.-F.K., D.F.L., S.D.L., A.L, A.X.M., G.A.M., D.M., J.M., V.M., E.A.M., G.A.P., A.R., A.S., J.S.S., F.R.W., B.S.W., C.W., Y. Xia, Y. Xiong, and C.C.Z.

Genomics: M.P.B., J.B.-G., M.B.-H., N.P.D., T.d.-C., F.D., A.D., K.D., H.E., L.G., M.A.H., J.J., P.-F.K., S.D.L., J.J.L., I.K., J.M., L.M., K.J.R., B.P.F.R., S.S.d.V., A.S., C.H.V., and D.E.W.

PGC-PTSD management group: M.H. and M.Z.
